# Pre-existing anti-polyethylene glycol antibodies in pregnant women and newborns

**DOI:** 10.1101/2024.11.29.24317450

**Authors:** Haiyang Wang, Yan Feng, Lin Zhang, Changzheng Yuan, Junyang Xue, Jicheng Li, Xiao Xu, Wenbin Zhou, Baohua Li, Yisha Wang, Gan Luo, Yue Zheng, Meihua Sui

**Affiliations:** School of Basic Medical Sciences, Zhejiang University School of Medicine, Hangzhou 310058, China; Women’s Hospital, Zhejiang University School of Medicine, Hangzhou 311500, China; Shaoxing People’s Hospital (Shaoxing Hospital, Zhejiang University School of Medicine), Shaoxing 312300, China; School of Public Health, Zhejiang University School of Medicine, Hangzhou 310058, China; Department of Hepatobiliary and Pancreatic Surgery, People’s Hospital of Hangzhou Medical College, Hangzhou 310014, China

**Keywords:** Pre-existing anti-polyethylene glycol (PEG) antibodies, Pregnant women, Newborns, Seroepidemiological characteristics, Influencing factors

## Abstract

Pre-existing anti-polyethylene glycol (PEG) antibodies represent risk factors for reduced efficacy and increased adverse reactions in seropositive individuals, but neither the seropositivities, nor levels nor influencing factors have been investigated in pregnant women or newborns. Herein, maternal and cord blood samples were respectively collected from 256 pregnant women and corresponding 256 newborns at the Women’s Hospital, Zhejiang University School of Medicine in China for further determination of pre-existing anti-PEG antibodies, along with questionnaire interviews, demographic and clinical data collections. Our data showed that the seropositivities of total anti-PEG antibodies, anti-PEG IgG1 and IgG2, anti-PEG IgM, and coexistence of anti-PEG IgM and IgG were 19.14%, 2.34%, 7.03%, 10.94% and 1.17%, respectively, in pregnant women, and 5.47%, 2.73%, 2.73%, 0% and 0%, respectively, in newborns. Anti-PEG IgG3, IgG4 and IgE were undetectable in all blood samples. Median anti-PEG IgG1, IgG2 and IgM concentrations were 273.88 ng/mL, 748.35 ng/mL and 175.07 ng/mL, respectively, in pregnant women. Median anti-PEG IgG1 and IgG2 concentrations were 207.92 ng/mL and 336.52 ng/mL, respectively, in newborns. Interestingly, in-depth statistical analyses revealed that maternal age, take-out food consumption and cosmetic use were influencing factors of maternal anti-PEG antibodies, while newborn anti-PEG antibodies were affected by maternal age and cosmetic use. These seroepidemiological characteristics raise concerns over the clinical use of PEGylated drugs in pregnant women and newborns, and provide valuable insight into the induction of risky pre-existing anti-PEG antibodies.

## INTRODUCTION

Polyethylene glycol (PEG) is a synthetic polymer comprised of repeating subunits of ethylene glycol and often covalently attached to small molecules, macromolecules and nanomaterials to improve their properties, a process called “PEGylation”^1^. PEGylation has become one of the most preferred methods to enhance the delivery of therapeutic molecules^2^. Currently there are 41 PEGylated medicines approved by the Food and Drug Administration (FDA), including two COVID-19 mRNA vaccines BNT162b2 (Comirnaty^®^) and mRNA-1273 (Spikevax^®^), with many other PEGylated agents under clinical development^3, 4^. In addition, PEG and its derivatives have been approved for use in cosmetics and personal care products, as well as food additives and packaging materials in processed foods and beverages, etc^5, 6^.

Although PEG was initially regarded as non-immunogenic, it has been recognized as a polyvalent hapten^7–9^. Anti-PEG antibodies could be elicited in animals immunized with PEGylated proteins or nanocarriers^7–10^ and in patients treated with PEGylated drugs^11–17^. Surprisingly, normal population actually possesses pre-existing anti-PEG antibodies in the absence of treatment with PEGylated therapeutics^18^, which was firstly documented in 1984^19^. It is noteworthy that the presence of anti-PEG antibodies may represent risk factors for reduced efficacy and adverse reactions in patients requiring treatment with PEGylated drugs^20–22^. Specifically, anti-PEG antibodies may induce the formation of immune complexes with PEG-modified medicines, which could be rapidly cleared via activation of the complement system and subsequent phagocytosis by macrophages, thereby altering the pharmacokinetics and biodistribution^21, 22^. Complement activation may also contribute to the development of infusion-related allergic reactions to PEGylated drugs^21, 22^. For instance, the PEGylated nanoparticles in COVID-19 mRNA vaccine Comirnaty^®^, which could induce anti-PEG antibodies as recently demonstrated by us^23^ (23), have been suspected to trigger allergic reactions after vaccinations^24^.

Up to date, there are seven seroepidemiological studies on pre-existing anti-PEG antibodies in general adults, including Chinese adults in Taiwan of China^25^, American^26–29^, Australian^27^, Japanese^19^, Italian^19^ and German adults^19, 30^, with interesting data obtained (Tables S1 and 2 in Supplementary Appendix; see Supplementary Appendix for all online-only materials, e.g. Methods S1, Results S1, Table S1, Fig. S1 and Discussion S1 in the following sections). Nevertheless, pregnant women represent a unique population with distinctive and dynamic immunological milieus, and they provide passive immunity to their fetuses/newborns^31^. Hence, parallel investigation on the pre-existing anti-PEG antibodies in pregnant women and their newborns has offered an opportunity to reveal the potential maternal-fetal/newborn disparities in addition to identifying their respective seroepidemiological characteristics. Meanwhile, in some circumstances, pregnant women need to be treated with PEGylated drugs for medical conditions arising during pregnancy^32^. Investigating maternal-fetal/newborn anti-PEG antibodies is thus crucial for assessing the maternal-fetal/newborn safety and efficacy of PEGylated drug treatment. Motivated by these concerns, 256 pregnant women and their newborns were enrolled in this cross-sectional study, with corresponding maternal and cord blood samples carefully collected and examined for pre-existing anti-PEG antibodies. Using internationally recognized direct ELISA and competitive ELISA, along with questionnaire interviews, demographic and clinical data collections, and in-depth statistical analysis, the characteristics of pre-existing maternal and newborn anti-PEG antibodies, and the potential independent influencing factors were investigated.

## RESULTS

### Study Population

A total of 256 pregnant women enrolled had a median (IQR) age of 32 (29-36) years and a median (IQR) BMI of 26.70 (24.80-29.00) kg/m^2^. The education levels of participants were as follows: 50 (19.53%) with vocational degrees or below; 61 (23.83%) with associate’s degrees; 107 (41.80%) with bachelor’s degrees; 38 (14.84%) with master’s degrees or above. Moreover, 182 (71.09%) resided in cities while 74 (28.91%) lived in towns. Regarding cosmetic use, 205 (80.09%), 31 (12.11%) and 20 (7.81%) respectively reported 0 days, 1-3 days and 4-7 days for each week. In addition, 70 (27.34%), 97 (37.89%), 64 (25.00%) and 25 (9.77%) respectively reported 0 times, 1-3 times, 4-6 times and 7-9 times take-out food consumption for each week. All newborns were delivered at a mean (SD) gestational age of 38.45 (0.86) weeks, with a mean (SD) birth weight of 3.32 (0.41) kg. Moreover, 127 newborns (49.61%) were males and 129 (50.39%) were females (Table 1).

**Table 1.** Demographic and Clinical Characteristics of Pregnant Women.

| Characteristic | Participant (n = 256) |
| --- | --- |
| **Data on pregnant women** |  |
| Maternal age, median (IQR), y | 32 (29-36) |
| BMI, median (IQR), kg/m<sup>2</sup> | 26.70 (24.80-29.00) |
| Education, No. (%) |  |
| Vocational degree or below<sup>a</sup> | 50 (19.53) |
| Associate's degree | 61 (23.83) |
| Bachelor's degree | 107 (41.80) |
| Master's degree or above | 38 (14.84) |
| Permanent address, No. (%) |  |
| Town | 74 (28.91) |
| City | 182 (71.09) |
| Cosmetic use (per week), No. (%) |  |
| 0 days | 205 (80.09) |
| 1-3 days | 31 (12.11) |
| 4-7 days | 20 (7.81) |
| Take-out food consumption (per week), No. (%) |  |
| 0 times | 70 (27.34) |
| 1-3 times | 97 (37.89) |
| 4-6 times | 64 (25.00) |
| 7-9 times | 25 (9.77) |
| **Data on newborns** |  |
| Gestational age at delivery, mean (SD), wk | 38.45 (0.86) |
| Newborn gender, No. (%) |  |
| Male | 127 (49.61) |
| Female | 129 (50.39) |
| Newborn weight, mean (SD), kg | 3.32 (0.41) |

Abbreviations: BMI, body mass index (calculated as weight in kilograms divided by height in meters squared); IQR, interquartile range; SD, standard deviation.
<sup>a</sup>Elementary school or junior high school (6 cases), high school or technical secondary school (44 cases).

### Prevalence and levels of Pre-existing Anti-PEG Antibodies in Pregnant Women and Newborns

By using direct ELISA (Tables S3; Figs. S1-24) and subsequent competitive ELISA (Table S4; Figs. S25-36), anti-PEG antibody seropositive samples were verified and quantified according to standard curves (Figs. S37-42). Our data showed that anti-PEG antibodies were detectable in 49 (19.14%) pregnant women, with 2.34%, 7.03%, 10.94% and 1.17% of all pregnant women respectively positive for anti-PEG IgG1, IgG2, IgM, and both IgG1 and IgM. No pregnant woman was positive for anti-PEG IgG3, IgG4 or IgE. In addition, double-positivity neither for IgG1 and IgG2, nor for IgG2 and IgM were detected (Fig. 1A and B; Tables S5-9). Importantly, anti-PEG antibodies were also detected in 14 (5.47%) newborns, with 7 (2.73%) positive for anti-PEG IgG1 and 7 (2.73%) positive for anti-PEG IgG2. No newborn was positive for anti-PEG IgG3, IgG4, IgM or IgE (Fig. 1A and B; Tables S5-9). These data have provided initial characterizations of anti-PEG antibodies in pregnant women (mainly IgM, followed by IgG2 and IgG1) and newborns (IgG1 and IgG2).

**Figure 1.**
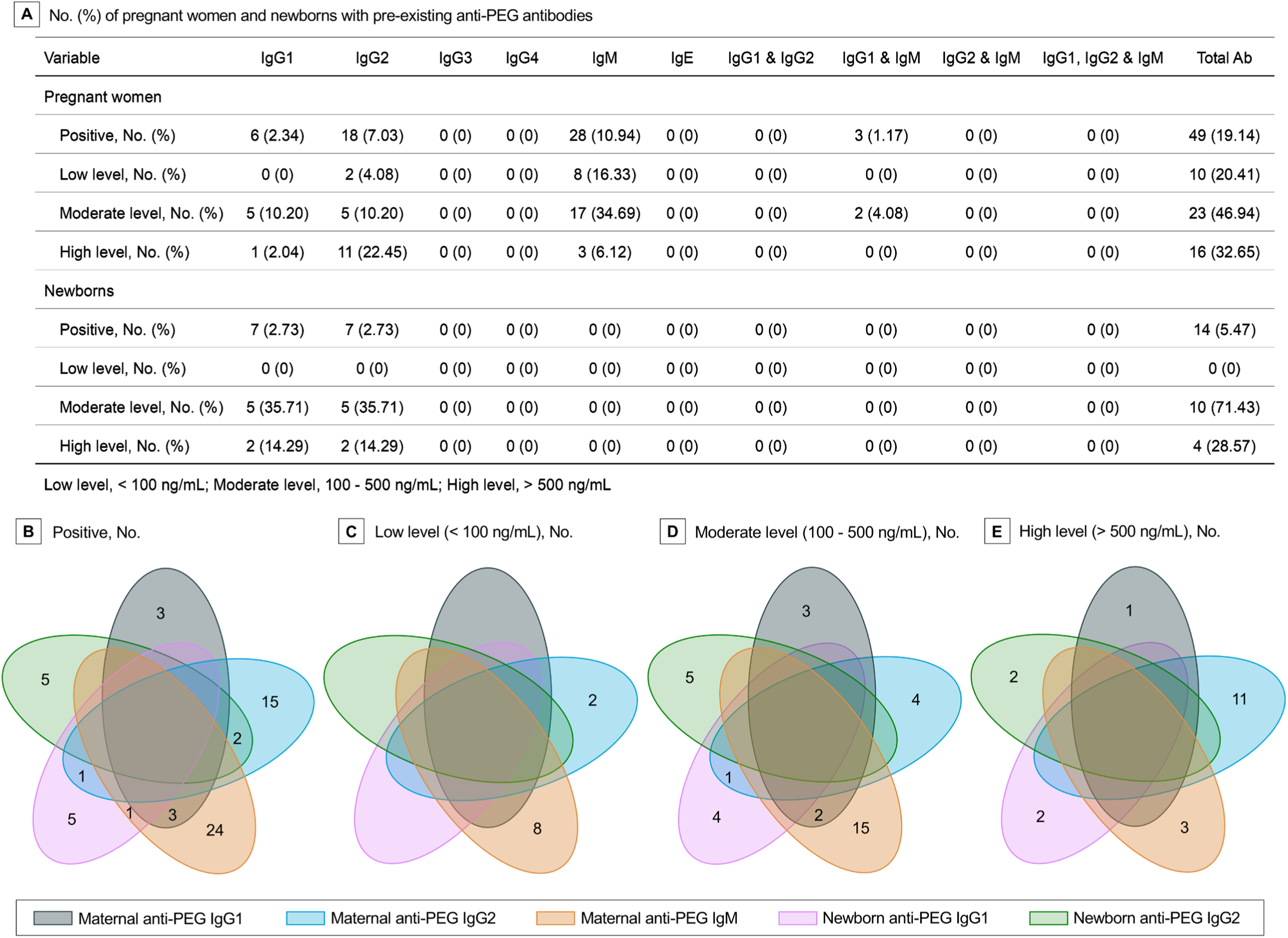
Prevalence and levels of Pre-existing Anti-PEG Antibodies in Pregnant Women and Newborns. Total Ab included all known isotypes/subclasses of anti-PEG antibodies (A). Venn diagrams display the numbers of serum samples with or without coexistence of maternal anti-PEG IgG1, maternal anti-PEG IgG2, maternal anti-PEG IgM, newborn anti-PEG IgG1, and newborn anti-PEG IgG2 (B), as well as with low level (< 100 ng/mL) (C), moderate level (100-500 ng/mL) (D) and high level (> 500 ng/mL) of anti-PEG antibodies (E). When the number of samples is 0, 0 is not displayed in the Venn diagrams.

Herein, anti-PEG antibody levels were defined as low (< 100 ng/mL), moderate (100 - 500 ng/mL) and high (> 500 ng/mL) levels according to the literatures^26, 28^. When regarding anti-PEG antibody seropositive pregnant women as the whole population (100%), the following distributions of anti-PEG antibodies were revealed: 10 (20.41%) with low levels (IgG2, 4.08%; IgM, 16.33%), 23 (46.94%) with moderate levels (IgG1, 10.20%; IgG2, 10.20%; IgM, 34.69%; both IgG1 and IgM, 4.08%) and 16 (32.65%) with high levels (IgG1, 2.04%; IgG2, 22.45%; IgM, 6.12%) (Fig. 1A, C, D and E; Tables S5-9). Correspondingly, the following distributions were found when regarding 14 seropositive newborns as the whole population (100%): no newborn with low levels, 10 (71.43%) with moderate levels (IgG1, 35.71%; IgG2, 35.71%), and 4 (28.57%) with high levels (IgG1, 14.29%; IgG2, 14.29%) of anti-PEG antibodies (Fig. 1A, C, D and E; Tables S5-9). Moreover, seropositive pregnant women had a median (Range; IQR) anti-PEG IgG1, IgG2 and IgM concentrations of 273.88 ng/mL (183.74-513.90; 191.70-376.08), 748.35 ng/mL (75.54-2604.89; 159.09-1200.81) and 175.07 ng/mL (55.43-23649.14; 95.95-315.19), respectively (Fig. 2A). Seropositive newborns had a median (Range; IQR) anti-PEG IgG1 and IgG2 concentrations of 207.92 ng/mL (120.40-1513.98; 133.97-524.58) and 336.52 ng/mL (100.24-1069.62; 275.80-527.07), respectively (Fig. 2B).

**Figure 2.**
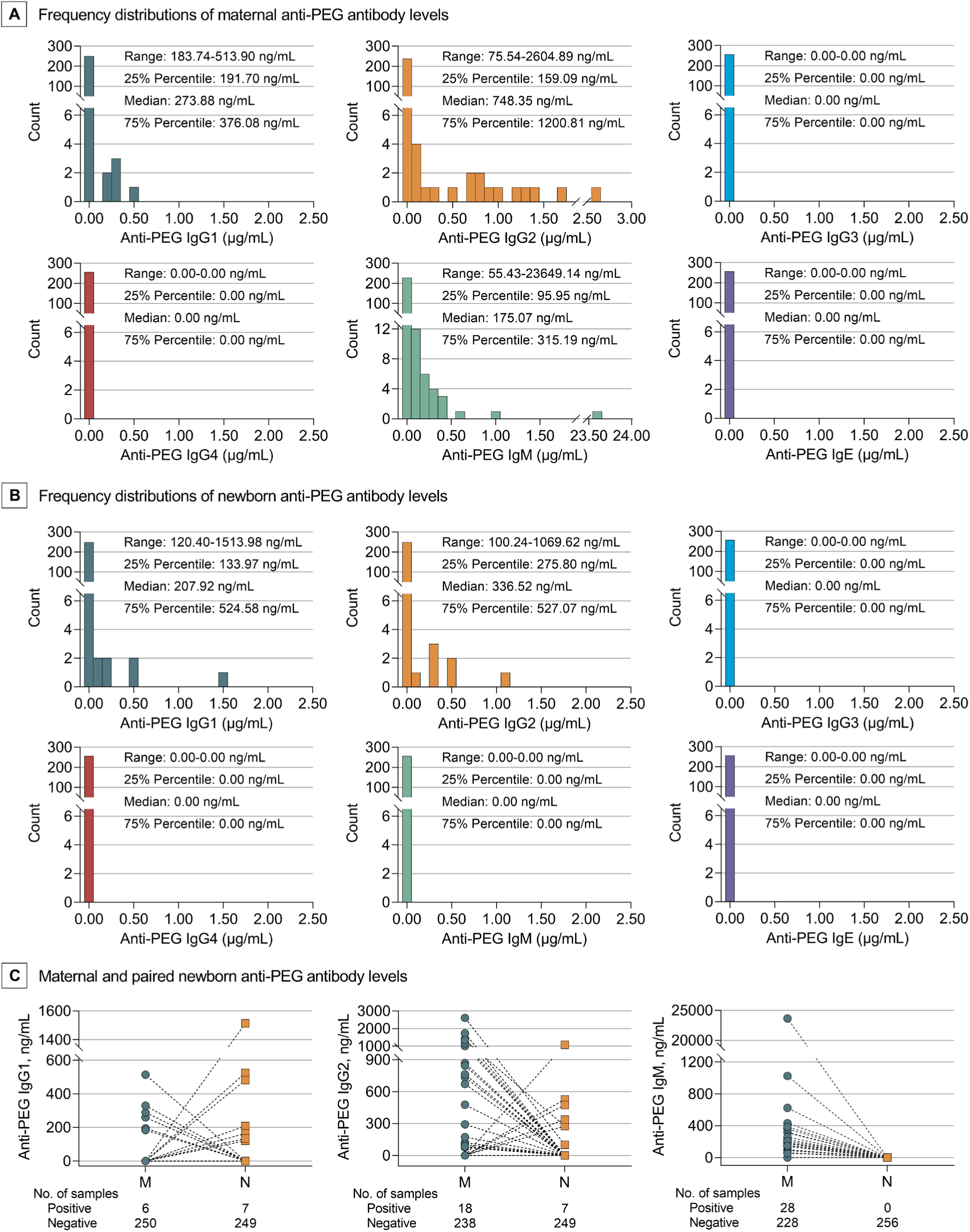
Frequency Distributions of Maternal and Newborn Pre-existing Anti-PEG Antibody Levels in Seropositive Pregnant Women and Newborns. Concentration range, 25% percentile, median and 75% percentile of each antibody isotype/subclass in seropositive pregnant women (A) and newborns (B). Specifically, maternal (M) and paired newborn (N) anti-PEG antibody levels were illustrated (C). Only detected isotypes/subclasses of anti-PEG antibodies were presented.

### Maternal-newborn Differences and Associations of Pre-existing Anti-PEG Antibodies

The prevalence and levels of anti-PEG antibodies were further analyzed to uncover the maternal-newborn differences and associations (Fig. 1B and Fig. 2C; Tables S5-9). Our data showed that anti-PEG IgG2 was detected in 2 pregnant women and their newborns, with higher antibody levels in mothers (1337.37 vs 275.80 ng/mL; 2604.89 vs 100.24 ng/mL). As neither newborns nor fetuses could generate IgG antibodies owing to immature immune systems^33–35^, this data suggest a transfer of anti-PEG IgG2 from mothers to their fetuses. Interestingly, although 7 newborns were positive for anti-PEG IgG1, their mothers were negative for anti-PEG IgG1 despite one mother positive for anti-PEG IgG2 and another positive for anti-PEG IgM. Similarly, although 5 newborns were positive for anti-PEG IgG2, their mothers were undetectable for anti-PEG antibodies. Moreover, 6 pregnant women were positive for anti-PEG IgG1, but their newborns were seronegative for anti-PEG antibodies. Sixteen pregnant women were detected with anti-PEG IgG2, but none of their newborns were positive for anti-PEG IgG2 despite one newborn had anti-PEG IgG1. The complexities of maternal-newborn distribution of anti-PEG IgG not only suggest a transmission of anti-PEG IgG antibodies between mothers and fetuses, but also indicate dynamic changes in antibody subclasses and levels, which may be attributed to varied metabolism and clearance of these antibodies between mothers and fetuses. In addition, 28 pregnant women had anti-PEG IgM, whereas their newborns did not. This data aligns with previous findings that maternal IgM antibodies are unable to cross the placental barrier to reach the fetus^33–35^.

### Multivariable Logistic Regression Analysis for the Prevalence of Pre-existing Maternal Anti-PEG Antibodies

Variables positive in univariable logistic regression analysis (Results S1; Table S10), including maternal age and take-out food consumption, were entered into a multivariable logistic regression analysis to estimate adjusted associations with the prevalence of maternal total anti-PEG antibodies. Our data showed that: with same take-out food consumption, for each 1-year increase in maternal age, the prevalence of maternal total anti-PEG antibodies dropped to 88.3% (95% CI, 81.5-95.8; *P* = 0.003) (Fig. 3A); with same maternal age, compared with those without take-out food consumption, the prevalence of maternal total anti-PEG antibodies increased to 332.7% (95% CI, 125.0-885.4; *P* = 0.016) and 422.4% (95% CI, 123.0-1449.9; *P* = 0.022), respectively, for pregnant women with take-out food 1-3 times and 7-9 times per week (Fig. 3A). These data elucidate that maternal age (inverse association) and take-out food consumption (positive association) are independent influencing factors of the prevalence of maternal total anti-PEG antibodies.

**Figure 3.**
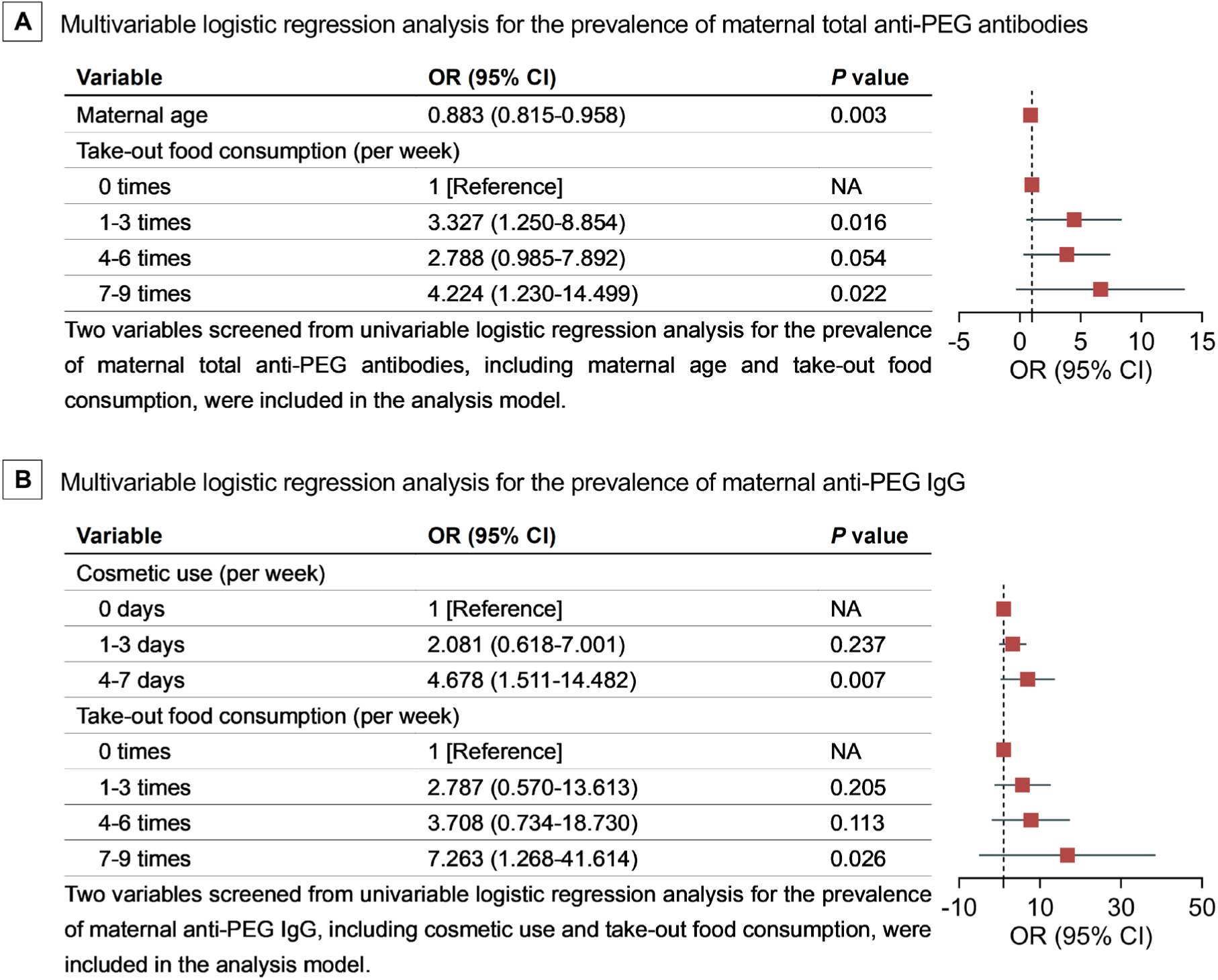
Multivariable Logistic Regression Analysis for the Prevalence of Pre-existing Anti-PEG Antibodies in Pregnant Women. Abbreviations: NA, not applicable; OR, odds ratio. A, maternal total anti-PEG antibodies in pregnant women (n = 256). B, maternal anti-PEG IgG in pregnant women (n = 256). OR for maternal age represents the change in the prevalence of maternal total anti-PEG antibodies for each 1-year increase in the age of pregnant women after adjustment for take-out food consumption; and OR for take-out food consumption represents the change in the prevalence of maternal total anti-PEG antibodies for pregnant women who had take-out food 1-3 times, 4-6 times, and 7-9 times per week compared with those without take-out food consumption after adjustment for maternal age (A). OR for cosmetic use represents the change in the prevalence of maternal anti-PEG IgG for pregnant women who use cosmetics 1-3 days or 4-7 days per week compared with no cosmetic use after adjustment for take-out food consumption; and OR for take-out food consumption represents the change in the prevalence of maternal anti-PEG IgG for pregnant women who had take-out food 1-3 times, 4-6 times and 7-9 times per week compared with those without take-out food consumption after adjustment for cosmetic use (B).

Moreover, variables positive in univariable logistic regression analysis (Results S1; Table S11), including cosmetic use and take-out food consumption, were entered into a multivariable logistic regression analysis to estimate adjusted associations with the prevalence of maternal anti-PEG IgG. Our data indicate that: with same take-out food consumption, compared with no cosmetic use, the prevalence of maternal anti-PEG IgG increased to 467.8% (95% CI, 151.1-1448.2; *P* = 0.007) for pregnant women using cosmetics 4-7 days per week (Fig. 3B); with same cosmetic use, compared with those without take-out food consumption, the prevalence of maternal anti-PEG IgG increased to 726.3% (95% CI, 126.8-4161.4; *P* = 0.026) for pregnant women with take-out food 7-9 times per week (Fig. 3B). These data suggest that maternal cosmetic use (positive association) and take-out food consumption (positive association) are independent influencing factors of the prevalence of maternal anti-PEG IgG.

In addition, univariable logistic regression analysis for the prevalence of maternal anti-PEG IgM only showed that for each 1-year increase in the age of pregnant women, the prevalence of maternal anti-PEG IgM dropped to 88.6% (95% CI, 78.3-95.8; *P* = 0.005) (Results S1; Table S11). That is, requirements for further multivariable logistic regression analysis were not fulfilled for the prevalence of maternal anti-PEG IgM. Similarly, univariable logistic regression analysis for the prevalence of newborn total anti-PEG antibodies only showed that compared with those without cosmetic use, the prevalence of newborn total anti-PEG antibodies increased to 725.9% (95% CI, 215.9-2441.1; *P* = 0.001) for pregnant women with 4-7 days per week cosmetic use (Results S1; Table S12). Hence, multivariable logistic regression analysis was not eligible for the prevalence of newborn total anti-PEG antibodies.

### Multivariable Generalized Linear Regression Analysis for the Levels of Pre-existing Maternal Anti-PEG Antibodies

Variables positive in Spearman correlation analysis (Results S2; Fig. S43) and univariable generalized linear regression analysis (Results S3; Table S13), including maternal age and cosmetic use, were entered into a multivariable generalized linear regression analysis to estimate adjusted associations with the levels of maternal total anti-PEG antibodies after log_10_ transformation among seropositive pregnant women. Our data showed that: with same cosmetic use, for each 1-year increase in the maternal age, the anti-PEG antibody levels increased by 0.047 (95% CI, 0.014-0.079; *P* = 0.005) (Fig. 4A); with same maternal age, compared with no cosmetic use, the anti-PEG antibody levels increased by 0.507 (95% CI, 0.148-0.867; *P* = 0.006) for pregnant women with cosmetic use 4-7 days per week (Fig. 4A). These data revealed that maternal age (positive association) and cosmetic use (positive association) are independent influencing factors of the levels of maternal total anti-PEG antibodies.

**Figure 4.**
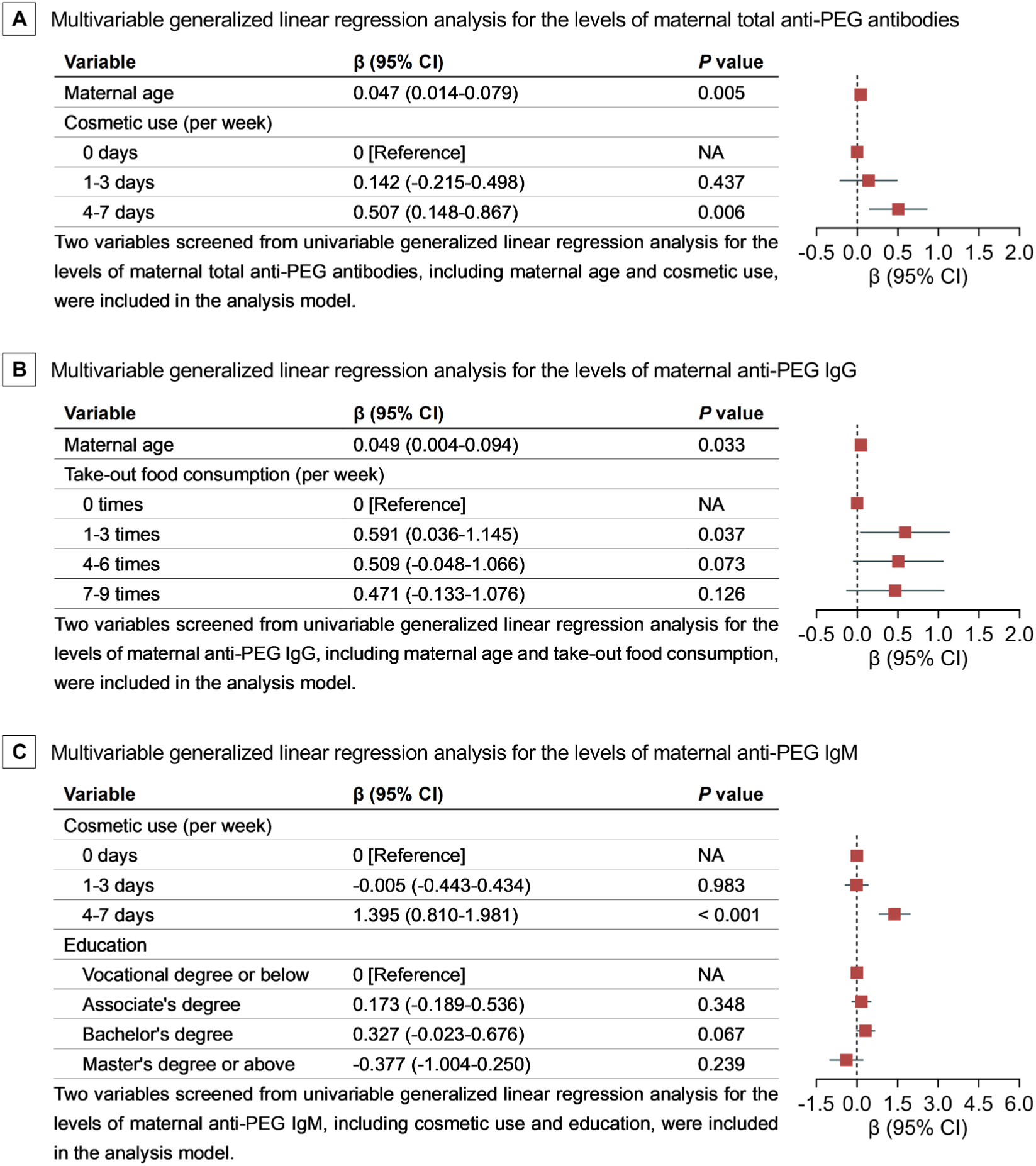
Multivariable Generalized Linear Regression Analysis for the Levels of Pre-existing Anti-PEG Antibodies in Seropositive Pregnant Women. Abbreviations: NA, not applicable; β, standardized regression coefficient. A, total anti-PEG antibodies in seropositive pregnant women (n = 49). B, anti-PEG IgG in seropositive pregnant women (n = 24). C, anti-PEG IgM in seropositive pregnant women (n = 28). β for maternal age represents the change of total anti-PEG antibody level for each 1-year increase in the age after adjustment for cosmetic use; β for cosmetic use represents the change of total anti-PEG antibody level between cosmetic use at indicated frequency and no cosmetic use after adjustment for maternal age (A). β for maternal age represents anti-PEG IgG level for each 1-year increase in the age after adjustment for take-out food consumption; β for take-out food consumption represents the change of anti-PEG IgG level between take-out food consumption at indicated frequency and no take-out food consumption after adjustment for maternal age (B). β for cosmetic use represents the change of anti-PEG IgM level between cosmetic use at indicated frequency and no cosmetic use after adjustment for education; β for education represents the change of anti-PEG IgM level between indicated degree and vocational degree or below after adjustment for cosmetic use (C).

Moreover, variables positive in Spearman correlation analysis (Results S2; Fig. S43) and univariable generalized linear regression analysis (Results S3; Table S14), including maternal age and take-out food consumption, were entered into a multivariable generalized linear regression analysis to estimate adjusted associations with the levels of maternal anti-PEG IgG after log_10_ transformation among anti-PEG IgG seropositive pregnant women. Our data revealed that: with same take-out food consumption, for each 1-year increase in the maternal age, the anti-PEG IgG levels increased by 0.049 (95% CI, 0.004-0.094; *P* = 0.033) (Fig. 4B); with same maternal age, compared with those without take-out food consumption, the anti-PEG IgG levels increased by 0.591 (95% CI, 0.036-1.145; *P* = 0.037) for pregnant women with take-out food 1-3 times per week (Fig. 4B). These data demonstrate that maternal age (positive association) and take-out food consumption (positive association) are independent influencing factors of the levels of maternal anti-PEG IgG. Interestingly, it is noteworthy that no positive associations were detected between anti-PEG IgG levels with more frequent take-out food consumption such as 4-6 times per week and 7-9 times per week. This data may coincide with previous studies indicating that the positive correlation between anti-PEG antibody level and exposure to a specific antigen was only observed within a certain range of PEG antigen dosages^10, 36^.

Furthermore, variables positive in Spearman correlation analysis (Results S2; Fig. S43) and univariable generalized linear regression analysis (Results S3; Table S14), including maternal education and cosmetic use, were entered into a multivariable generalized linear regression analysis to estimate adjusted associations with the levels of maternal anti-PEG IgM after log_10_ transformation among anti-PEG IgM seropositive pregnant women. Our data indicate that: with same education background, compared with no cosmetic use, the anti-PEG IgM levels increased by 1.395 (95% CI, 0.810-1.981; *P* < 0.001) for pregnant women using cosmetics 4-7 days per week (Fig. 4C). However, anti-PEG IgM levels were not affected by education (with same cosmetic use) (Fig. 4C). These data suggest that cosmetic use is an independent influencing factor (positive association) of the levels of maternal anti-PEG IgM, while education is not.

Lastly, Spearman correlation analysis for the levels of newborn total anti-PEG antibodies after log_10_ transformation only showed a positive correlation between the levels of newborn total anti-PEG antibodies and maternal age (*r* = 0.711, *P* = 0.006) (Results S2; Fig. S44), and univariable generalized linear regression analysis for the levels of newborn total anti-PEG antibodies after log_10_ transformation only showed that for each 1-year increase in the age of pregnant women, the levels of newborn total anti-PEG antibodies increased by 0.065 (95% CI, 0.032-0.098; *P* < 0.001) (Results S3; Table S15). Therefore, no further multivariable logistic regression analysis was conducted for the levels of newborn total anti-PEG antibodies.

## DISCUSSION

It is noteworthy that the firstly reported low overall seropositivity of pre-existing anti-PEG antibodies (0.2%) might be attributed to the low sensitivity of detection method^37, 38^. Certainly, it may also reflect the low exposure to PEG antigens in the early 1980s^37, 38^. Afterwards, the detection methods for anti-PEG antibodies were improved, with the combination of direct ELISA with nanotechnology-based competitive ELISA being regarded as most reliable^37, 39^. Using this internally recognized method, herein we revealed that the seropositivities of total anti-PEG antibodies in pregnant women and newborns ware 19.14% and 5.47%, respectively. Further profile analysis on anti-PEG IgG1-G4, IgM and IgE showed that only anti-PEG IgM, IgG1 and IgG2 were detectable in pregnant women, with IgG2 as the dominant IgG subclass (Fig. 1; Fig. S45). Previously there was one literature that determined the anti-PEG IgG subclasses in general adults, in which anti-PEG IgG2 was also most dominant^26^. IgG2 and IgM antibodies are commonly associated with T cell-independent immune responses against non-protein antigens such as lipids, nucleic acids, polysaccharides, and other natural and synthetic polymers^21, 22, 40^. In contrast, protein antigens predominantly induce IgG1 and IgG3 antibodies, with some IgG4 and IgE, by T cell-dependent immune responses^21, 22, 40^. Nevertheless, IgG1 could also arise through a T cell-independent pathway^40^. Therefore, the absence of IgG3, IgG4 and IgE, the presence of IgG1, IgG2, IgM and co-existence of IgG1 and IgM suggest that pre-existing anti-PEG antibodies in pregnant women are more likely generated through T cell-independent mechanisms, supporting the hypothesis that these antibodies stem from exposure to non-protein everyday chemicals containing PEG and its derivatives. Moreover, consistent with five previous studies conducted in general adults^25–29^ (Table S1), anti-PEG IgM and IgG exhibited similar seropositivity in pregnant women.

Moreover, parallel investigation in pregnant women and newborns has provided an opportunity to uncover the potential maternal-fetal/newborn transfer or associations in pre-existing anti-PEG antibodies between two generations (Fig. S45). As fetuses are incapable of synthesizing IgG antibodies^33–35^, the presence of fetal anti-PEG IgG in newborns may suggest active maternal-fetal/newborn antibody transfers across the placenta. Nevertheless, as several factors such as IgG subclasses, maternal antibody titers, gestational age and nature of antigen may affect the placental transfer efficacy of IgG^34, 41^, it is understandable to see discrepancies in prevalence and levels of anti-PEG IgG between pregnant women and newborns. Particularly, mothers and fetuses have disparities in antibody metabolism^42^. On the other hand, different from IgG, maternal IgM antibodies could not cross the placental barrier to achieve maternal-fetal transfer^34, 43–45^. However, fetuses could produce IgM antibodies by themselves in response to T-independent antigens including PEG and PEGylated derivatives^43–45^. Therefore, the absence of anti-PEG IgM antibodies in newborns may suggest lack of uterus PEG antigen exposure and/or degradation of fetal anti-PEG IgM induced by uterus PEG antigen exposure. These findings have deepened our understandings of the induction of pre-existing anti-PEG antibodies and provided initial evidence for their maternal-fetal/newborn transfer.

Previously, several “inherent” factors such as gender and age have been preliminarily evaluated for their possible influences on pre-existing anti-PEG antibodies in general adults^25–30^ (Tables S1 and 2). However, inconsistent findings were obtained across very limited literatures. For instance, both the prevalence and levels of pre-existing anti-PEG IgG exhibited either a negative correlation^25, 26^ or no correlation^28^ with donor age in general adults, while no correlation was reported between either prevalence or levels of anti-PEG IgM and donor age^25–28, 30^. In addition, the highest prevalence of anti-PEG IgG was observed among American adults aged 18-24 years in one report^28^. These findings suggest age-related disparities as well as potential population heterogeneity in pre-existing anti-PEG antibodies. Interestingly, herein maternal age exhibited a negative correlation with the prevalence of total anti-PEG antibodies and anti-PEG IgM (but not IgG), and a positive correlation with the levels of total anti-PEG antibodies and anti-PEG IgG (but not IgM). Meanwhile, we discovered that maternal age had a positive correlation with total newborn anti-PEG antibody (total newborn anti-PEG IgG) levels (Fig. S45). Although further studies on regulatory mechanisms are needed, the specific age range (23∼39 years old) of seropositive pregnant women, distinctive and dynamic immunological milieus during pregnancy and potential maternal-fetal/newborn crosstalk should be taken into account.

As mentioned above, it has been hypothesized that pre-existing anti-PEG antibodies are associated with exposure to everyday chemicals containing PEG and its derivatives^21, 25, 26, 39^. However, this hypothesis has been rarely tested, and till now only two reports suggesting the association between cosmetic use (skin contact) and pre-existing anti-PEG antibodies, one conducted in mice^46^ and another in general adults^47^. Herein, we revealed that the frequency of maternal cosmetic use is a positive influencing factor for the prevalence of maternal anti-PEG IgG, and for the levels of maternal total anti-PEG antibodies and anti-PEG IgM. Interestingly, we discovered that the frequency of take-out food (ingestion) was a positive influencing factor for the prevalence of maternal total anti-PEG antibodies and anti-PEG IgG, and for the levels of maternal anti-PEG IgG, suggesting that anti-PEG antibodies could be induced by gastrointestinal exposure to PEG antigens (Fig. S45).

Most importantly, as a group of anti-drug antibodies (ADA), anti-PEG antibodies have been demonstrated to induce accelerated blood clearance and increase risks of adverse reactions of PEGylated drugs, including hypersensitivity reactions, in a number of animal studies^7, 9, 10, 23, 48–59^ and in several clinical investigations^11, 14–17, 47, 60–62^ (Discussion S1; Tables S16 and 17). Particularly, a positive correlation between the concentration of ADA and changes in pharmacokinetics (PK) has been confirmed in clinical trials, with alterations of PK parameters even induced by certain low level ADA (30-100 ng/mL)^63^. Considering that moderate to high levels of pre-existing anti-PEG antibodies with potential complement-activating activity were observed in a subset of pregnant women and newborns, our findings have naturally raised efficacy and safety concerns over the use of PEGylated drugs in seropositive pregnant women and newborns (Figs. 1 and 2). Indeed, tests for anti-PEG antibodies have been recently recommended by FDA guidelines in patients for assessing the potential immune response to PEGylated therapies^64^. Finally, it is worthy to note that as passively acquired maternal antibodies in newborns wane throughout 6 to 12 months after birth^33^, the anti-PEG antibodies detected in newborns may have long-term immunological effects. This is of particular importance as some PEGylated drugs, including Pfizer and Moderna COVID-19 vaccines, have been officially approved for infants as young as 6 months^65^.

With 256 maternal and 256 newborn blood samples collected around delivery, this cross-sectional study has characterized the prevalence and levels of pre-existing anti-PEG antibodies at a specific time point with representative maternal-newborn populations. In addition, although all the isotypes and subclasses of anti-PEG antibodies previously reported in literatures have been examined in this study, it remains unclear whether there is undiscovered isotype or subclass of anti-PEG antibodies. Moreover, as a reasonable cost of time was needed for questionnaire interviews, we selected a number of factors that we are particularly interested in for data collection and subsequent in-depth statistical analysis. Further evaluations assessing additional factors are needed, which will broaden our understandings on the origins of PEG antigens and induction of risky pre-existing anti-PEG antibodies.

## CONCLUSIONS

Sero-prevalence and levels of pre-existing anti-PEG antibodies were revealed in pregnant women and their newborns with this cross-sectional study, which has raised efficacy and safety concerns over the use of PEGylated drugs in seropositive pregnant women and newborns. Moreover, several influencing factors on maternal and newborn pre-existing anti-PEG antibodies were discovered. These findings have broadened our understandings on seroepidemiological characteristics of pre-existing anti-PEG antibodies, and provided useful clues for identifying the origins of PEG antigens inducing risky anti-PEG antibodies. Specifically, our data may provide initial evidence that gastrointestinal exposure to PEG and its derivatives could induce pre-existing anti-PEG antibodies.

## METHODS

### Study Design and Participants

This cross-sectional study was approved by the Ethics Committee of Women’s Hospital, Zhejiang University School of Medicine (approval No. IRB-20210139-R) and carefully followed the Strengthening the Reporting of Observational Studies in Epidemiology (STROBE) guidelines^66^. Pregnant women admitted for delivery at the Women’s Hospital, Zhejiang University School of Medicine were approached for enrollment with a cross-sectional survey conducted between May 2021 and May 2022. Eligibility criteria included an age of 20 years or older, a singleton pregnancy, a full-term pregnancy (ranging from 37^+0^ to 41^+6^), no history of unhealthy lifestyle (e.g. smoking, drinking or drug abuse), no evidence for exposure to PEGylated drugs before and throughout the pregnancy, no contagious disease, no disease requiring drug treatment upon enrollment and a willingness to participate in this study. Pregnant women participating in other clinical studies were excluded. Eligible participants were identified by dedicated clinicians, with written informed consents obtained from all the participants in accordance with institutional requirements and the Declaration of Helsinki. Maternal blood samples were collected from the peripheral vein within 2 days before delivery. Newborn blood samples were collected from the umbilical cord vein immediately after delivery. All blood samples were centrifuged at 1000 × g for 10 minutes at 4°C, and the serums were immediately harvested and stored at −80°C for further determination of anti-PEG antibodies.

### Questionnaire Interviews and Data Collections

The questionnaire was designed following the ACCADEMY and CHERRIES guides^67, 68^. A face-to-face questionnaire interview was conducted with each participant by a specially trained physician within three days before or after delivery. It is noteworthy that considering the extremely common application of PEG and its derivatives in cosmetics, as well as packaging materials in processed foods and beverages^25, 30^, weekly frequency of cosmetic (e.g. mascara, brow gel, liquid foundation, blemish balm cream, color correcting cream, primer, makeup remover oil, make up removing milk, etc.) use and take-out food consumption during pregnancy were evaluated. In addition, demographic and clinical data including maternal age, body mass index (BMI), education level, permanent address, gestation age at delivery, newborn gender and newborn weight were obtained from hospital electronic medical record (EMR) systems.

### Screening and Quantification of Anti-PEG Antibodies in Serum Samples by ELISA

Anti-PEG antibodies in all serum samples were initially screened using a direct ELISA as described in previous studies^25, 30^. Briefly, Maxisorp^TM^ 96-well microplates were coated with 0.05 mg/well NH_2_-PEG_10000_-NH_2_ in 100 µL of PBS overnight at 4℃. Subsequently, plates were gently washed with 350 μL of DPBS for three times, followed by incubation with blocking buffer (5% (w/v) skim milk powder in DPBS, 200 μL/well) at room temperature (RT) for 1.5 hours. Then the plates were washed three times with DPBS again. Afterwards, 100 μL of each serum sample diluted at 1:10 with sample dilution buffer (2% (w/v) skim milk powder in DPBS), together with six serial dilutions of human anti-PEG IgG1-4, IgM or IgE standards (respectively at 10.3, 30.9, 92.6, 277.8, 833.3 and 2500.0 ng/mL) in standard dilution buffer (10% reference human serum tested negative for anti-PEG antibodies (Methods S1; Figs. S46 and 47), 2% (w/v) skim milk powder in DPBS) were added into corresponding detection plates in duplicate (for serum samples) or triplicate (for anti-PEG antibody standards). Same volumes of sextuplicate standard dilution buffer were used as negative controls. After further incubation for 1 hour at RT, and five successive washes including four with 350 μL of washing buffer (0.05% (w/v) CHAPS in DPBS) and one with 350 μL of DPBS, 50 µL of diluted mouse anti-human secondary antibodies (IgG1 Fc, 1:2500; IgG2 Fc, 1:5000; IgG3 Hinge, 1:5000; IgG4 Fc, 1:500; IgE, 1:10000) and goat anti-human IgM µ-chain secondary antibody (1:10000) were respectively added and incubated for 1 hour at RT. Again, unbounded antibodies were removed by five successive washes, followed by incubation with 100 µL of TMB for 30 minutes at RT in the dark. Finally, the HRP-TMB reaction was stopped with 100 μL of 2 N H_2_SO_4_ and the absorbance was measured at 450 nm. Detailed information on the materials used in ELISA was listed in Methods S2, and details on the establishment of detection cutoffs of anti-PEG IgG1-4, IgM and IgE were introduced in Methods S3. Serum samples with average absorbance values higher than the detection cutoffs in direct ELISA were further verified with a nanotechnology-based competitive ELISA, in order to confirm the PEG specificity of the antibodies detected and minimize false positives (Methods S4). Only serum samples containing PEG-specific antibodies were ultimately deemed positive for anti-PEG antibodies, and further quantified based on the standard curves established for each batch of the direct ELISA. Detailed information on direct ELISA was described in Methods S5.

### Statistical Analysis

Continuous variables (maternal age, BMI, gestation age at delivery and newborn weight) were presented as mean with standard deviation (SD) or median with interquartile range (IQR). Categorical variables (maternal education, permanent address, cosmetic use frequency, take-out food consumption frequency and newborn gender) were presented as numbers with percentages (%). To evaluate the correlations between the prevalence of maternal and newborn anti-PEG antibodies with demographic and clinical variables, univariable logistic regression analysis was firstly performed. Then variables with *P* < .05 in the univariable logistic regression analysis were re-entered into a multivariable logistic regression analysis to evaluate their potential independent associations with the prevalence of anti-PEG antibodies. Furthermore, for pregnant women and newborns seropositive for anti-PEG antibodies, correlations between antibody levels after log_10_ transformation and each continuous variable were analyzed using Spearman correlation coefficients (*r*). Meanwhile, univariable generalized linear regression analysis was conducted to evaluate the correlation of antibody levels after log_10_ transformation with demographic and clinical variables in seropositive pregnant women and newborns. Then variables with *P* < 0.05 in the univariable generalized linear regression analysis were re-entered into a multivariable generalized linear regression analysis to determine their potential independent associations with maternal and newborn anti-PEG antibody levels after log_10_ transformation. Multivariable analysis was only performed in case of 10 or more events per variable (EPV) to avoid bias of the regression coefficients^69^. Statistical analyses were performed using GraphPad Prism version 9.0 and IBM SPSS statistics version 29. All statistical tests were based on 2-tailed hypotheses. Differences were considered significant at *P* < 0.05 unless specifically stated.

## Data Availability

All data produced in the present work are contained in the manuscript

## ASSOCIATED CONTENT

## Supporting Information

The Supporting Information are available free of charge.

Supplementary methods for direct and competitive ELISA; supplementary results from univariable logistic regression analysis, Spearman correlation analysis, and univariable generalized linear regression analysis, focusing on the prevalence or levels of pre-existing anti-PEG antibodies in pregnant women and newborns; supplementary discussion on the clinical implications of anti-PEG antibodies; raw data regarding the detection of anti-PEG antibodies in serum samples by direct ELISA and confirmation of PEG specificity by competitive ELISA; and other additional information.

## AUTHOR INFORMATION

## Author contributions

M.H.S., the principal investigator of the major supporting grants, had full access to all the data in the study and took full responsibility for the integrity of the data and the accuracy of the data analysis. M.H.S., H.Y.W., Y.F. and C.Z.Y. conceived and designed the study. M.H.S. and H.Y.W. drafted the manuscript. All authors revised and edited the manuscript. M.H.S., H.Y.W. and L.Z. analyzed the data. M.H.S., Y.F., L.Z., C.Z.Y., J.C.L. and X.X. provided technical/platform supports. H.Y.W., Y.F. and L.Z. contributed equally as the first authors.

## Notes

The authors declare no competing financial interest.

A version of this manuscript was submitted to the preprint server *medRxiv* on December 10, 2024 as the following: Wang, H.Y.; Feng, Y.; Zhang, L.; Yuan, C.Z.; Xue, J.Y.; Li, J.C.; Xu, X.; Zhou, W.B.; Li, B.H.; Wang, Y.S.; Luo, G.; Zheng, Y.; Sui, M.H.. Pre-existing anti-polyethylene glycol antibodies in pregnant women and newborns. *medRxiv***2024**; 10.1101/2024.11.29.24317450 (accessed December 10, 2024).

## ACKNOWLEDGMENTS

This work was supported by the Zhejiang Provincial Natural Science Foundation of China (LZ23E030003 to Dr. Meihua Sui), National Natural Science Foundation of China (21722405 and 22075243 to Dr. Meihua Sui) and Startup Foundation for Hundred-Talent Program of Zhejiang University (to Dr. Meihua Sui). We would like to acknowledge Dr. Xuesi Chen at Chinese Academy of Sciences for insightful discussions.

