## Supplementary Appendix for "Pre-existing anti-polyethylene glycol antibodies in pregnant women and newborns"

**Content**

**Methods S1.** Screening of Reference Human Serum Negative for Anti-PEG Antibodies
**Methods S2.** Materials and Reagents Used in ELISA
**Methods S3.** Establishment of Detection Cutoffs of Anti-PEG IgG1-4, IgM and IgE Antibodies
**Methods S4.** Confirmation of the PEG Specificity of Antibodies in Serum Samples by Competitive ELISA
**Methods S5.** Direct ELISA for Quantification of Anti-PEG Antibodies
**Results S1.** Univariable Logistic Regression Analysis for the Prevalence of Pre-existing Anti-PEG Antibodies in Pregnant Women and Newborns
**Results S2.** Spearman Correlation Analysis for the Levels of Pre-existing Anti-PEG Antibodies in Seropositive Pregnant Women and Newborns
**Results S3.** Univariable Generalized Linear Regression Analysis for the Levels of Pre-existing Anti-PEG Antibodies in Seropositive Pregnant Women and Newborns
**Discussion S1.** Clinical Implications of Treatment-induced and Pre-existing Anti-PEG Antibodies
**Table S1.** Prevalence of Pre-existing Anti-PEG Antibodies in General Adults across Different Studies
**Table S2.** Levels of Pre-existing Anti-PEG Antibodies in Seropositive General Adults across Different Studies
**Table S3.** Precision of Direct ELISA for Quantification of Maternal and Newborn Anti-PEG Antibodies
**Figure S1.** Detection of Anti-PEG IgG1 in Maternal Serum Samples by Direct ELISA (Batch 1)
**Figure S2.** Detection of Anti-PEG IgG1 in Maternal Serum Samples by Direct ELISA (Batch 2)
**Figure S3.** Detection of Anti-PEG IgG1 in Newborn Serum Samples by Direct ELISA (Batch 1)
**Figure S4.** Detection of Anti-PEG IgG1 in Newborn Serum Samples by Direct ELISA (Batch 2)
**Figure S5.** Detection of Anti-PEG IgG2 in Maternal Serum Samples by Direct ELISA (Batch 1)
**Figure S6.** Detection of Anti-PEG IgG2 in Maternal Serum Samples by Direct ELISA (Batch 2)
**Figure S7.** Detection of Anti-PEG IgG2 in Newborn Serum Samples by Direct ELISA (Batch 1)
**Figure S8.** Detection of Anti-PEG IgG2 in Newborn Serum Samples by Direct ELISA (Batch 2)
**Figure S9.** Detection of Anti-PEG IgG3 in Maternal Serum Samples by Direct ELISA (Batch 1)
**Figure S10.** Detection of Anti-PEG IgG3 in Maternal Serum Samples by Direct ELISA (Batch 2)
**Figure S11.** Detection of Anti-PEG IgG3 in Newborn Serum Samples by Direct ELISA (Batch 1)
**Figure S12.** Detection of Anti-PEG IgG3 in Newborn Serum Samples by Direct ELISA (Batch 2)
**Figure S13.** Detection of Anti-PEG IgG4 in Maternal Serum Samples by Direct ELISA (Batch 1)
**Figure S14.** Detection of Anti-PEG IgG4 in Maternal Serum Samples by Direct ELISA (Batch 2)
**Figure S15.** Detection of Anti-PEG IgG4 in Newborn Serum Samples by Direct ELISA (Batch 1)
**Figure S16.** Detection of Anti-PEG IgG4 in Newborn Serum Samples by Direct ELISA (Batch 2)
**Figure S17.** Detection of Anti-PEG IgM in Maternal Serum Samples by Direct ELISA (Batch 1)
**Figure S18.** Detection of Anti-PEG IgM in Maternal Serum Samples by Direct ELISA (Batch 2)
**Figure S19.** Detection of Anti-PEG IgM in Newborn Serum Samples by Direct ELISA (Batch 1)
**Figure S20.** Detection of Anti-PEG IgM in Newborn Serum Samples by Direct ELISA (Batch 2)
**Figure S21.** Detection of Anti-PEG IgE in Maternal Serum Samples by Direct ELISA (Batch 1)
**Figure S22.** Detection of Anti-PEG IgE in Maternal Serum Samples by Direct ELISA (Batch 2)
**Figure S23.** Detection of Anti-PEG IgE in Newborn Serum Samples by Direct ELISA (Batch 1)
**Figure S24.** Detection of Anti-PEG IgE in Newborn Serum Samples by Direct ELISA (Batch 2)
**Table S4.** Precision of Competitive ELISA for Confirmation of the PEG Specificity of Maternal and Newborn Anti-PEG IgG1-4 and IgM
**Figure S25.** Establishment of Specificity Cutoffs of Maternal Anti-PEG IgG1-4 and IgM
**Figure S26.** Establishment of Specificity Cutoffs of Newborn Anti-PEG IgG1-4 and IgM
**Figure S27.** Confirmation of the PEG Specificity of Anti-PEG IgG1 in Maternal Serum Samples by Competitive ELISA
**Figure S28.** Confirmation of the PEG Specificity of Anti-PEG IgG1 in Newborn Serum Samples by Competitive ELISA
**Figure S29.** Confirmation of the PEG Specificity of Anti-PEG IgG2 in Maternal Serum Samples by Competitive ELISA
**Figure S30.** Confirmation of the PEG Specificity of Anti-PEG IgG2 in Newborn Serum Samples by Competitive ELISA
**Figure S31.** Confirmation of the PEG Specificity of Anti-PEG IgG3 in Maternal Serum Samples by Competitive ELISA
**Figure S32.** Confirmation of the PEG Specificity of Anti-PEG IgG3 in Newborn Serum Samples by

Competitive ELISA
**Figure S33.** Confirmation of the PEG Specificity of Anti-PEG IgG4 in Maternal Serum Samples by Competitive ELISA
**Figure S34.** Confirmation of the PEG Specificity of Anti-PEG IgG4 in Newborn Serum Samples by Competitive ELISA
**Figure S35.** Confirmation of the PEG Specificity of Anti-PEG IgM in Maternal Serum Samples by Competitive ELISA
**Figure S36.** Confirmation of the PEG Specificity of Anti-PEG IgM in Newborn Serum Samples by Competitive ELISA
**Figure S37.** Standard Curves of Direct ELISA for Detecting Anti-PEG IgG1 in Maternal and Newborn Serum Samples
**Figure S38.** Standard Curves of Direct ELISA for Detecting Anti-PEG IgG2 in Maternal and Newborn Serum Samples
**Figure S39.** Standard Curves of Direct ELISA for Detecting Anti-PEG IgG3 in Maternal and Newborn Serum Samples
**Figure S40.** Standard Curves of Direct ELISA for Detecting Anti-PEG IgG4 in Maternal and Newborn Serum Samples
**Figure S41.** Standard Curves of Direct ELISA for Detecting Anti-PEG IgM in Maternal and Newborn Serum Samples
**Figure S42.** Standard Curves of Direct ELISA for Detecting Anti-PEG IgE in Maternal and Newborn Serum Samples
**Table S5.** Anti-PEG IgG1 Levels in Seropositive Pregnant Women
**Table S6.** Anti-PEG IgG1 Levels in Seropositive Newborns
**Table S7.** Anti-PEG IgG2 Levels in Seropositive Pregnant Women
**Table S8.** Anti-PEG IgG2 Levels in Seropositive Newborns
**Table S9.** Anti-PEG IgM Levels in Seropositive Pregnant Women
**Table S10.** Univariable Logistic Regression Analysis for the Prevalence of Pre-existing Total Anti-PEG Antibodies in Pregnant Women (n = 256)
**Table S11.** Univariable Logistic Regression Analysis for the Prevalence of Pre-existing Anti-PEG IgG and IgM in Pregnant Women (n = 256)
**Table S12.** Univariable Logistic Regression Analysis for the Prevalence of Pre-existing Total Anti-PEG Antibodies in Newborns (n = 256)
**Figure S43.** Correlation between the Levels of Pre-existing Total Anti-PEG Antibodies, Anti-PEG IgG and IgM with Maternal Age and BMI in Pregnant Women Seropositive for Anti-PEG Antibodies (n = 49), Anti-PEG IgG (n = 24) and IgM (n = 28)
**Table S13.** Univariable Generalized Linear Regression Analysis for the Levels of Pre-existing Total Anti-PEG Antibodies in Pregnant Women Seropositive for Anti-PEG Antibodies (n = 49)
**Table S14.** Univariable Generalized Linear Regression Analysis for the Levels of Pre-existing Anti-PEG IgG and IgM in Pregnant Women Seropositive for Anti-PEG IgG (n = 24) and IgM (n = 28)
**Figure S44.** Correlation Between the Levels of Pre-existing Total Anti-PEG Antibodies and Maternal Age, Maternal BMI, Gestational Age at Delivery, and Newborn Weight in Newborns Seropositive for Anti-PEG Antibodies (n = 14)
**Table S15.** Univariable Generalized Linear Regression Analysis for the Levels of Pre-existing Total Anti-PEG Antibodies in Newborns Seropositive for Anti-PEG Antibodies (n = 14)
**Figure S45.** Overview Diagram Describing the Design, Procedures and Major Research Findings of this Study
**Table S16.** Representative Preclinical Studies Indicating Accelerated Blood Clearance Phenomenon of PEGylated Drugs
**Table S17.** Clinical Trials Indicating Accelerated Blood Clearance Phenomenon of PEGylated Drugs
**Figure S46.** Screening of Paired Maternal and Newborn Serum Samples Negative for Anti-PEG IgG and IgM
**Figure S47.** Confirmation of the Absence of Anti-PEG IgE in Maternal and Newborn Serum Samples Negative for Anti-PEG IgG and IgM
**References**

#### Methods S1. Screening of Reference Human Serum Negative for Anti-PEG Antibodies

When quantitatively detecting anti-PEG antibodies in serum using direct ELISA, the presence of matrix in the serum can interfere with the detection of anti-PEG antibodies, impacting the accuracy of the results, known as matrix effects[1]. To mitigate these effects and ensure the precision of our assays, we used a standard dilution buffer (10% reference human serum negative for anti-PEG antibodies, 2% (w/v) skim milk powder in DPBS) to create serial dilutions of human anti-PEG antibody standards (see Methods). To prepare the standard dilution buffer, commercial human anti-PEG IgG ELISA kits and human anti-PEG IgM ELISA kits were used to screen a batch of paired maternal and newborn serum samples that were negative for anti-PEG IgG and IgM according to the manufacturer's instructions.

Lacking commercial kits for detecting anti-PEG IgE, we intend to employ competitive ELISA (see below eMethods 4 for detailed experimental procedures) to confirm the presence of anti-PEG IgE in sera negative for anti-PEG IgG and IgM measured by commercial kits. In this experiment, anti-PEG IgG and IgM-negative maternal and newborn pooled serum samples diluted 1:5 with dilution buffer (2% (w/v) skim milk powder in DPBS) were used as the tested samples. Six serial dilutions of human anti-PEG IgE standards (respectively at 8.2, 24.6, 74.0, 222.2, 666.6 and 2000.0 ng/mL) in standard dilution buffer (20% maternal or newborn pooled serum tested negative for anti-PEG IgG and IgM, 2% (w/v) skim milk powder in DPBS) were used as the positive control.

#### Methods S2. Materials and Reagents Used in ELISA

$NH_2$-$PEG_{10000}$-$NH_2$ (Cat. No. 8218815000) was obtained from Sigma-Aldrich (St. Louis, MO, USA). 3-[(3-cholamidopropyl)dimethylammonio]-1-propanesulfonate (CHAPS, Cat. No. ST1145), 3,3′,5,5′-tetramethylbenzidine dihydrochloride hydrate (TMB 2HCl, Cat. No. ST1708) and nonfat powdered milk (Cat. No. P0216) were purchased from Beyotime Biotechnology (Shanghai, China). Maxisorp 96-well microplates (Cat. No. 44-2404-21) were acquired from Nalge-Nunc International (Rochester, NY, USA). Chimeric human anti-PEG IgG1 (Cat. No. cHu-3.3-IgG1, Clone No. c3.3-IgG1), chimeric human anti-PEG IgG2 (Cat. No. cHu-r33G-IgG2, Clone No. cr33G-IgG2), chimeric human anti-PEG IgG3 (Cat. No. cHu-r33G-IgG3, Clone No. cr33G-IgG3), chimeric human anti-PEG IgG4 (Cat. No. cHu-3.3-IgG4, Clone No. c3.3-IgG4), chimeric human anti-PEG IgM (Cat. No. cHu-AGP4-PABG-A, Clone No. cAGP4-IgM) and human anti-PEG IgE (Cat. No. Hu-6.3-IgE, Clone No. Hu 6.3-IgE) were obtained from Academia Sinica (Taipei, China). Mouse anti-human IgG1 Fc secondary antibody (Cat. No. 9054-05, Clone No. HP6001), mouse anti-human IgG2 Fc secondary antibody (Cat. No. 9060-05, Clone No. 31-7-4), and mouse anti-human IgG3 Hinge secondary antibody (Cat. No. 9210-05, Clone No. HP6050) were purchased from Southern Biotech (Birmingham, AL, USA). Goat anti-human IgM μ-chain secondary antibody (Cat. No. 609-1307) was acquired from Rockland Immunochemicals (Philadelphia, Pennsylvania, USA). Mouse anti-human IgE secondary antibody (Cat. No. SA5-10306, Clone No. 24A) and mouse anti-human IgG4 Fc secondary antibody (Cat. No. A-10654, Clone No. HP6025) were obtained from ThermoFisher Scientific (Waltham, MA, USA). Human anti-PEG IgM ELISA kit (Cat. No. PEG-020), and human anti-PEG IgG ELISA kit (Cat. No. PEG-010) were purchased from Alpha Diagnostic Intl (San Antonio, Texas, USA). LIBOD® (PEGylated liposomal doxorubicin) was acquired from Shanghai Fudan-zhangjiang Bio-Pharmaceutical Co. Ltd (Shanghai, China).

#### Methods S3. Establishment of Detection Cutoffs of Anti-PEG IgG1-4, IgM and IgE Antibodies

Detection cutoffs of anti-PEG IgG1-4, IgM and IgE antibodies were established based on the absorbance value of six negative controls (standard dilution buffer: 10% human reference serum negative for anti-PEG antibodies, 2% (w/v) skim milk powder in DPBS) in each batch of the direct ELISA according to the method described by Frey et al[2]. The formula for Detection cutoff is given below: Detection cutoff = $\bar{X} + \mathrm{SD}f$, where $\bar{X}$ is the mean absorbance value of six independent negative controls, SD is the standard deviation of absorbance value of six independent negative controls, $f$ is the standard deviation multipliers based on the number of negative controls (n = 6) and the 95% confidence level.

#### Methods S4. Confirmation of the PEG Specificity of Antibodies in Serum Samples by Competitive ELISA

Serum samples with average absorbance values higher than the corresponding detection cutoffs in the direct ELISA were further verified by a competitive ELISA to confirm the PEG specificity of the antibodies detected. The competitive ELISA experiment was performed based on a previously published procedure from Chen et al[1]. Maxisorp 96-well microplates were prepared following the same procedure as direct ELISA (see Methods). After incubation with blocking buffer (5% (w/v) skim milk powder in DPBS, 200 μL/well) at RT for 5.5 hours, plates were gently washed three times with 350 μL of DPBS. Dilution buffer with competitors was prepared by dissolving clinically relevant PEGylated liposomal doxorubicin (the PEGylated liposomal doxorubicin (LIBOD®), congruent with the formulation of Doxil®, exhibits stable physicochemical characteristics; Given the robust binding affinity of pre-existing anti-PEG antibodies to clinically relevant PEGylated liposomal doxorubicin[1], we have elected this formulation as a competitor in our study) into dilution buffer (2% (w/v) skim milk powder in DPBS) at a concentration of 400 mg phospholipids/mL. Then 50 μL of dilution buffer with competitors or dilution buffer without competitors were added into plates and further incubated for 15 minutes at RT. Following this, 50 μL of anti-PEG antibody-positive serum samples diluted 1:25 with dilution buffer, together with eight serial dilutions of human anti-PEG IgG1-4 and IgM standards (respectively at 2.3, 6.9, 20.6, 61.7, 185.2, 555.6, 1666.7 and 5000.0 ng/mL) in standard dilution buffer (4% human reference serum tested negative for anti-PEG antibodies, 2% (w/v) skim milk powder in DPBS), were added to the wells containing PEGylated liposomes (competitors) or no PEGylated liposomes of corresponding detection plates in duplicate (for serum samples) or triplicate (for anti-PEG antibody standards). Same volumes of triplicate standard dilution buffer were used as negative controls. After further incubation for 1 hour at RT, the same procedure was followed as direct ELISA (see Methods).

Specificity cutoffs of anti-PEG IgG1-4 and IgM antibodies were established based on the absorbance value of anti-PEG antibody standards with the addition of competitors and without the addition of competitors according to the method described by Chen et al[1]. Specifically, the percentage of antibody binding for anti-PEG antibody standards with the addition of competitors and without the addition of competitors was calculated separately by dividing the absorbance value of anti-PEG antibody standards with the addition of competitors and without the addition of competitors by the average absorbance value of those without the addition of competitors. Specificity cutoffs of anti-PEG antibodies were defined as the maximum percentage of antibody binding for anti-PEG antibody standards with the addition of competitors (NOTE: The percentage of antibody binding for anti-PEG antibody standards with the addition of competitors must be significantly less than the percentage of antibody binding for those without the addition of competitors). The percentage of antibody binding for serum samples with the addition of competitors and without the addition of competitors was calculated using the same method as that of antibody binding for anti-PEG antibody standards. Antibodies detected in serum samples via direct ELISA are considered anti-PEG antibodies (antibodies with PEG specificity) if the percentage of antibody binding for serum samples with the addition of competitors was significantly less than that of antibody binding for serum samples without the addition of competitors and no more than the specificity cutoffs of corresponding anti-PEG antibodies.

#### Methods S5. Direct ELISA for Quantification of Anti-PEG Antibodies

Anti-PEG IgG1-4, IgM and IgE standard curves in each batch of the direct ELISA were constructed by plotting the average absorbance values (OD450 nm) and corresponding antibody concentrations with Four Parameter Logistic (4PL) curve fit using Origin 2021 software (OriginLab Corporation, Northampton, Massachusetts, USA). The goodness of fit of each standard curve was evaluated by the coefficient of determination ($R^2$). To evaluate the quality control of ELISA, intra-assay precision was determined by calculating the Coefficient of Variation (CV% = (Standard deviation/Mean) × 100%) for all detectable standards and samples in all batches of ELISA[3]. In addition, inter-assay precision was determined by calculating the Coefficient of Variation for serially diluted anti-PEG antibody standards among all batches of ELISA[3]. The acceptance criteria for ELISA's mean intra-assay and inter-assay coefficient of variation (CV%) are < 20% and < 25%, respectively[3].

#### Results S1. Univariable Logistic Regression Analysis for the Prevalence of Pre-existing Anti-PEG Antibodies in Pregnant Women and Newborns

To reveal the demographic and clinical factors associated with the prevalence of maternal and newborn anti-PEG antibodies, univariable logistic regression analysis was firstly conducted. Interestingly, our data showed that for each 1-year increase in the age of pregnant women, the prevalence of maternal total anti-PEG antibodies and anti-PEG IgM respectively dropped to 88.7% (95% CI, 82.0-95.9; *P* = 0.003; eTable 10) and 88.6% (95% CI, 78.3-95.8; *P* = 0.005; eTable 11). Compared with those without cosmetic use, the prevalence of maternal anti-PEG IgG and newborn total anti-PEG antibodies respectively increased to 584.1% (95% CI, 194.7-1755.8; *P* = 0.002; eTable 11) and 725.9% (95% CI, 215.9-2441.1; *P* = 0.001; eTable 12) for pregnant women with 4-7 days per week cosmetic use. Moreover, compared with those without take-out food consumption, the prevalence of maternal total anti-PEG antibodies increased to 312.9% (95% CI, 119.5-819.1; *P* = 0.020), 298.7% (95% CI, 107.1-832.8; *P* = 0.036) and 414.8% (95% CI, 123.8-1390.1; *P* = 0.021), respectively, for pregnant women who had take-out food 1-3 times, 4-6 times and 7-9 times per week (eTable 10). Meanwhile, compared with those without take-out food consumption, the prevalence of maternal anti-PEG IgG increased to 850.0% (95% CI, 153.1-4718.2; *P* = 0.014) for pregnant women who had take-out food 7-9 times per week (eTable 11). These data revealed inverse associations of the prevalence of maternal total anti-PEG antibodies and anti-PEG IgM with maternal age, while positive associations of the prevalence of maternal total anti-PEG antibodies and anti-PEG IgG with take-out food consumption, as well as a positive association of the prevalence of maternal anti-PEG IgG and newborn total anti-PEG antibodies with cosmetic use. Nevertheless, no other association was found between the prevalence of maternal or newborn anti-PEG antibodies (either total or any isotype) and all other demographic or clinical factors (eTables 10-12). Variables positive in univariable logistic regression analysis, including maternal age and take-out food consumption, were entered into a multivariable logistic regression analysis to estimate adjusted associations with the prevalence of maternal total anti-PEG antibodies. Moreover, variables positive in univariable logistic regression analysis, including cosmetic use and take-out food consumption, were entered into a multivariable logistic regression analysis to estimate adjusted associations with the prevalence of maternal anti-PEG IgG.

#### Results S2. Spearman Correlation Analysis for the Levels of Pre-existing Anti-PEG Antibodies in Seropositive Pregnant Women and Newborns

For pregnant women and newborns seropositive for anti-PEG antibodies, the Spearman correlation analysis was conducted to evaluate the correlation between the levels of maternal and newborn anti-PEG antibodies after $log_{10}$ transformation and each continuous variable. Correlation plots showed a moderate positive correlation between the levels of maternal total anti-PEG antibodies and maternal age ($r$ = 0.363, $P$ = 0.01; eFigure 43A), as well as a higher positive correlation between the levels of newborn total anti-PEG antibodies and maternal age ($r$ = 0.711, $P$ = 0.006; eFigure 44A). Nevertheless, no correlation was found between the levels of maternal total anti-PEG antibodies and maternal BMI (eFigure 43B), as well as between the levels of maternal anti-PEG IgG and IgM with both maternal age and BMI (eFigure 45C-F). Nor did we find any correlation between the levels of newborn total anti-PEG antibodies and maternal BMI, gestational age at delivery and newborn weight (eFigure 44B-D).

#### Results S3. Univariable Generalized Linear Regression Analysis for the Levels of Pre-existing Anti-PEG Antibodies in Seropositive Pregnant Women and Newborns

For pregnant women and newborns seropositive for anti-PEG antibodies, to reveal the demographic and clinical factors associated with the levels of maternal and newborn anti-PEG antibodies after $log_{10}$ transformation, univariable generalized linear regression analysis was firstly conducted. Univariable analysis showed that for each 1-year increase in the age of pregnant women, the levels of maternal total anti-PEG antibodies, maternal anti-PEG IgG, and newborn total anti-PEG antibodies increased by 0.053 (95% CI, 0.018-0.088; *P* = 0.003; eTable 13), 0.057 (95% CI, 0.010-0.104; *P* = 0.018; eTable 14) and 0.065 (95% CI, 0.032-0.098; $P$ < 0.001; eTable 15), respectively. Compared with those with vocational degree or below, the levels of maternal anti-PEG IgM increased by 0.453 (95% CI, 0.005-0.900; *P* = 0.047; eTable 14) for pregnant women with bachelor's degree. Compared with no cosmetic use, the levels of maternal total anti-PEG antibodies and anti-PEG IgM increased by 0.576 (95% CI, 0.191-0.960; *P* = 0.003; eTable 14) and 1.220 (95% CI, 0.645-1.795; $P$ < 0.001; eTable 14), respectively, for pregnant women with cosmetic use 4-7 days per week. Moreover, compared with those without take-out food consumption, the levels of maternal anti-PEG IgG increased by 0.702 (95% CI, 0.107-1.297; *P* = 0.021) and 0.642 (95% CI, 0.006-1.279; *P* = 0.048), respectively, for pregnant women who had take-out food 1-3 times and 7-9 times per week (eTable 14). These data demonstrate positive associations between the levels of maternal total anti-PEG antibodies, anti-PEG IgG and newborn total anti-PEG antibodies with maternal age, a positive association between the levels of maternal anti-PEG IgG and take-out food consumption, as well as a positive association between the levels of maternal total anti-PEG antibodies and anti-PEG IgM with cosmetic use. Nevertheless, no other association was found between the levels of maternal or newborn anti-PEG antibodies (either total or any isotype) and all other demographic or clinical factors (eTables 13-15). Variables positive in univariable generalized linear regression analysis, including maternal age and cosmetic use, were entered into a multivariable generalized linear regression analysis to estimate adjusted associations with the levels of maternal total anti-PEG antibodies after $log_{10}$ transformation among seropositive pregnant women. Moreover, variables positive in univariable generalized linear regression analysis, including maternal age and take-out food consumption, were entered into a multivariable generalized linear regression analysis to estimate adjusted associations with the levels of maternal anti-PEG IgG after $log_{10}$ transformation among anti-PEG IgG seropositive pregnant women. Lastly, variables positive in univariable generalized linear regression analysis, including maternal education and cosmetic use, were entered into a multivariable generalized linear regression analysis to estimate adjusted associations with the levels of maternal anti-PEG IgM after $log_{10}$ transformation among anti-PEG IgM seropositive pregnant women.

#### Discussion S1. Clinical Implications of Treatment-induced and Pre-existing Anti-PEG Antibodies

To date, researchers have confirmed that in rats[4-8], mice[6,7,9,10], rabbits[7], beagle dogs[6] and guinea pigs[7], anti-PEG antibodies induced by previous intravenous injection of PEGylated liposomes led to accelerated blood clearance (ABC) phenomenon in subsequent or repeated injections of same liposomes (eTable 16). Moreover, this anti-PEG antibody-mediated ABC phenomenon were also observed for intravenously injected PEGylated proteins[11-13], PEGylated polymer nanoparticles[14], PEGylated micelles[15], PEGylated nanoemulsions[16], PEGylated exosomes[17] and PEGylated lipid nanoparticles in animal models (eTable 16). Interestingly, ABC phenomenon is not confined to intravenous administration of PEGylated agents. For instance, Elsadek et al. reported that ABC phenomenon occurred following repeated subcutaneous injections of Peginterferon Alfa-2a (Pegasys®)[12] in mice. Most recently, we demonstrated the presence of accelerated blood clearance of the PEGylated lipid nanoparticles of mRNA vaccine Comirnaty® following repeated intramuscular injections in rats[3].

Importantly, several clinical studies have documented the occurrence of the ABC phenomenon in patients following treatment with FDA-approved PEGylated therapeutics[18-23] (eTable 17). For instance, Armstrong et al. discovered that Oncaspar® (PEGylated asparaginase) induced anti-PEG antibodies in pediatric patients with acute lymphoblastic leukemia (ALL), leading to ABC phenomenon and reduced drug efficacy[18]. Impressively, in several clinical studies, both subcutaneous and intravenous injections of Krystexxa® (PEGylated uricase) induced the production of anti-PEG antibodies in gout patients, which resulted in significantly increased drug clearance rates and reduced therapeutic effecticacy[19-22]. Moreover, in a Phase II clinical trial, Zori et al. found that after intravenous infusion of Palynziq® (PEGylated phenylalanine ammonia lyase), approximately 90% of phenylketonuria patients were seropositive for anti-PEG IgG or anti-PEG IgM, which was correlated with augmented drug clearance rates[23]. It is noteworthy that similar to treatment-induced anti-PEG antibodies, several clinical investigations have delineated pre-existing anti-PEG antibody-mediated alterations in pharmacokinetic profiles of PEGylated therapeutics[21,24]. For example, in pediatric subjects undergoing treatment with Oncaspar® for ALL, the presence of pre-existing anti-PEG antibodies was associated with an enhanced systemic clearance of the initially administered drug and a subsequently diminished therapeutic efficacy[24]. Analogously, in individuals seropositive for pre-existing anti-PEG antibodies, there were pronounced reductions in both the plasma concentration and therapeutic efficacy of Krystexxa®[21].

On the other hand, Kozma et al. revealed that anti-PEG IgM could activate the complement system via the classical pathway, causing severe hypersensitivity reactions to PEGylated liposomes in pigs[25]. Later, in mice models, Stavnsbjerg et al. identified a correlation between acute hypersensitivity reactions and production of anti-PEG IgG following repeated intravenous administrations of PEGylated liposomes containing Toll-like receptor (TLR7/8) agonists[26], while Chen et al. found that anti-PEG IgG could activate innate immune cells and induce hypersensitivity reactions to PEGylated liposomes[27]. Notably, Kozma and colleagues linked higher rates of allergic reactions in women to the presence of pre-existing anti-PEG antibodies[28]. In addition, pre-existing anti-PEG antibodies were correlated with mild to moderate hypersensitivity reactions in pediatric patients with ALL following the initial administration of Oncaspar®[24]. Indeed, pre-existing anti-PEG antibodies were associated with severe first-exposure hypersensitivity reactions to PEGylated RNA aptamer (Pegnivacogin), which led to the discontinuation of phase III clinical trials[29].

#### Table S1. Prevalence of pre-existing anti-PEG antibodies in general adults across different studies

| Year[Ref] | Population location | Sample number | Gender (female/male) | Age, range, (y) | Prevalence of anti-PEG antibodies | | | | | Correlations between the prevalence of anti-PEG antibodies and demographic characteristics | Method |
|---|---|---|---|---|---|---|---|---|---|---|---|
| | | | | | Total[a] | IgM | IgG | IgM & IgG | IgE | | |
| 1984[30] | Japan<br>GER<br>Italy | Japan: 142<br>GER: 151<br>Italy: 160 | NA | NA | 0.2% | NA | NA | NA | NA | NA | Passive hemagglutination |
| 2016[31] | Austria | 710 | NA | 18-67 | 23% | 13% | 15% | 5% | NA | IgG prevalence was negatively correlated with donor age; IgM prevalence showed no correlation with donor age. | Flow cytometry with validation assays |
| | Austria USA | Austria:100<br>USA: 500 | NA | 18-67 | 24% | 14% | 12% | 2% | NA | IgG prevalence was negatively correlated with donor age, and was correlated with donor location; IgM prevalence showed no correlation with either donor age or location. | Direct ELISA & competitive ELISA |
| 2016[2] | Taiwan, China | 1504 | 748/756 | 20-80 | 44.3% | 27.1% | 25.7% | 8.4% | NA | Females have a higher prevalence of IgM (32.0% vs 22.2%, $p < 0.0001$) and IgG (28.3% vs 23.0%, $p = 0.018$) than males; IgG prevalence showed negative correlations with donor age. | Direct ELISA & competitive ELISA |
| 2016[32] | USA | 377 | 151/226 | 19-70 | 72% | 55% | 48% (IgG1：26%；IgG2：57%；IgG3：1%；IgG4: 1%) | 30% | NA | IgG prevalence was negatively correlated with donor age; IgM prevalence showed no correlation with donor age; no correlation was observed between donor gender or race and the prevalence of either IgM or IgG. | Competitive ELISA with validation assays |
| | USA | 1970s: 30<br>1980s: 30<br>1990s: 19 | 1970s: 15/15<br>1980s: 15/15<br>1990s: 6/13 | 19-70 | 56% | 35% | 36% | 16% | NA | NA | Competitive ELISA with validation assays |
| 2021[33] | USA | 300 | 116/184 | 18-65 | 65.3% | 44% | 46.3% | 25% | NA | No correlation between donor age, gender or race and the prevalence of either IgM or IgG. | Flow cytometry with validation assays |
| 2023[34] | Germany | 500 | NA | 18-60 | 83% | NA | NA | 48.8% | NA | IgM and IgG prevalence was negatively correlated with donor age. | Direct ELISA & competitive ELISA |
| 2024[35] | USA | 12 | NA | NA | NA | 83% | 83% | NA | 0% | NA | Competitive ELISA |
| 2024 (Present study) | Zhejiang, China | Pregnant woman: 256 | NA | 23-44 | 19.14% | 10.94% | 9.38% (IgG1：2.34%；IgG2：7.03%；IgG3 & IgG4：0%) | 1.17% | 0% | Maternal age (inverse association) and take-out food consumption (positive association) were independent influencing factors of the prevalence of maternal total antibodies; maternal age (inverse association) was an independent influencing factor of the prevalence of maternal IgM; maternal cosmetic use (positive association) and take-out food consumption (positive association) were independent influencing factors of the prevalence of maternal IgG; no other association was found between the prevalence of maternal antibodies (either total or any isotype) and all other demographic factors. | Direct ELISA & competitive ELISA |
| | Zhejiang, China | Newborns: 256 | 129/127 | NA | 5.47% | 0% | 5.47% (IgG1：2.73%；IgG2：2.73%；IgG3 & IgG4：0%) | 0% | 0% | A positive association between total newborn antibody prevalence and maternal cosmetic use was observed; no other association was found between total newborn antibody prevalence and all other demographic factors. | Direct ELISA & competitive ELISA |

Abbreviations: GER, Germany; NA, not applicable. [a]Total Ab included all anti-PEG antibody isotypes.

#### Table S2. Levels of pre-existing anti-PEG antibodies in seropositive general adults across different studies

| Year[Ref] | Levels (ng/mL) of anti-PEG antibodies | | | | Correlations between the levels of anti-PEG antibodies and demographic characteristics | Detection cutoffs |
|---|---|---|---|---|---|---|
| | Total[a] | IgM | IgG | IgE | | |
| 1984[30] | NA | NA | NA | NA | NA | NA |
| 2016[31] | NA | NA | NA | NA | NA | 1:20 titer[b] |
| | NA | NA | NA | NA | NA | 1:20 titer[c] |
| 2016[2] | NA | Range: 300 - 238000;<br>Mean: 5760;<br>**Median: 1790** | Range: 100 - 57300;<br>Mean: 1780;<br>**Median: 960** | NA | IgG levels were negatively correlated with donor age; IgM levels showed no correlation with donor age; neither IgM nor IgG levels were associated with donor gender. | NA[d] |
| 2016[32e] | NA | Range: 0.0 - 2858.4;<br>**Median: 19.5** | Range: 0.0 - 6488.6;<br>**Median: 49.3** | NA | IgG levels were inversely related to donor age, while IgM levels showed no such link; females had relatively higher IgM levels, but gender didn't affect IgG levels; neither IgM nor IgG levels were associated with donor race. | 14.2, 15.1, 3.9, 4.4 and 6.4 ng/mL for anti-PEG IgG1, IgG2, IgG3, IgG4 and IgM, respectively[f] |
| | NA | Range: 0.0 - 101.2 for 1970s<br>**Median: 9.1 for 1970s**<br>Range: 0.0 - 233.9 for 1980s<br>**Median: 14.0 for 1980s**<br>Range: 0.0 - 874.6 for 1990s<br>**Median: 33.2 for 1990s** | Range: 0.0 - 2685.9 for 1970s<br>**Median: 31.6 for 1970s**<br>Range: 0.0 - 2071.9 for 1980s<br>**Median: 12.6 for 1980s**<br>Range: 0.0 - 3504.2 for 1990s<br>**Median: 25.3 for 1990s** | NA | NA | |
| 2021[33] | NA | Range: 26 - 11604;<br>Mean: 930;<br>**Median: 310** | Range: 39 - 18713;<br>Mean: 880;<br>**Median: 260** | NA | No correlation between donor age, gender or race and the levels of either IgM or IgG. | 35 and 25 ng/mL for anti-PEG IgG and IgM, respectively[g] |
| 2023[34] | NA | NA | NA | NA | No correlation between donor age and the levels of either IgM or IgG. | NA[h] |
| 2024[35] | NA | NA | NA | NA | NA | NA[i] |
| 2024<br>(Present study) | Pregnant woman<br>Range: 55.43 - 23649.14;<br>Mean: 941.67;<br>**Median: 292.02** | Pregnant woman<br>Range: 55.43 - 23649.14;<br>Mean: 1068.95;<br>**Median: 175.07** | Pregnant woman<br>Range: 75.54 - 2604.89;<br>Mean: 675.47;<br>**Median: 495.41** | Pregnant woman<br>Range: 0 - 0;<br>Mean: 0;<br>**Median: 0** | Maternal age (positive association) and cosmetic use (positive association) were independent influencing factors of maternal total antibody levels; maternal cosmetic use (positive association) was an independent influencing factor of maternal IgM levels; Maternal age (positive association) and take-out food consumption (positive association) were independent influencing factors of maternal total IgG levels; no other association was found between the levels of maternal antibodies (either total or any isotype) and all other demographic factors. | 14.7, 4.4, 0.3, 1.2, 1.2 and 7.7 ng/mL for anti-PEG IgG1, IgG2, IgG3, IgG4, IgM and IgE, respectively[j] |
| | Newborns<br>Range: 100.24 - 1513.98;<br>Mean: 443.27;<br>**Median: 306.22** | Newborns<br>Range: 0 - 0;<br>Mean: 0;<br>**Median: 0** | Newborns<br>Range: 100.24 - 1513.98;<br>Mean: 443.27;<br>**Median: 306.22** | Newborns<br>Range: 0 - 0;<br>Mean: 0;<br>**Median: 0** | Maternal age (positive association) was an independent influencing factor of total newborn antibody levels; no other association was found between total newborn antibody levels and all other demographic factors. | 11.4, 4.3, 0.2, 0.9, 0.1 and 9.4 ng/mL for anti-PEG IgG1, IgG2, IgG3, IgG4, IgM and IgE, respectively[j] |

Abbreviations: NA, not applicable; OD, absorbance values.

[a]Total Ab included all anti-PEG antibody isotypes.

[b]The study employed flow cytometry without constructing a standard curve to semi-quantitatively (a method between qualitative and quantitative) evaluate anti-PEG antibodies in serum samples by determining the antibody titer via serial dilution, with a titer of 1:20 defined as the detection cutoff. The antibody titer, typically expressed as "1:X," indicates the maximum dilution at which the antibody can be detected. For instance, a titer of 1:32,000 means the antibody is still detectable at a 32,000-fold dilution but not at a higher dilution.

[c]The study utilized ELISA without constructing a standard curve to semi-quantitatively assess anti-PEG antibodies in serum samples by determining the antibody titer through serial dilution. An antibody titer of 1:20 was set as the detection cutoff.

[d]The study employed a quantitative direct ELISA, which requires the construction of a standard curve, to measure the relative concentration of anti-PEG antibodies in serum samples. The article provides the formula for calculating the detection cutoff as $OD_{sample} \geq 3 \times$ mean $OD_{background}$, but does not specify an exact value for the detection cutoff.

[e]The study delineates the distribution of anti-PEG antibody levels across the entire spectrum of the healthy human population, encompassing both individuals with detectable levels of anti-PEG antibodies (anti-PEG antibody-positive) and those without such detectable levels (anti-PEG antibody-negative).

[f]The study used a quantitative competitive ELISA, which involves creating a standard curve, to determine the relative concentration of anti-PEG antibodies in serum samples. The calculation for the detection cutoff is detailed as $\overline{X} + \mathrm{SD}f^{2}$, where $\overline{X}$ is the mean absorbance of negative controls, SD is their standard deviation, and $f$ is a multiplier based on the number of negative controls and a 95% confidence level (for details, see eReference 4). Additionally, specific detection cutoff values are provided within the text.

[g]The study utilized flow cytometry, which required the construction of a standard curve, to assess the relative concentration of anti-PEG antibodies in serum samples. Although the calculation formula for the detection cutoff was not provided, specific values were given within the text.

[h]The study employed a quantitative direct ELISA, which requires the construction of a standard curve, to measure the relative concentration of anti-PEG antibodies in serum samples. The article provides the formula for calculating the detection cutoff as $OD_{sample} \geq \text{mean } OD_{background} + 3 \times SD_{background}$, but does not specify an exact value for the detection cutoff.

[i]The study employed a quantitative competitive ELISA, which requires the construction of a standard curve, to measure the relative concentration of anti-PEG antibodies in serum samples. The article provides the formula for calculating the detection cutoff as $OD_{sample} \geq \text{mean } OD_{background} + 10 \times SD_{background}$, but does not specify an exact value for the detection cutoff.

[j]The present study used a quantitative direct ELISA, which involves creating a standard curve, to determine the relative concentration of anti-PEG antibodies in serum samples. The calculation for the detection cutoff is detailed as $\overline{X} + \mathrm{SD}f^{2}$, where $\overline{X}$ is the mean absorbance of negative controls, SD is their standard deviation, and $f$ is a multiplier based on the number of negative controls and a 95% confidence level (for details, see eMethods 3). Additionally, specific detection cutoff values are provided within the text.

#### Table S3. Precision of direct ELISA for quantification of maternal and newborn anti-PEG antibodies

| | Intra-assay precision (CV%)[a] | | Inter-assay precision (CV%)[b] |
|---|---|---|---|
| | Anti-PEG antibody standards | Samples | |
| **Pregnant women** | | | |
| Anti-PEG IgG1 | 3.532 ± 1.694 | 3.640 ± 4.892 | 4.353 ± 1.870 |
| Anti-PEG IgG2 | 3.124 ± 2.404 | 6.018 ± 5.403 | 4.396 ± 1.393 |
| Anti-PEG IgG3 | 4.247 ± 3.403 | 3.982 ± 4.143 | 7.825 ± 4.511 |
| Anti-PEG IgG4 | 4.776 ± 2.557 | 4.477 ± 3.853 | 5.120 ± 2.263 |
| Anti-PEG IgM | 1.656 ± 0.825 | 2.864 ± 2.450 | 4.596 ± 2.043 |
| Anti-PEG IgE | 4.150 ± 2.756 | 2.346 ± 2.476 | 5.236 ± 2.265 |
| **Newborns** | | | |
| Anti-PEG IgG1 | 3.964 ± 3.573 | 3.553 ± 4.379 | 12.725 ± 11.025 |
| Anti-PEG IgG2 | 2.566 ± 1.855 | 4.245 ± 3.635 | 9.102 ± 6.077 |
| Anti-PEG IgG3 | 4.004 ± 1.728 | 3.403 ± 3.213 | 9.963 ± 5.808 |
| Anti-PEG IgG4 | 4.494 ± 4.509 | 3.220 ± 2.679 | 4.778 ± 3.877 |
| Anti-PEG IgM | 2.814 ± 1.138 | 3.163 ± 3.054 | 3.748 ± 0.592 |
| Anti-PEG IgE | 4.390 ± 3.514 | 1.351 ± 1.568 | 5.599 ± 2.640 |

[a]Intra-assay precision was evaluated by calculating the coefficient of variation (CV% = (Standard deviation/Mean) × 100%) for all anti-PEG antibody standards and samples in two independent batches of direct ELISA (see eMethods 5 for acceptance criteria).

[b]Inter-assay precision was determined by calculating the coefficient of variation for serially diluted anti-PEG antibody standards among two independent batches of direct ELISA (see eMethods 5 for acceptance criteria). Data were presented as "mean ± standard deviation", with n = 14 for intra-assay precision (CV%) of anti-PEG antibody standards in all direct ELISA, n = 256 for intra-assay precision (CV%) of serum samples in all direct ELISA, n = 7 for inter-assay precision (CV%) in all direct ELISA.

#### Figure S1. Detection of anti-PEG IgG1 in maternal serum samples by direct ELISA (Batch 1)

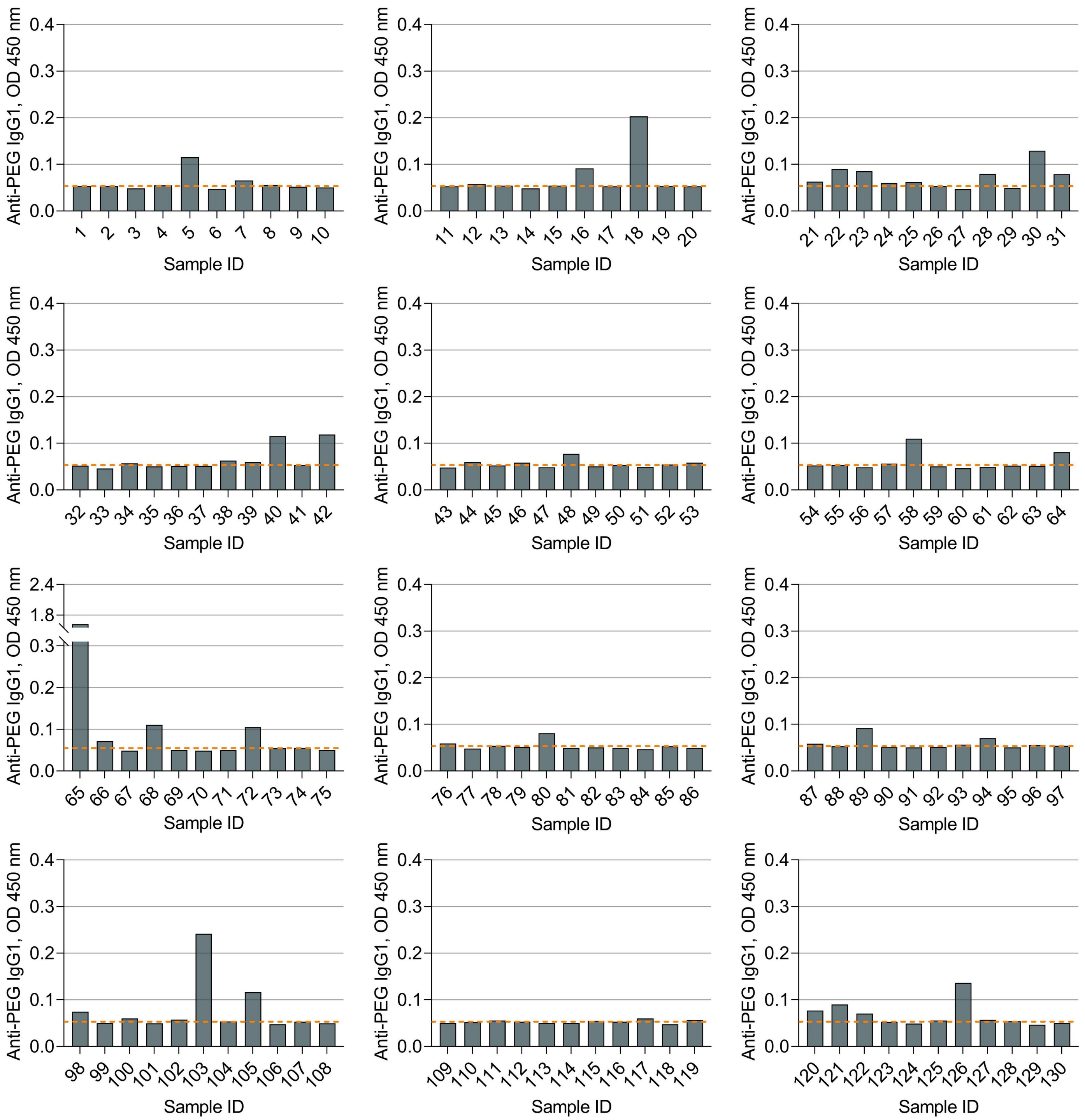


Dotted lines indicate the detection cutoff of anti-PEG IgG1 (see eMethods 3 for the establishment of detection cutoff of anti-PEG IgG1).

**Figure S2. Detection of anti-PEG IgG1 in maternal serum samples by direct ELISA (Batch 2)**

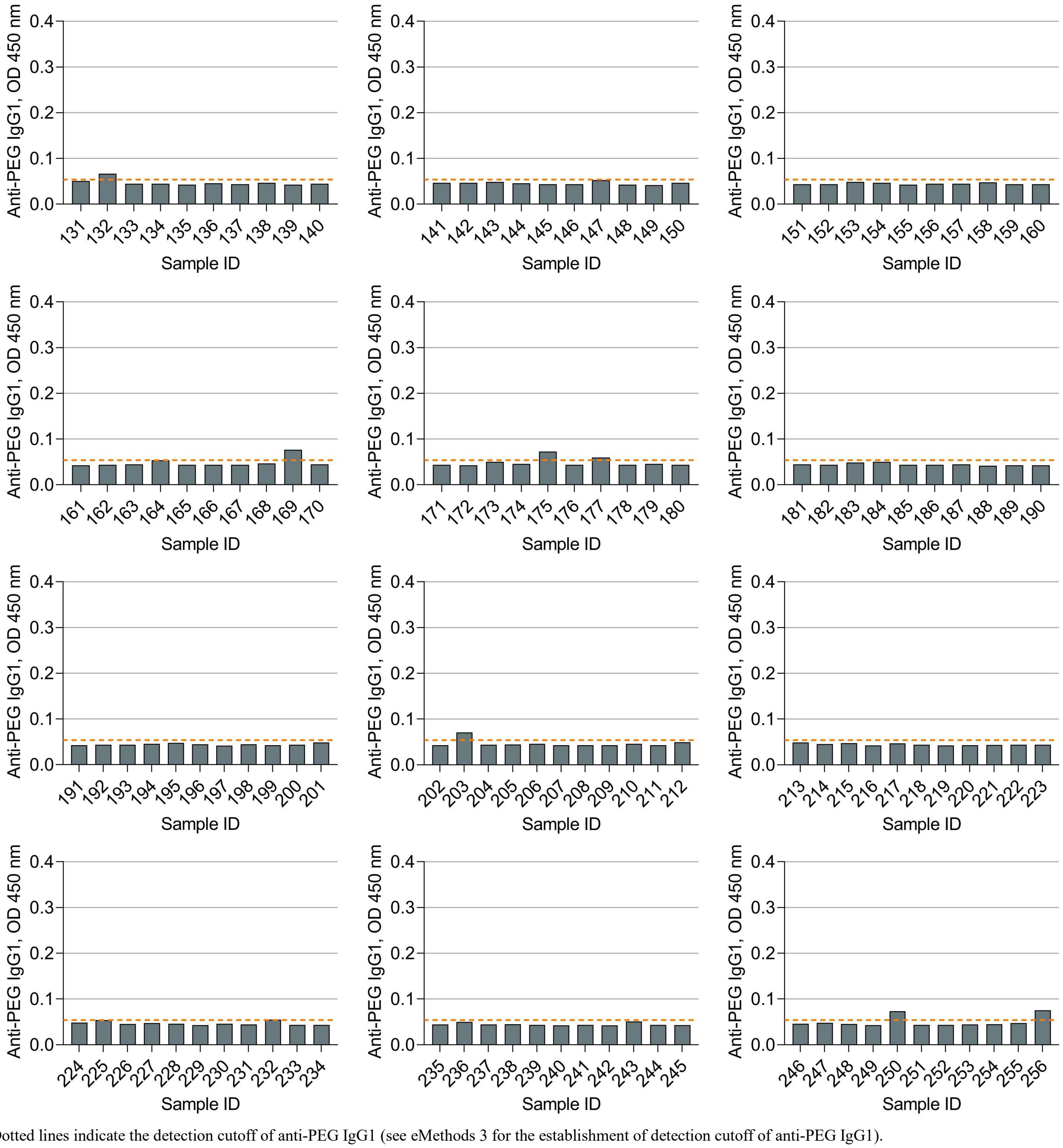


Dotted lines indicate the detection cutoff of anti-PEG IgG1 (see eMethods 3 for the establishment of detection cutoff of anti-PEG IgG1).

#### Figure S3. Detection of anti-PEG IgG1 in newborn serum samples by direct ELISA (Batch 1)

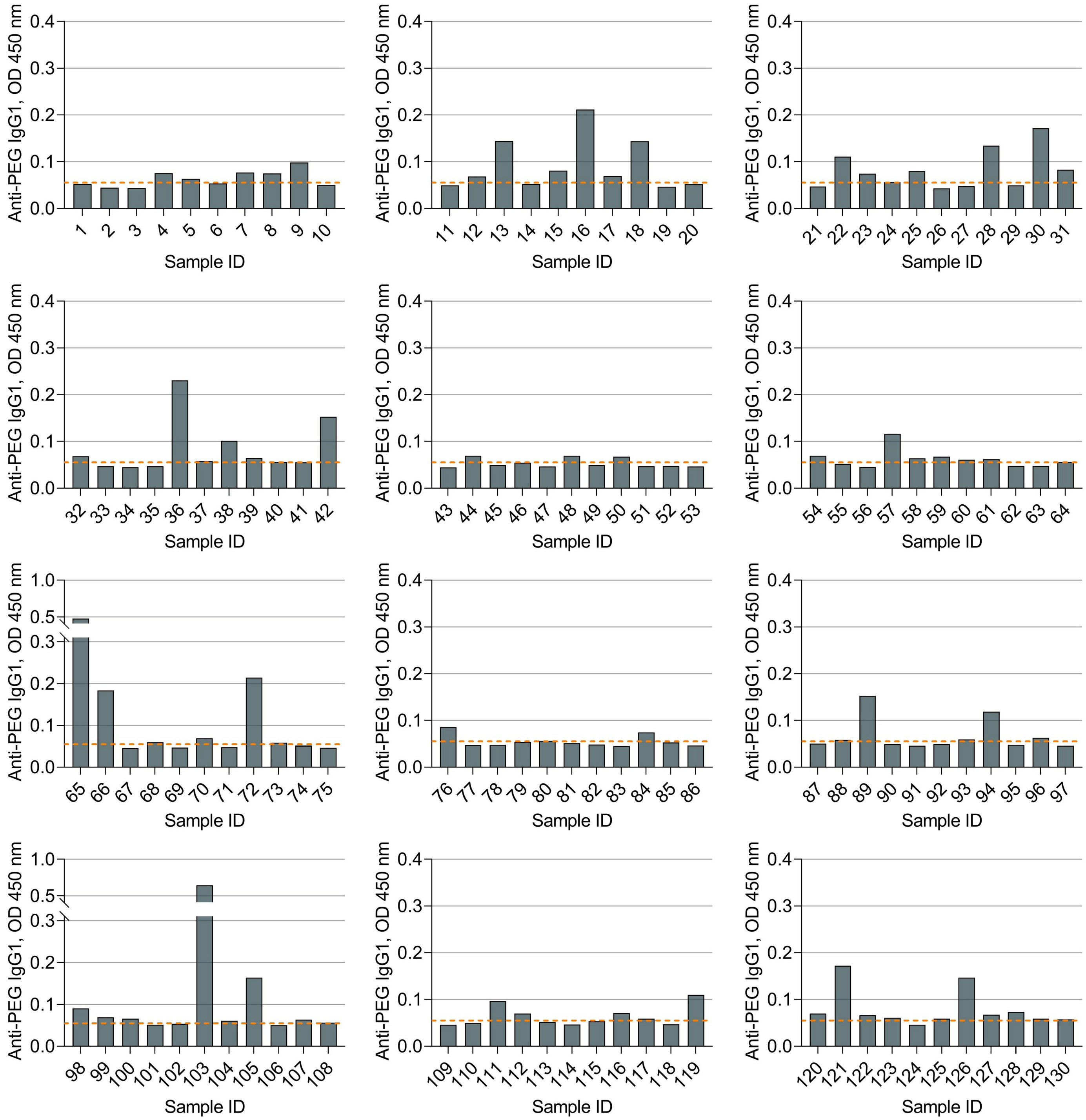


Dotted lines indicate the detection cutoff of anti-PEG IgG1 (see eMethods 3 for the establishment of detection cutoff of anti-PEG IgG1).

**Figure S4. Detection of anti-PEG IgG1 in newborn serum samples by direct ELISA (Batch 2)**

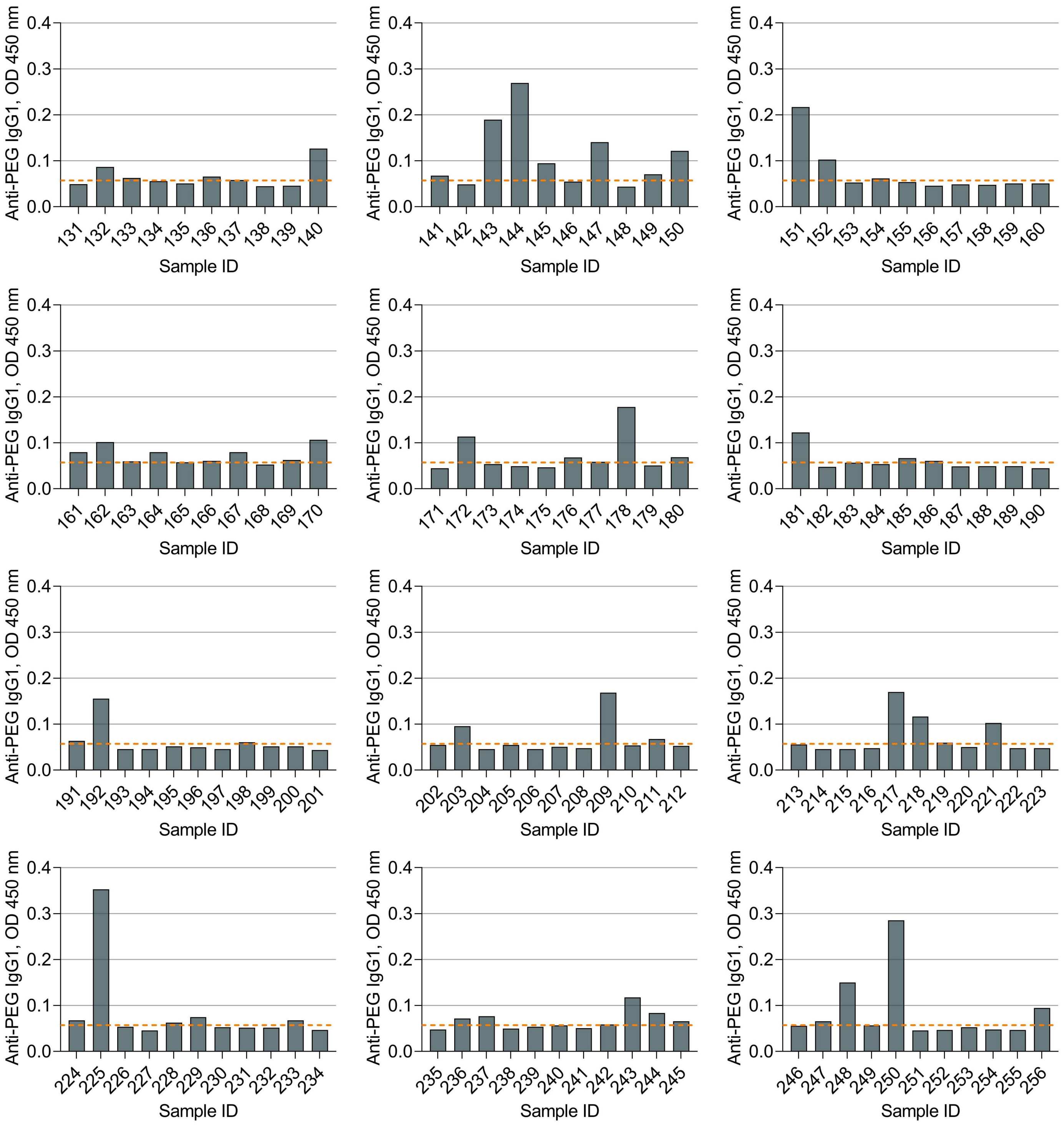


Dotted lines indicate the detection cutoff of anti-PEG IgG1 (see eMethods 3 for the establishment of detection cutoff of anti-PEG IgG1).

**Figure S5. Detection of anti-PEG IgG2 in maternal serum samples by direct ELISA (Batch 1)**

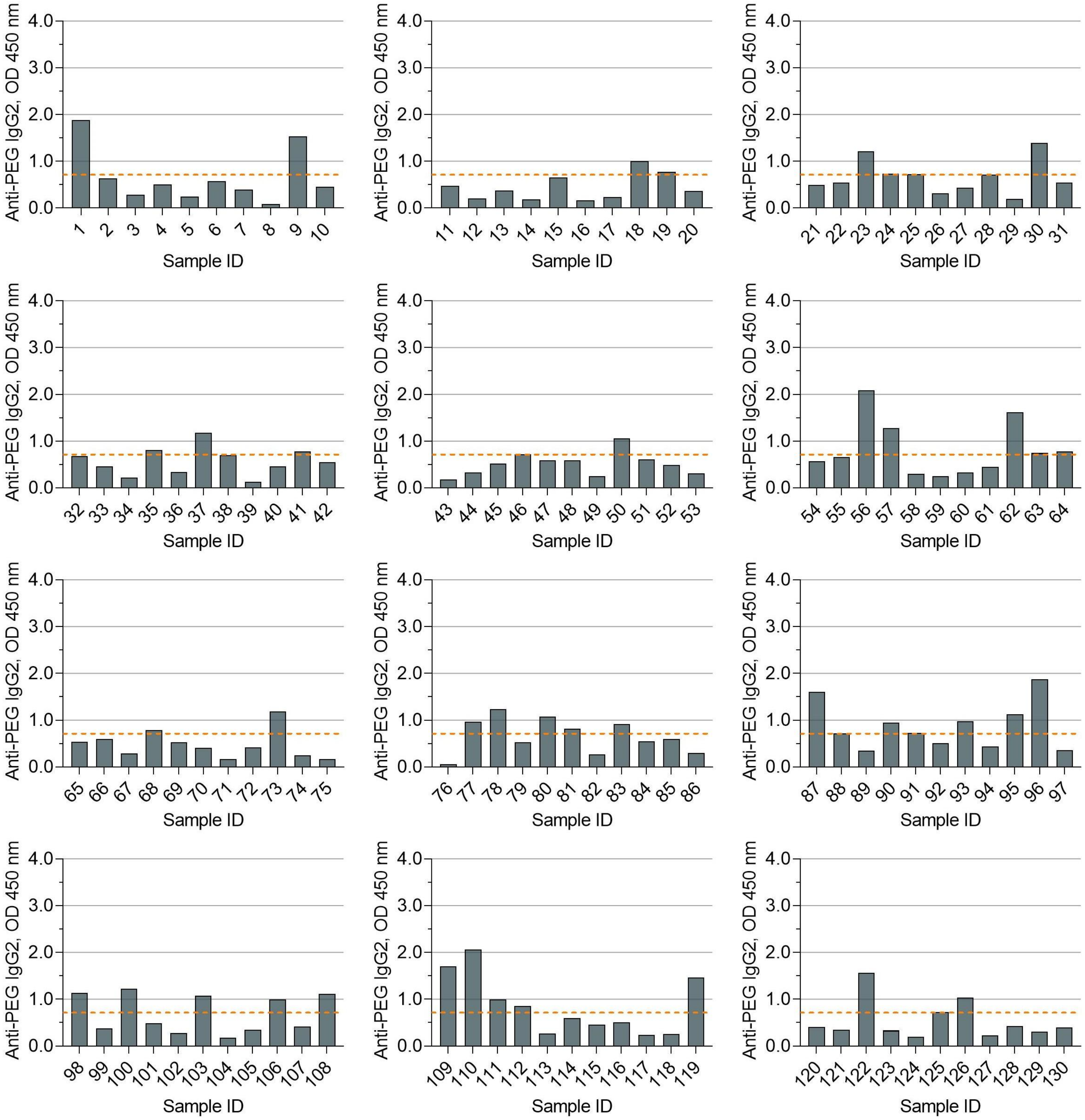


Dotted lines indicate the detection cutoff of anti-PEG IgG2 (see eMethods 3 for the establishment of detection cutoff of anti-PEG IgG2).

**Figure S6. Detection of anti-PEG IgG2 in maternal serum samples by direct ELISA (Batch 2)**

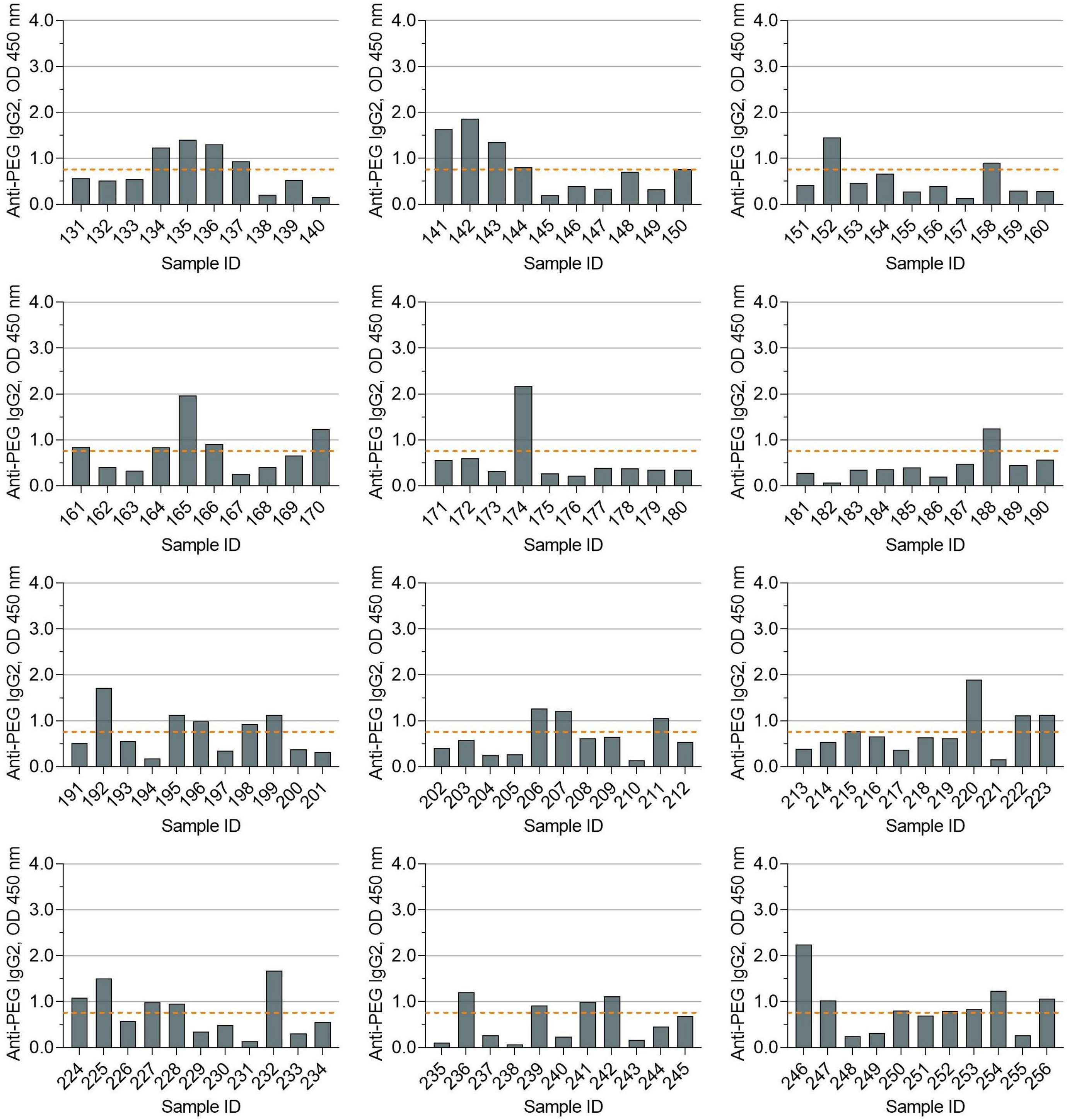


Dotted lines indicate the detection cutoff of anti-PEG IgG2 (see eMethods 3 for the establishment of detection cutoff of anti-PEG IgG2).

### Figure S7. Detection of anti-PEG IgG2 in newborn serum samples by direct ELISA (Batch 1)

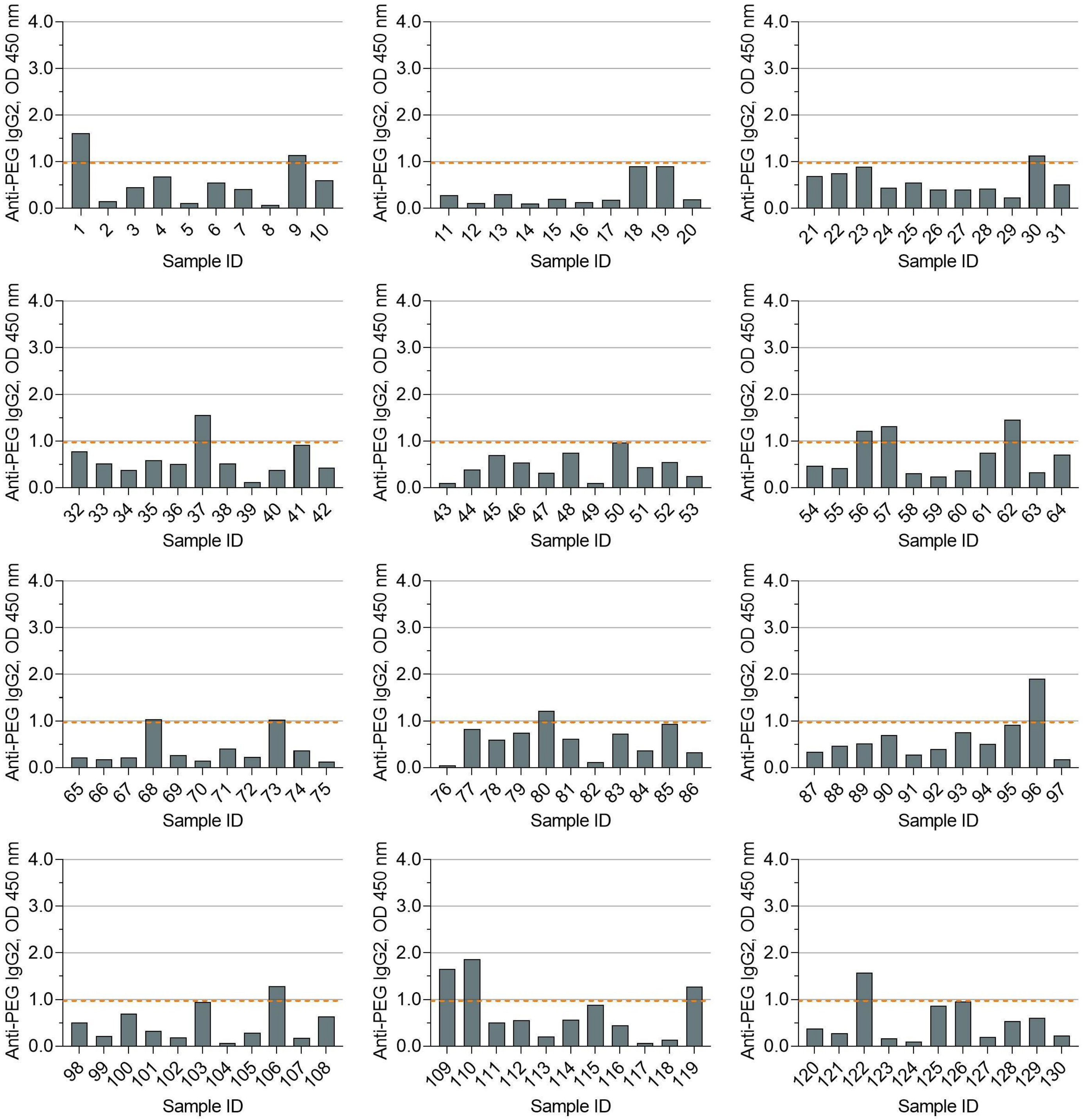


Dotted lines indicate the detection cutoff of anti-PEG IgG2 (see eMethods 3 for the establishment of detection cutoff of anti-PEG IgG2).

#### Figure S8. Detection of anti-PEG IgG2 in newborn serum samples by direct ELISA (Batch 2)

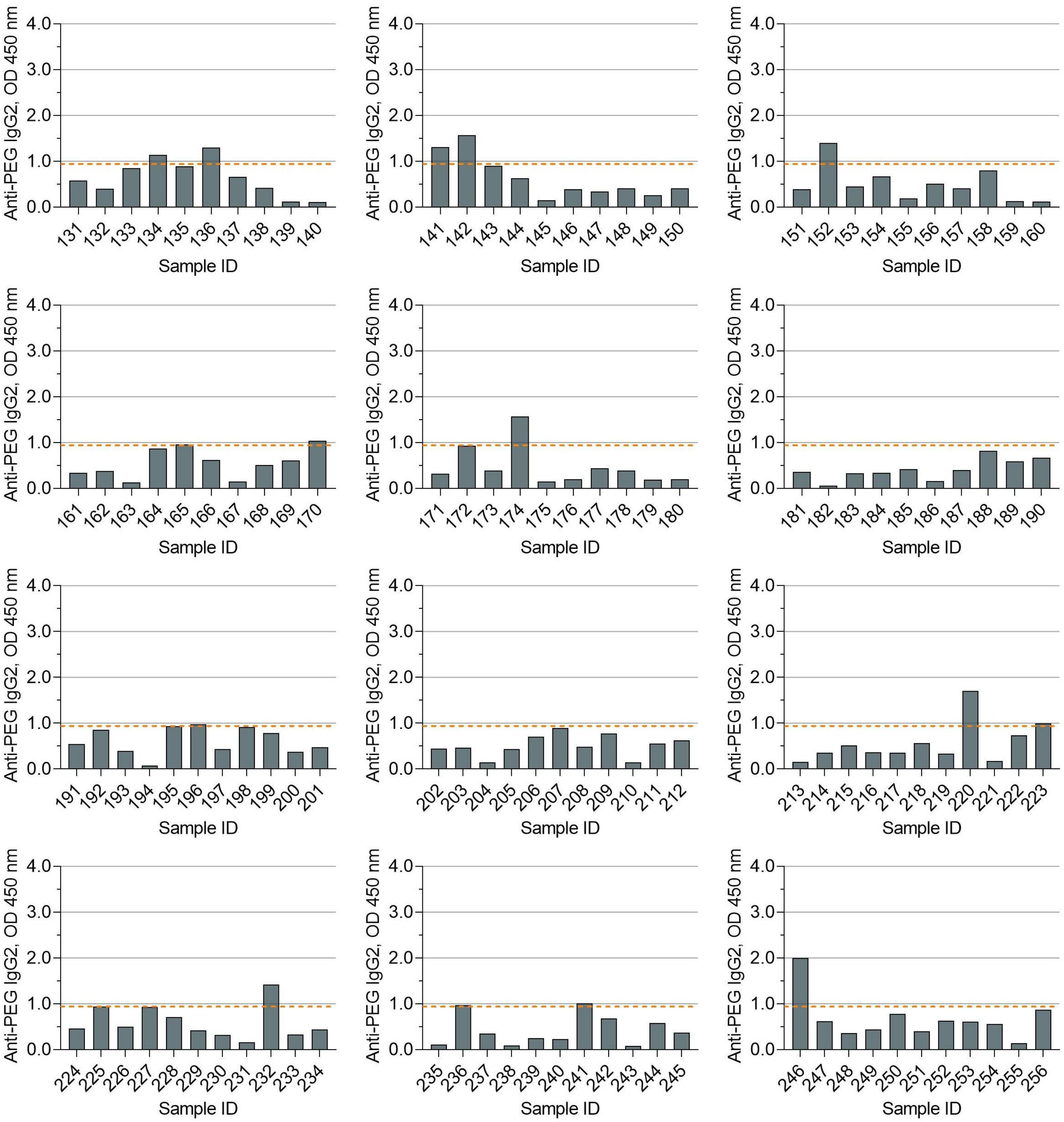


Dotted lines indicate the detection cutoff of anti-PEG IgG2 (see eMethods 3 for the establishment of detection cutoff of anti-PEG IgG2).

**Figure S9. Detection of anti-PEG IgG3 in maternal serum samples by direct ELISA (Batch 1)**

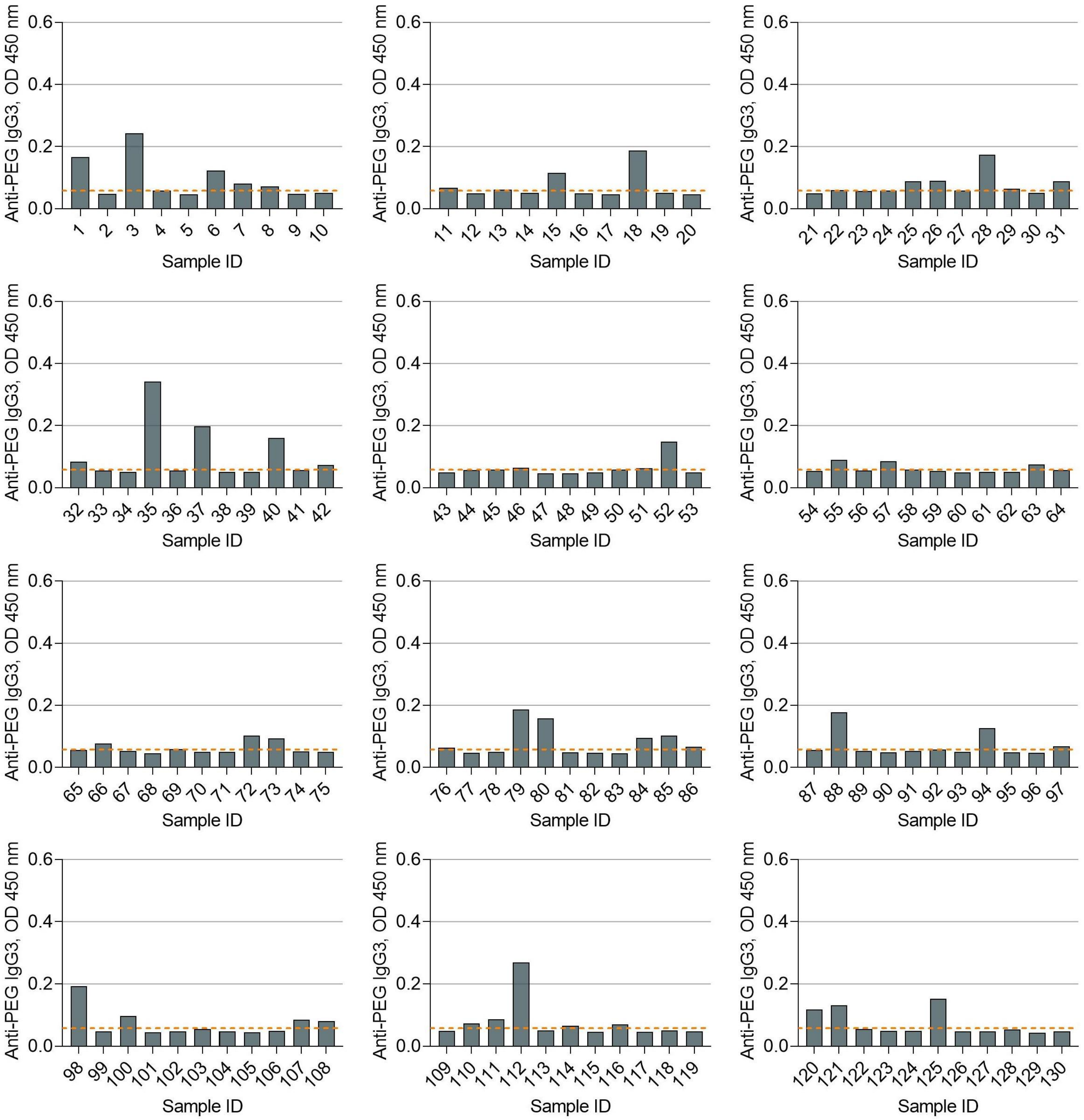


Dotted lines indicate the detection cutoff of anti-PEG IgG3 (see eMethods 3 for the establishment of detection cutoff of anti-PEG IgG3).

**Figure S10. Detection of anti-PEG IgG3 in maternal serum samples by direct ELISA (Batch 2)**

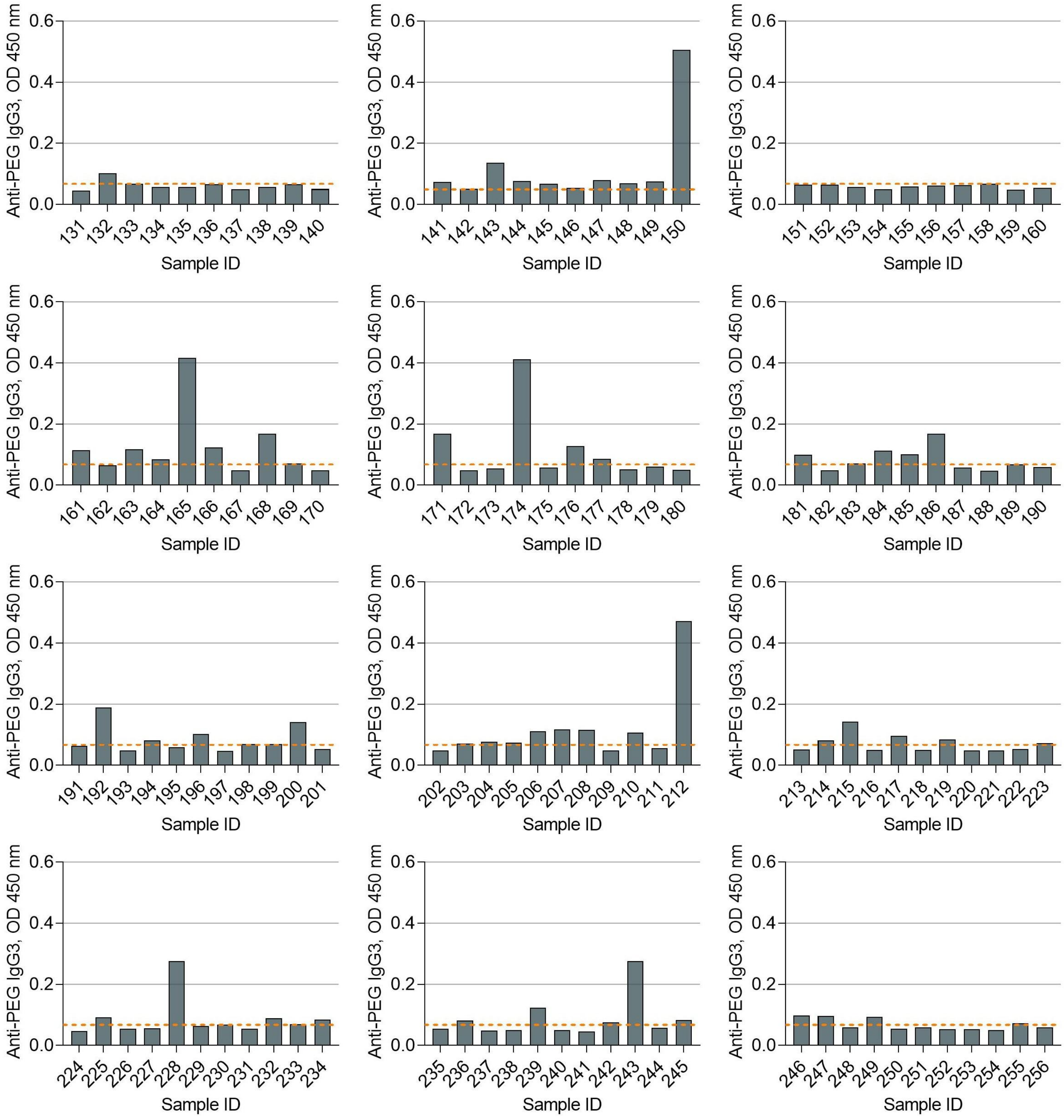


Dotted lines indicate the detection cutoff of anti-PEG IgG3 (see eMethods 3 for the establishment of detection cutoff of anti-PEG IgG3).

**Figure S11. Detection of anti-PEG IgG3 in newborn serum samples by direct ELISA (Batch 1)**

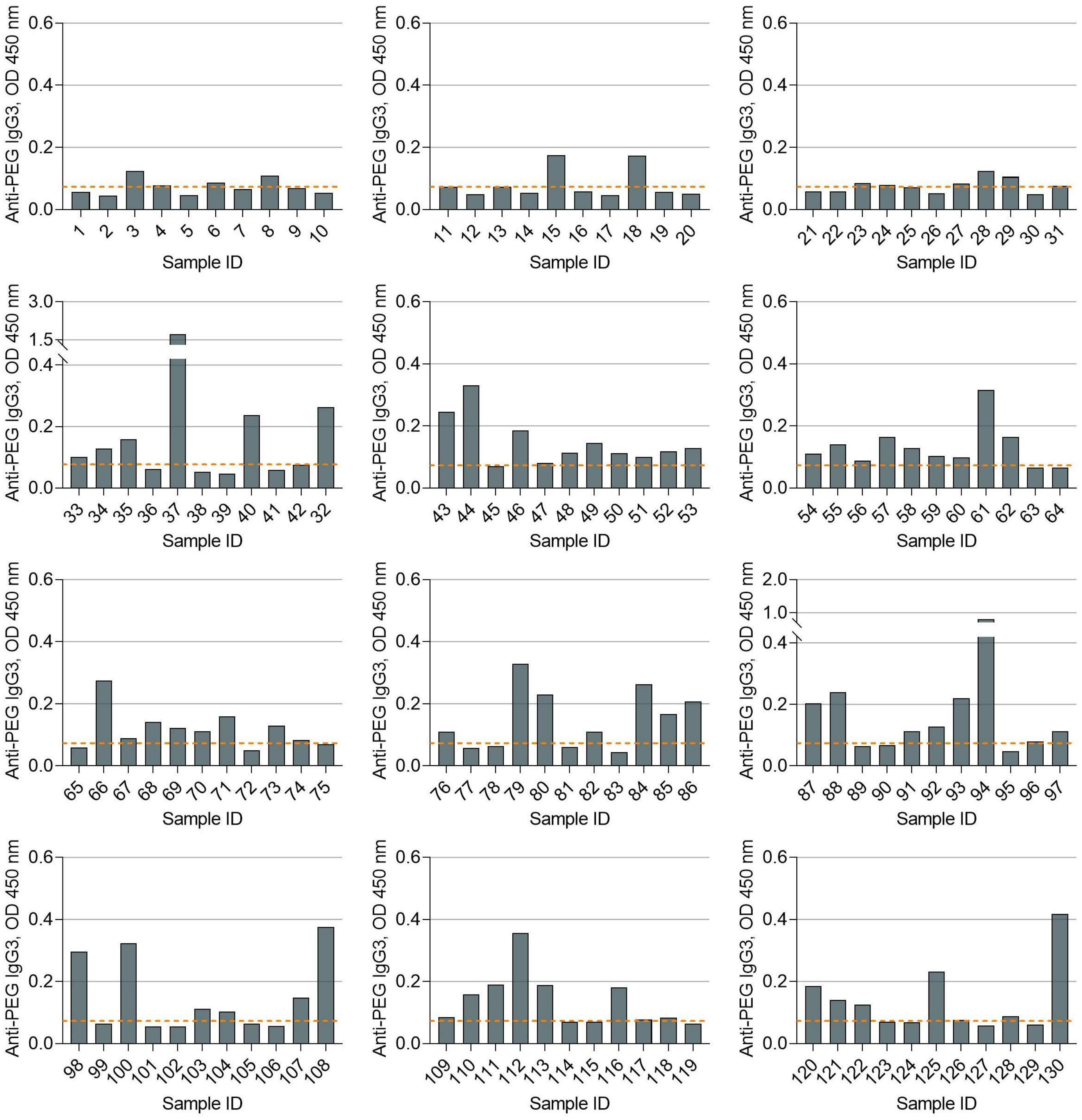


Dotted lines indicate the detection cutoff of anti-PEG IgG3 (see eMethods 3 for the establishment of detection cutoff of anti-PEG IgG3).

#### Figure S12. Detection of anti-PEG IgG3 in newborn serum samples by direct ELISA (Batch 2)

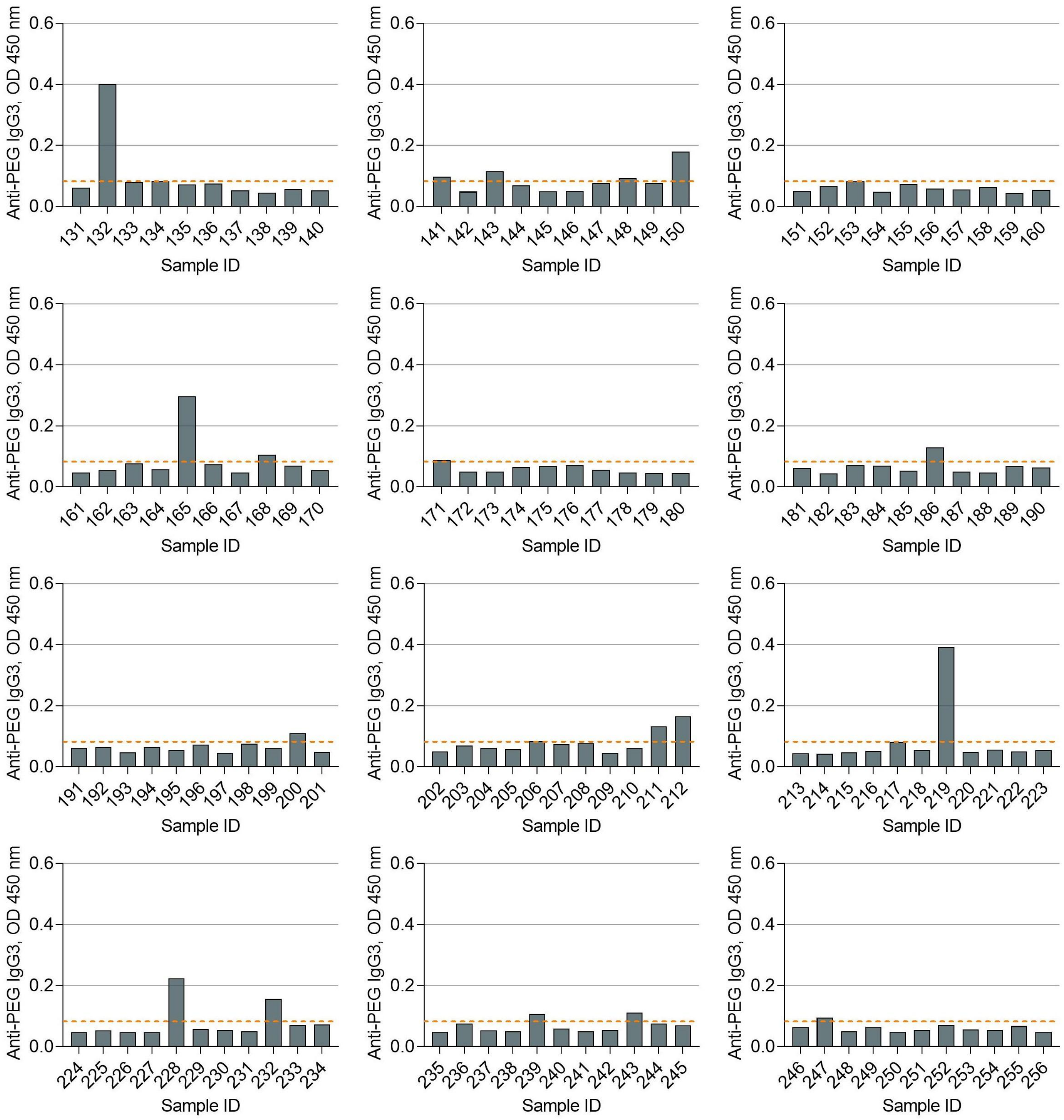


Dotted lines indicate the detection cutoff of anti-PEG IgG3 (see eMethods 3 for the establishment of detection cutoff of anti-PEG IgG3).

#### Figure S13. Detection of anti-PEG IgG4 in maternal serum samples by direct ELISA (Batch 1)

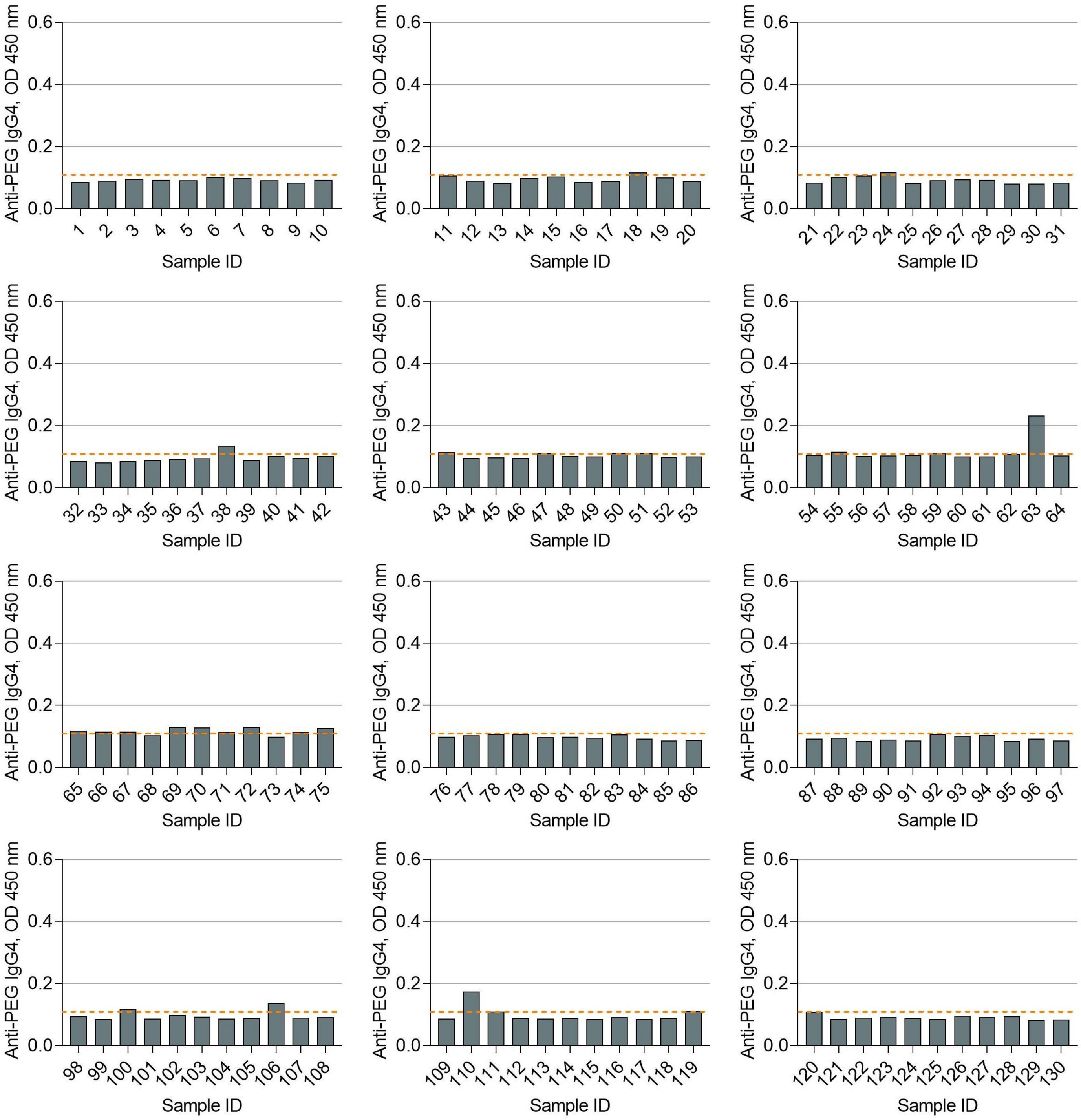


Dotted lines indicate the detection cutoff of anti-PEG IgG4 (see eMethods 3 for the establishment of detection cutoff of anti-PEG IgG4).

**Figure S14. Detection of anti-PEG IgG4 in maternal serum samples by direct ELISA (Batch 2)**

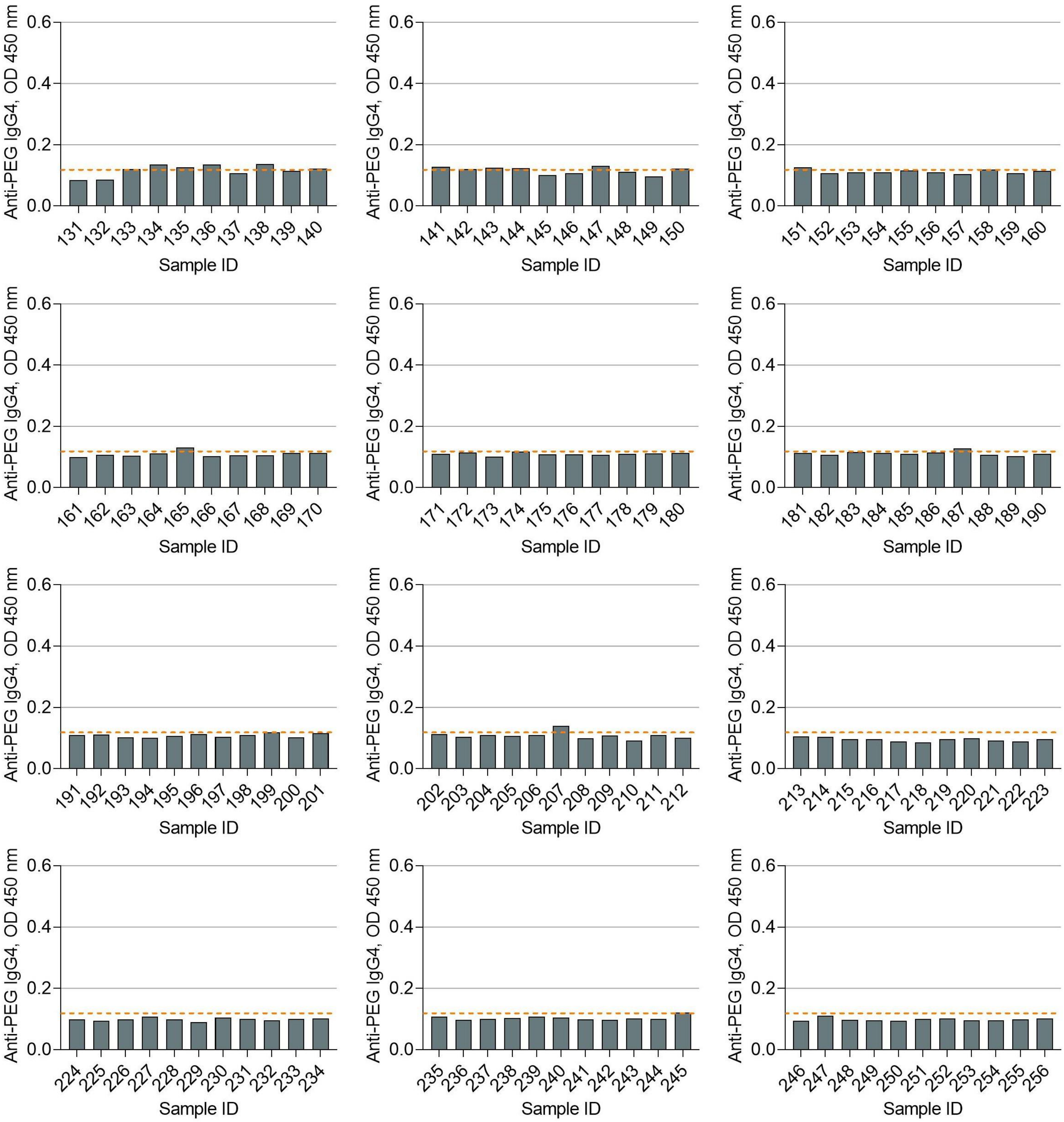


Dotted lines indicate the detection cutoff of anti-PEG IgG4 (see eMethods 3 for the establishment of detection cutoff of anti-PEG IgG4).

**Figure S15. Detection of anti-PEG IgG4 in newborn serum samples by direct ELISA (Batch 1)**

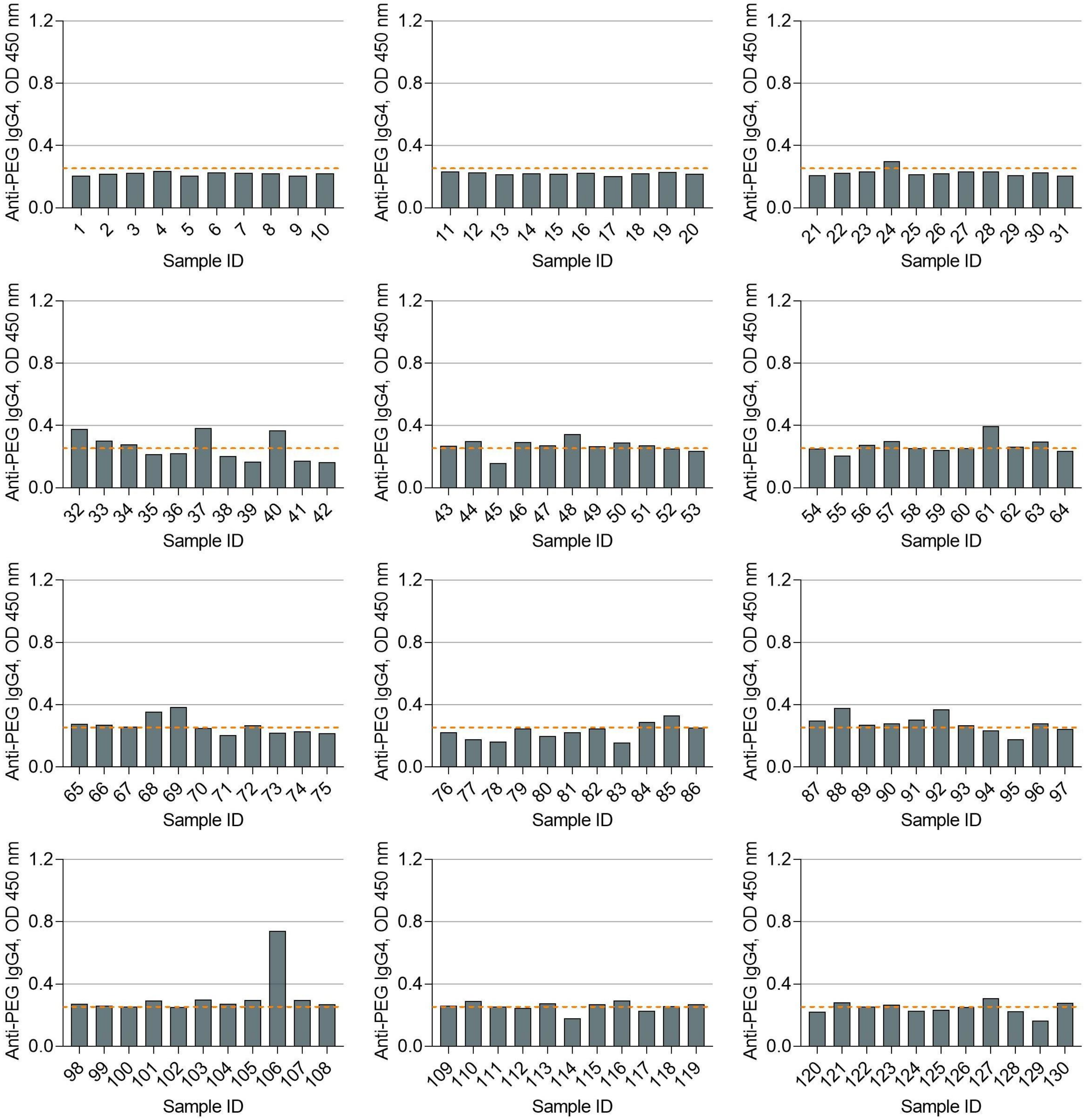


Dotted lines indicate the detection cutoff of anti-PEG IgG4 (see eMethods 3 for the establishment of detection cutoff of anti-PEG IgG4).

#### Figure S16. Detection of anti-PEG IgG4 in newborn serum samples by direct ELISA (Batch 2)

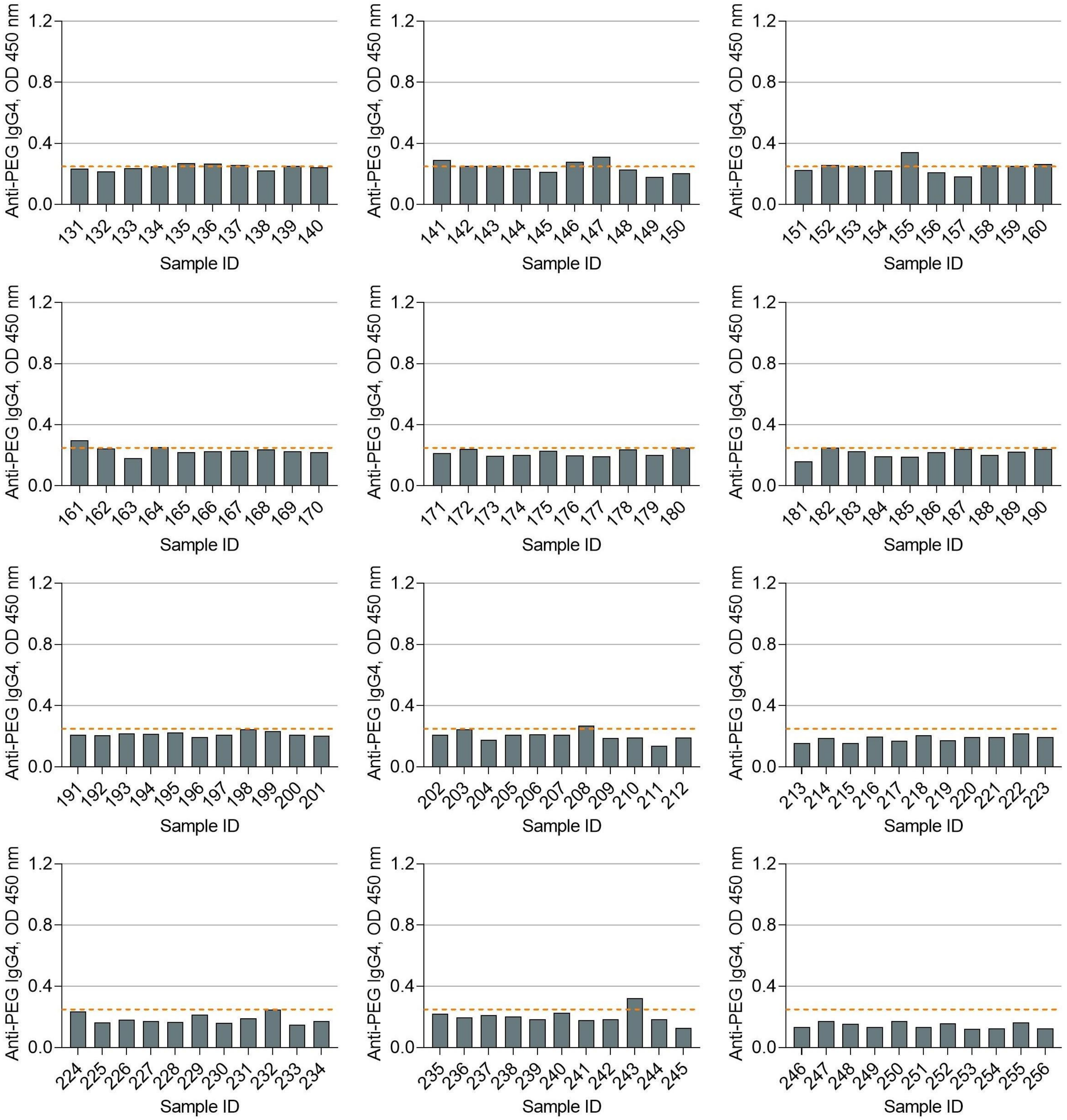


Dotted lines indicate the detection cutoff of anti-PEG IgG4 (see eMethods 3 for the establishment of detection cutoff of anti-PEG IgG4).

### Figure S17. Detection of anti-PEG IgM in maternal serum samples by direct ELISA (Batch 1)

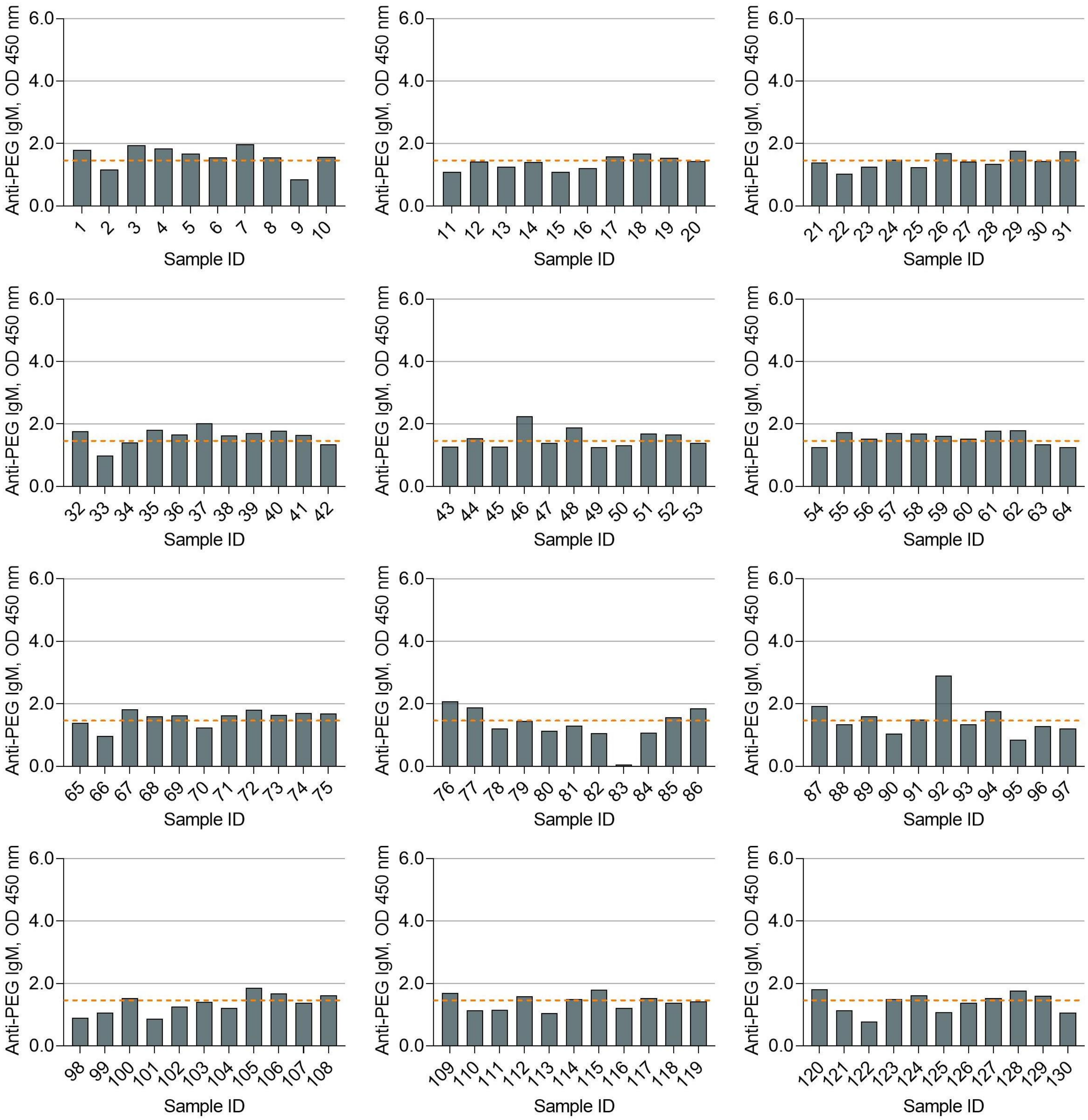


Dotted lines indicate the detection cutoff of anti-PEG IgM (see eMethods 3 for the establishment of detection cutoff of anti-PEG IgM).

**Figure S18. Detection of anti-PEG IgM in maternal serum samples by direct ELISA (Batch 2)**

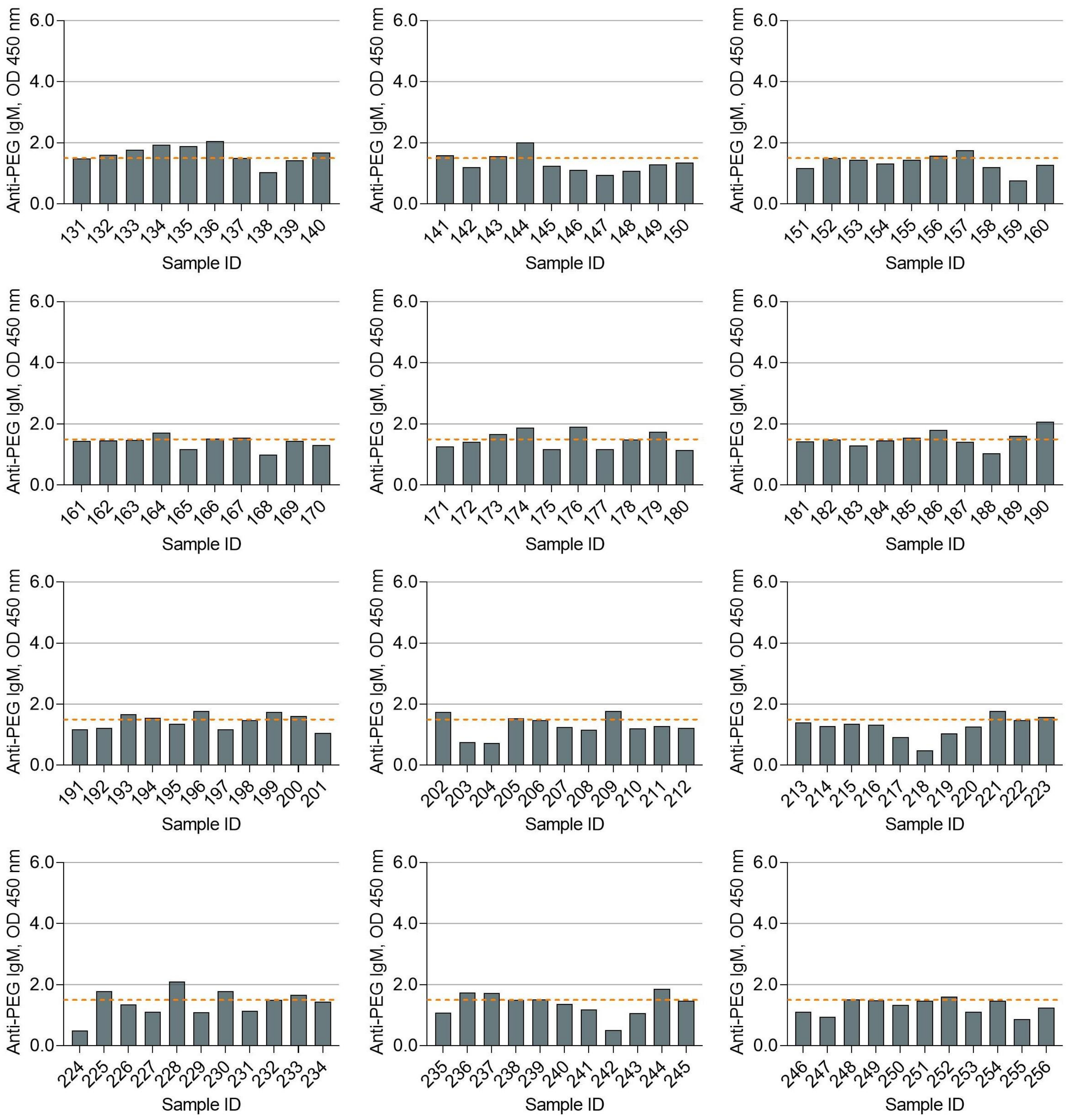


Dotted lines indicate the detection cutoff of anti-PEG IgM (see eMethods 3 for the establishment of detection cutoff of anti-PEG IgM).

#### Figure S19. Detection of anti-PEG IgM in newborn serum samples by direct ELISA (Batch 1)

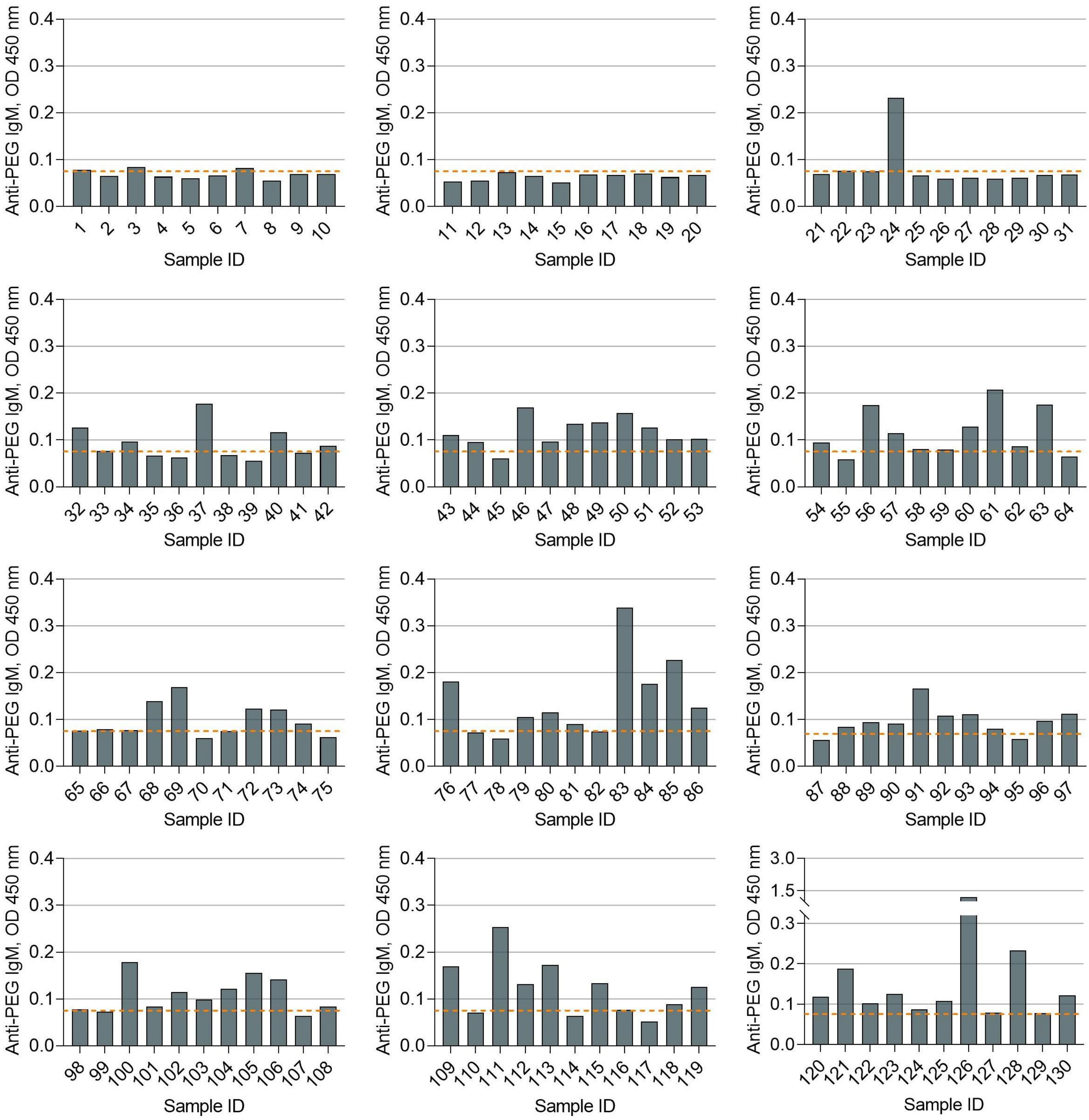


Dotted lines indicate the detection cutoff of anti-PEG IgM (see eMethods 3 for the establishment of detection cutoff of anti-PEG IgM).

#### Figure S20. Detection of anti-PEG IgM in newborn serum samples by direct ELISA (Batch 2)

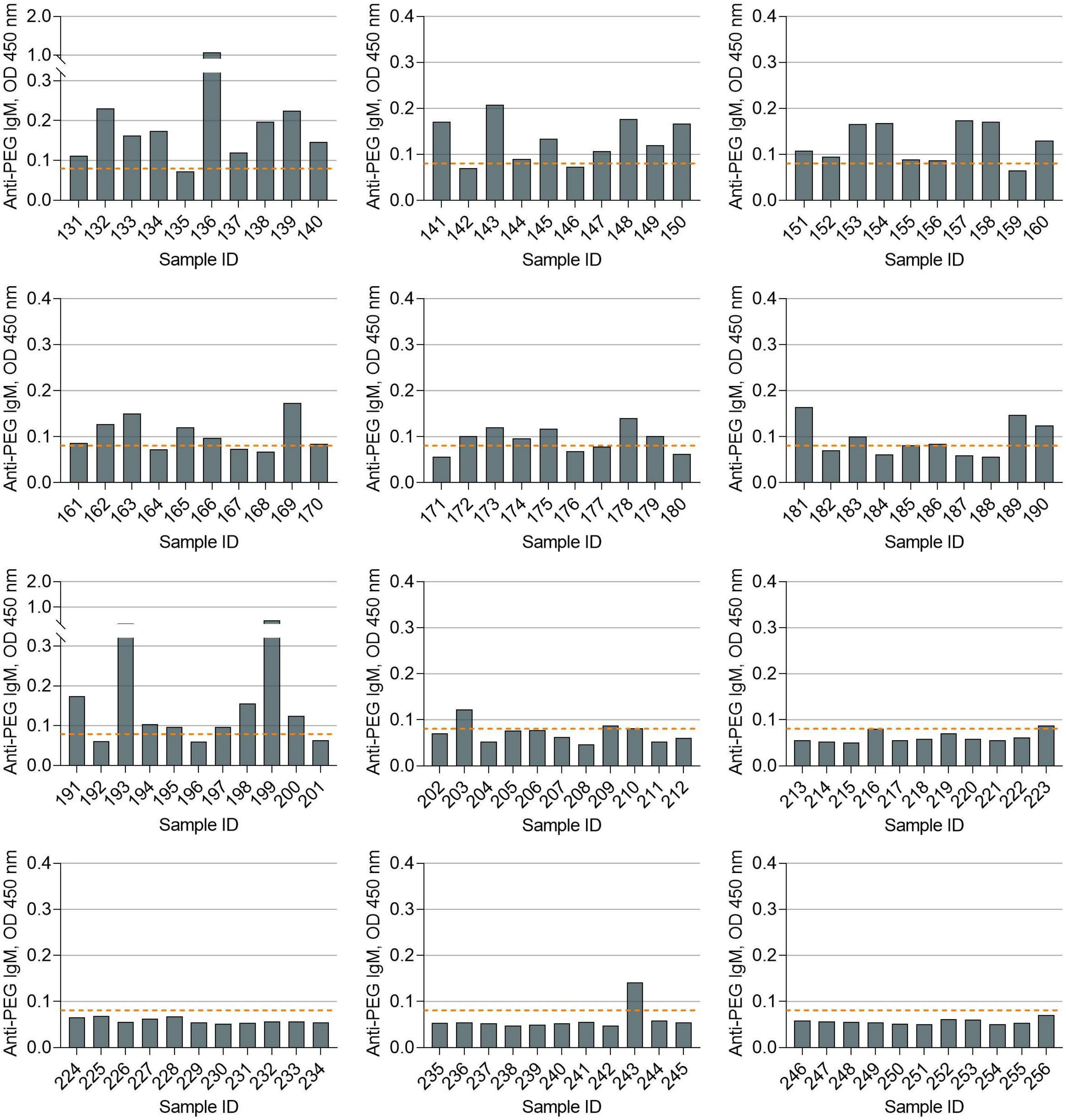


Dotted lines indicate the detection cutoff of anti-PEG IgM (see eMethods 3 for the establishment of detection cutoff of anti-PEG IgM).

### Figure S21. Detection of anti-PEG IgE in maternal serum samples by direct ELISA (Batch 1)

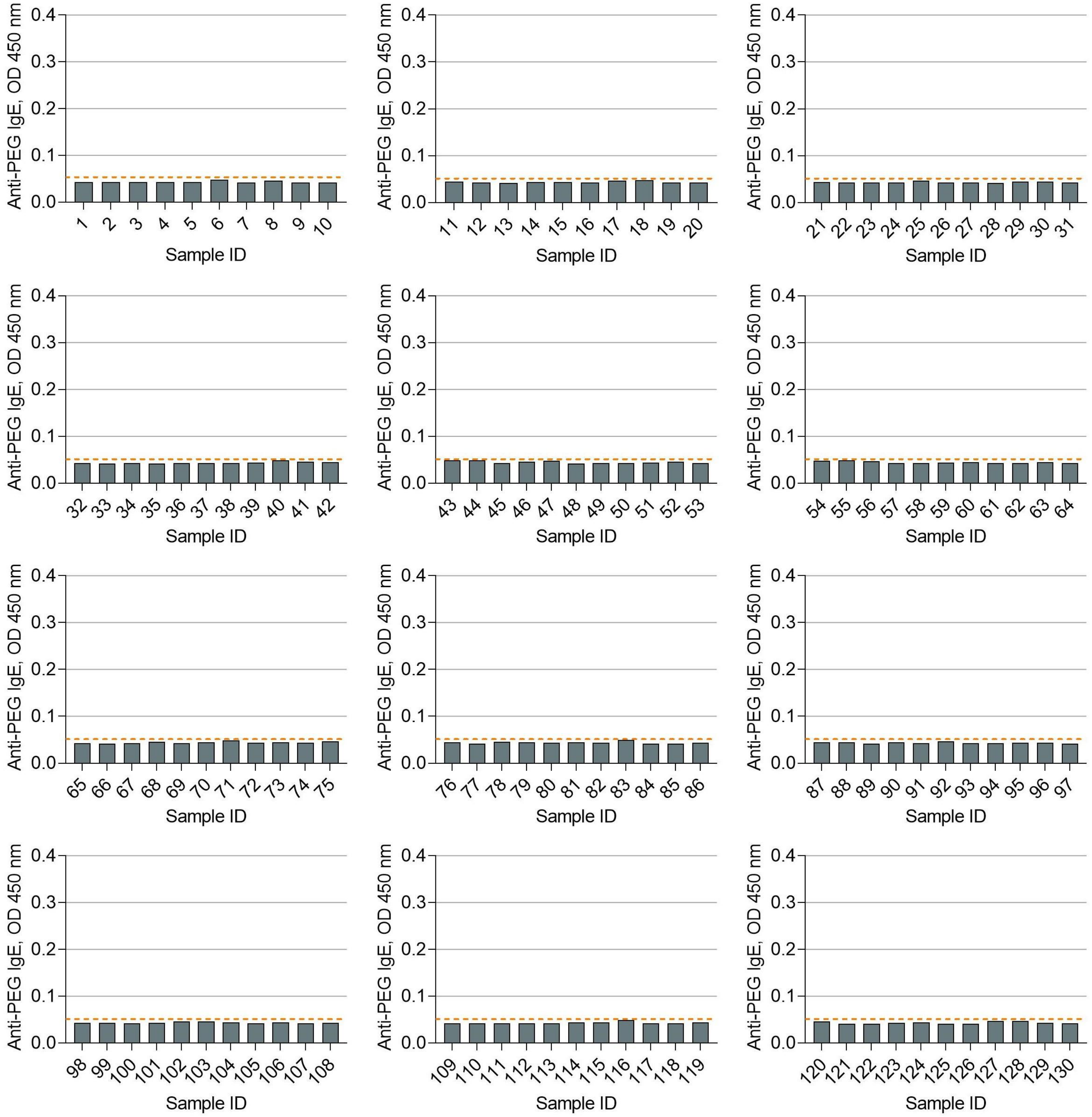


Dotted lines indicate the detection cutoff of anti-PEG IgE (see eMethods 3 for the establishment of detection cutoff of anti-PEG IgE).

#### Figure S22. Detection of anti-PEG IgE in maternal serum samples by direct ELISA (Batch 2)

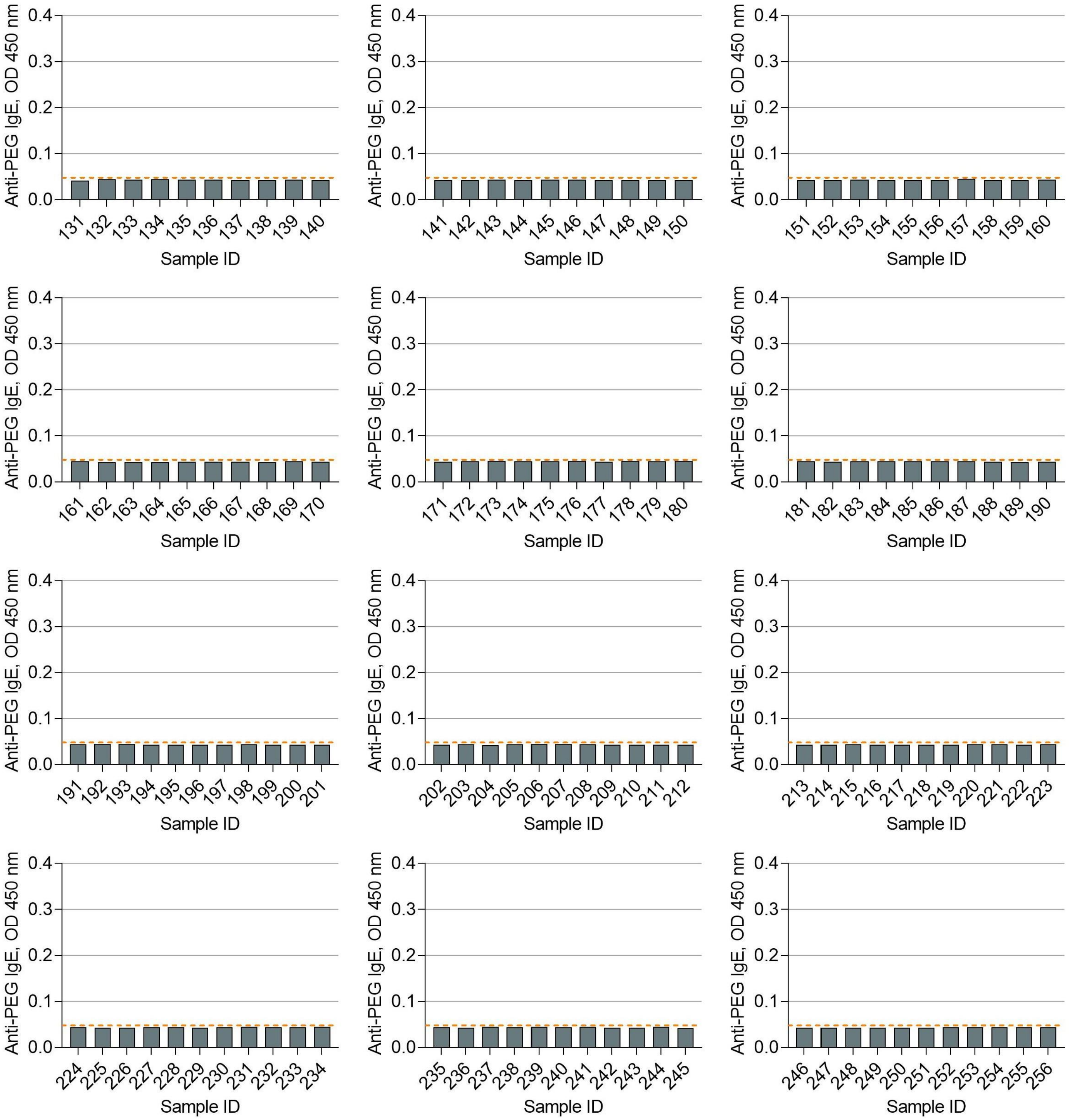


Dotted lines indicate the detection cutoff of anti-PEG IgE (see eMethods 3 for the establishment of detection cutoff of anti-PEG IgE).

#### Figure S23. Detection of anti-PEG IgE in newborn serum samples by direct ELISA (Batch 1)

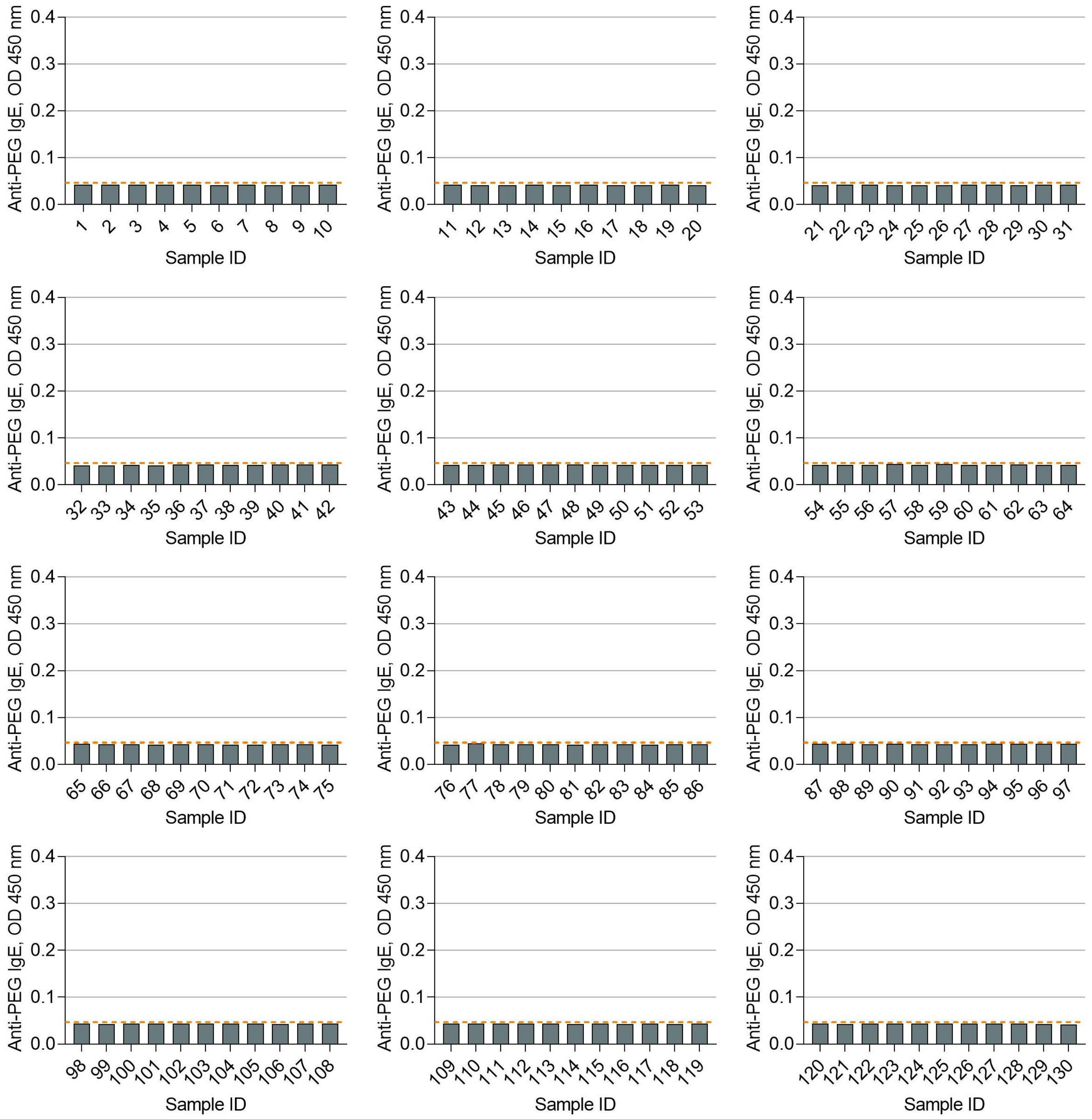


Dotted lines indicate the detection cutoff of anti-PEG IgE (see eMethods 3 for the establishment of detection cutoff of anti-PEG IgE).

**Figure S24. Detection of anti-PEG IgE in newborn serum samples by direct ELISA (Batch 2)**

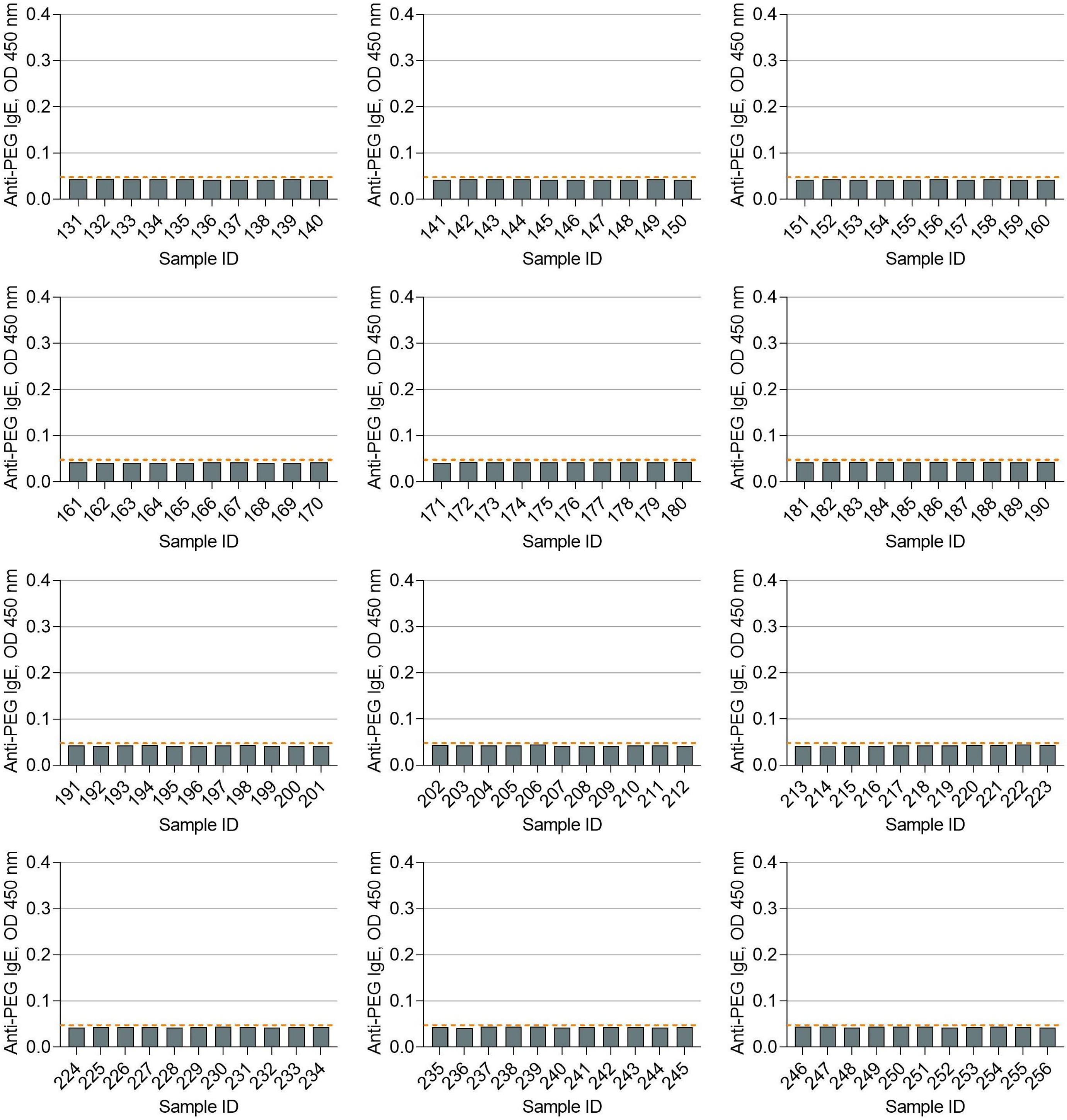


Dotted lines indicate the detection cutoff of anti-PEG IgE (see eMethods 3 for the establishment of detection cutoff of anti-PEG IgE).

**Table S4. Precision of competitive ELISA for confirmation of the PEG specificity of maternal and newborn anti-PEG IgG1-4 and IgM**

| | Intra-assay precision (CV%)[a] | | | |
|---|---|---|---|---|
| | Anti-PEG antibody standards without LIP | Anti-PEG antibody standards with LIP | Samples without LIP | Samples with LIP |
| **Pregnant women** | | | | |
| Anti-PEG IgG1 | 4.450 ± 3.277 | 5.298 ± 4.890 | 3.600 ± 3.091 | 3.617 ± 3.032 |
| Anti-PEG IgG2 | 2.829 ± 1.566 | 4.006 ± 3.028 | 2.908 ± 2.254 | 2.693 ± 2.233 |
| Anti-PEG IgG3 | 3.087 ± 1.620 | 5.827 ± 3.586 | 3.312 ± 3.028 | 2.979 ± 2.504 |
| Anti-PEG IgG4 | 2.920 ± 2.128 | 2.267 ± 1.683 | 1.799 ± 1.624 | 1.236 ± 1.401 |
| Anti-PEG IgM | 2.872 ± 2.146 | 1.995 ± 1.387 | 2.598 ± 2.160 | 2.771 ± 2.387 |
| **Newborns** | | | | |
| Anti-PEG IgG1 | 3.268 ± 3.264 | 4.555 ± 5.154 | 3.242 ± 2.613 | 2.453 ± 2.449 |
| Anti-PEG IgG2 | 3.571 ± 1.400 | 6.477 ± 2.463 | 3.323 ± 3.593 | 3.276 ± 2.490 |
| Anti-PEG IgG3 | 4.058 ± 1.919 | 5.319 ± 3.987 | 2.769 ± 2.401 | 2.557 ± 2.425 |
| Anti-PEG IgG4 | 4.220 ± 3.045 | 3.884 ± 2.679 | 2.377 ± 2.017 | 2.554 ± 2.060 |
| Anti-PEG IgM | 2.281 ± 0.900 | 3.390 ± 1.666 | 2.183 ± 1.908 | 1.962 ± 1.766 |

Abbreviations: LIP, PEGylated liposomal doxorubicin (competitors, see eMethods 4).

[a]Intra-assay precision was evaluated by calculating the coefficient of variation (CV% = (Standard deviation/Mean) × 100%) for all anti-PEG antibody standards without LIP, anti-PEG antibody standards with LIP, samples without LIP, and samples with LIP in one independent batch of competitive ELISA (see eMethods 5 for acceptance criteria). Data were presented as "mean ± standard deviation", with n = 9 for intra-assay precision (CV%) of anti-PEG antibody standards without LIP and with LIP in all competitive ELISA, n = 71, 118, 92, 35, 118, 74, 45, 84, 103, 138 for intra-assay precision (CV%) of samples without LIP and with LIP in competitive ELISA for the confirmation of PEG-specificity of maternal anti-PEG IgG1, newborn anti-PEG IgG1, maternal anti-PEG IgG2, newborn anti-PEG IgG2, maternal anti-PEG IgG3, newborn anti-PEG IgG3, maternal anti-PEG IgG4, newborn anti-PEG IgG4, maternal anti-PEG IgM and newborn anti-PEG IgM, respectively.

#### Figure S25. Establishment of specificity cutoffs of maternal anti-PEG IgG1-4 and IgM

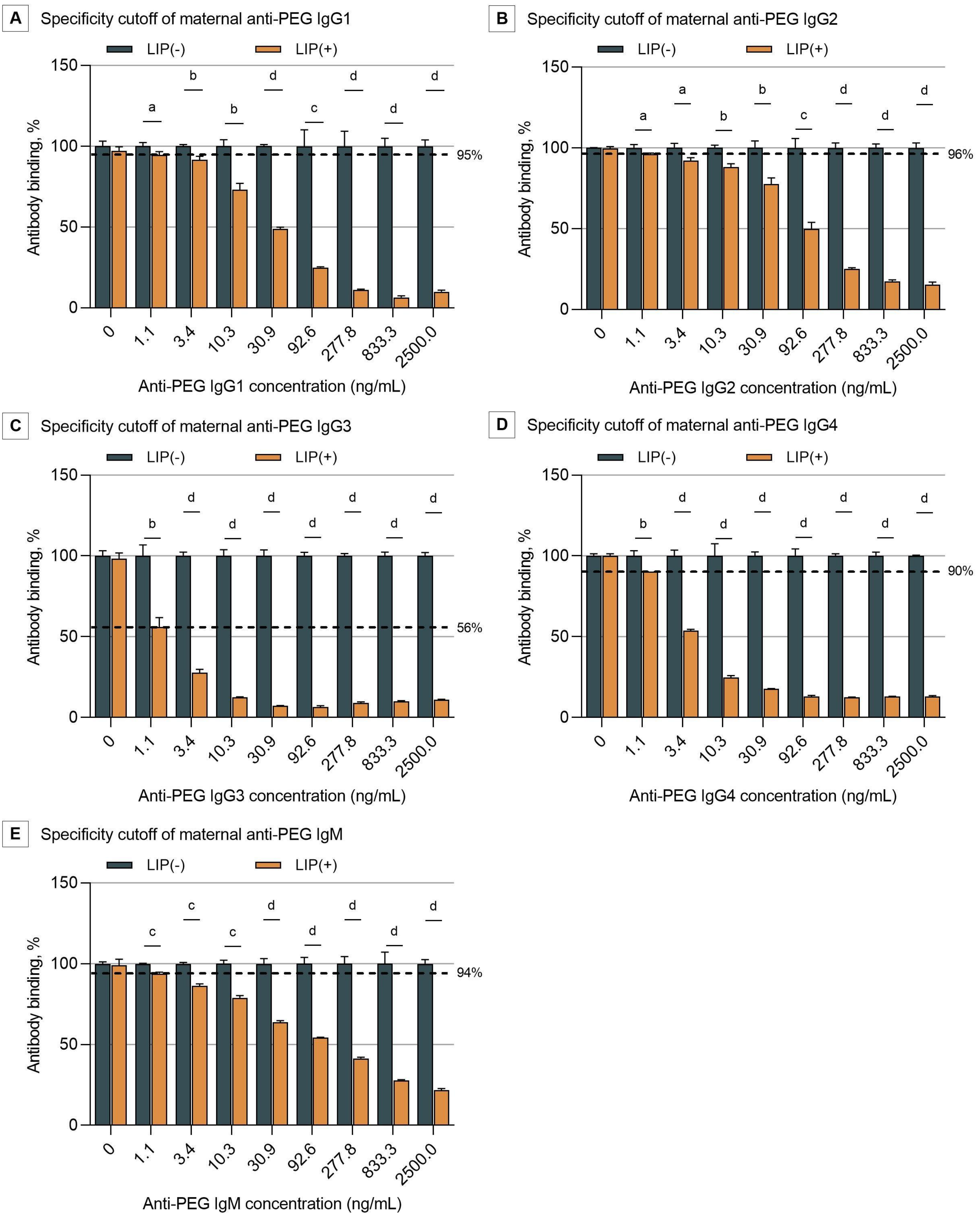


Abbreviations: LIP, PEGylated liposomal doxorubicin (competitors, see eMethods 4); LIP(-), anti-PEG antibody standards without the addition of LIP; LIP(+), anti-PEG antibody standards with the addition of LIP. Dotted lines indicate specificity cutoffs of maternal anti-PEG IgG1-4 and IgM (see eMethods 4 for establishment of specificity cutoffs of anti-PEG antibodies). All data were presented as "mean ± standard deviation" (n = 3), with differences between two groups analyzed using the two-tailed unpaired t-test. a, $P < 0.05$; b, $P < 0.01$; c, $P < 0.001$; d, $P < 0.0001$.

#### Figure S26. Establishment of specificity cutoffs of newborn anti-PEG IgG1-4 and IgM

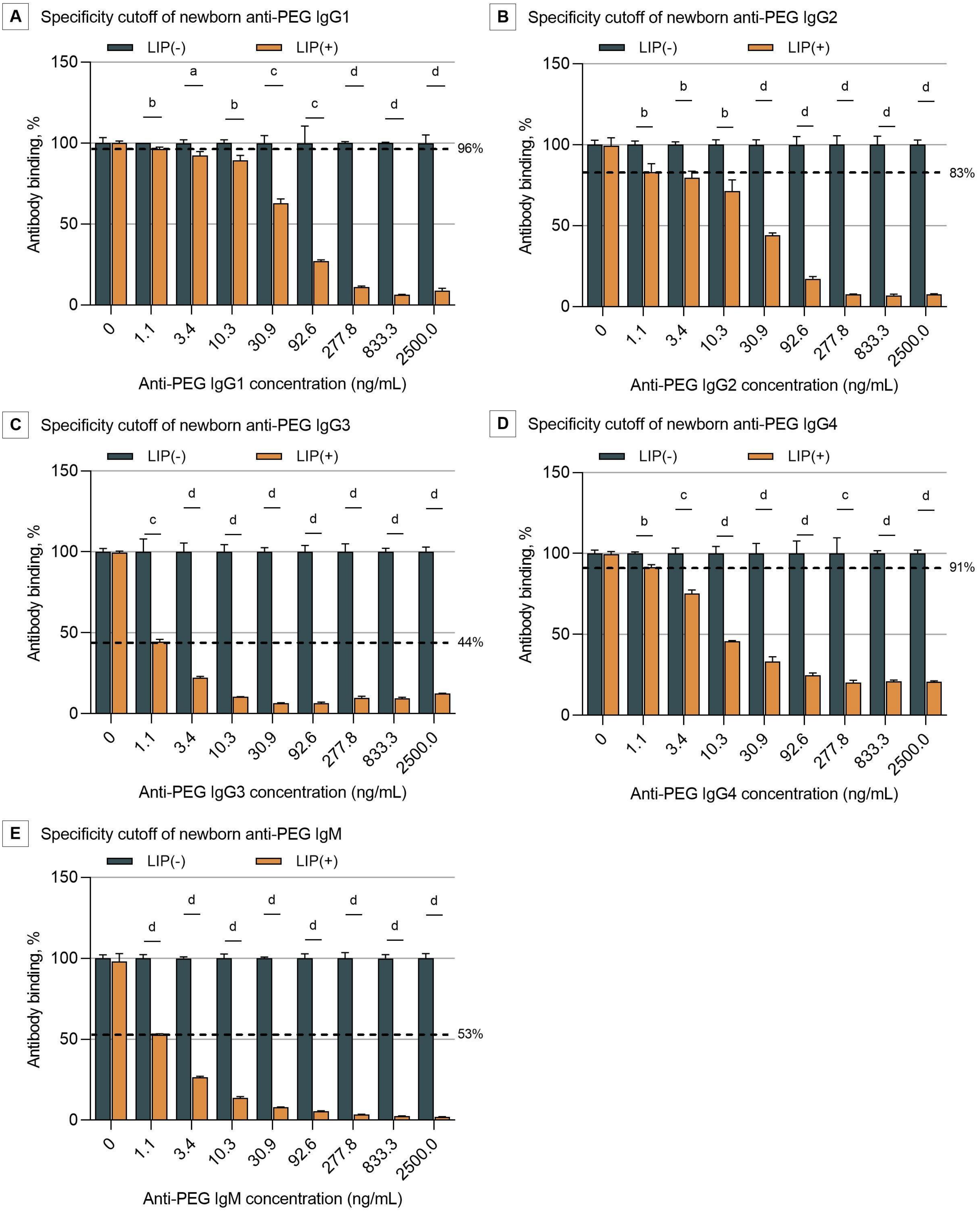


Abbreviations: LIP, PEGylated liposomal doxorubicin (competitors, see eMethods 4); LIP(-), anti-PEG antibody standards without the addition of LIP; LIP(+), anti-PEG antibody standards with the addition of LIP. Dotted lines indicate specificity cutoffs of maternal anti-PEG IgG1-4 and IgM (see eMethods 4 for establishment of specificity cutoffs of anti-PEG antibodies). All data were presented as "mean ± standard deviation" (n = 3), with differences between two groups analyzed using the two-tailed unpaired t-test. a, $P < 0.05$; b, $P < 0.01$; c, $P < 0.001$; d, $P < 0.0001$.

**Figure S27. Confirmation of the PEG specificity of anti-PEG IgG1 in maternal serum samples by competitive ELISA**

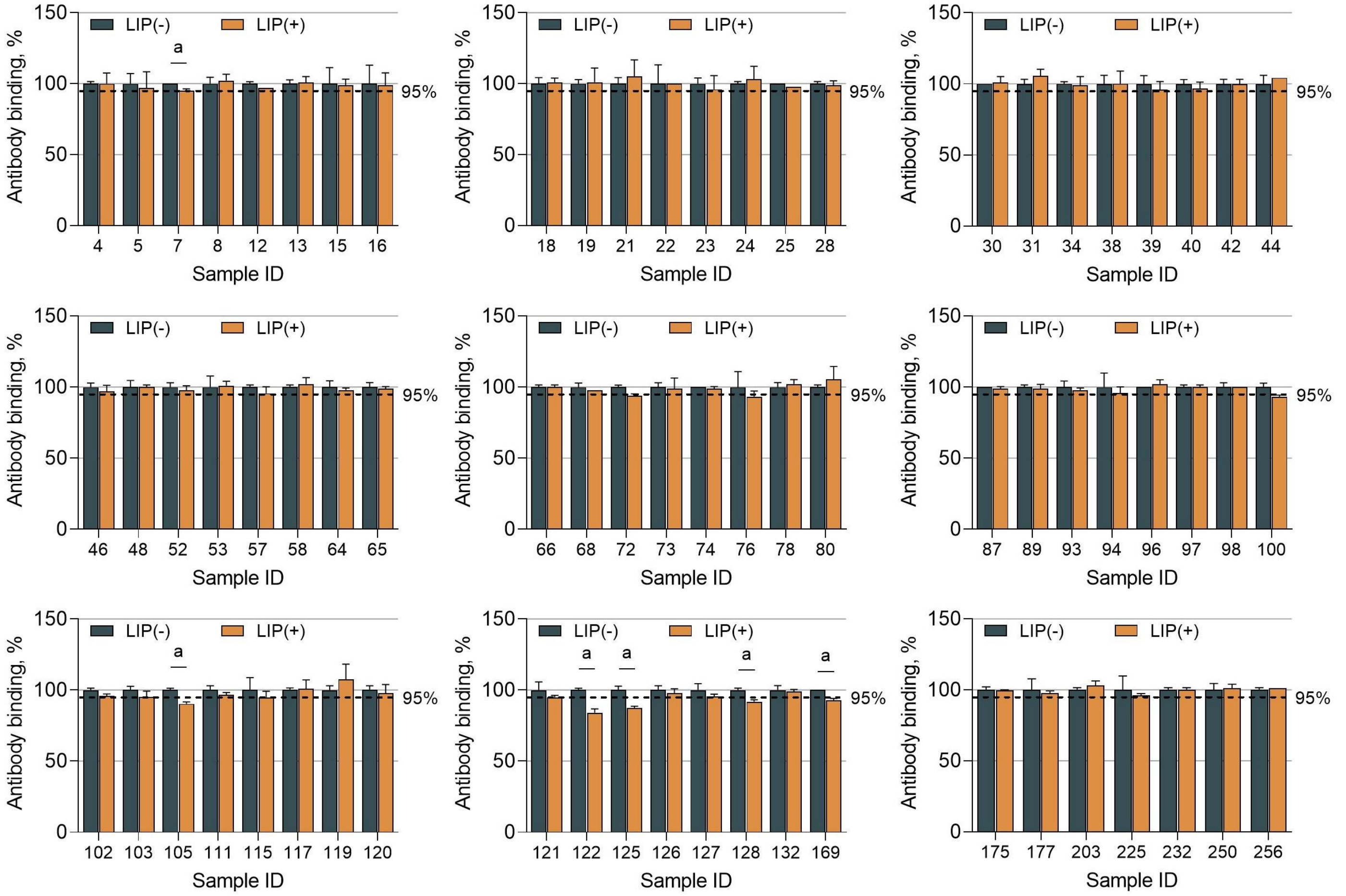

Abbreviations: LIP, PEGylated liposomal doxorubicin (competitors, see eMethods 4); LIP(-), maternal serum samples without the addition of LIP; LIP(+), maternal serum samples with the addition of LIP. Dotted lines indicate specificity cutoffs of maternal anti-PEG IgG1. All data were presented as “mean ± standard deviation” (n = 2), with differences between two groups analyzed using the two-tailed unpaired t-test. a, $P < 0.05$.

**Figure S28. Confirmation of the PEG specificity of anti-PEG IgG1 in newborn serum samples by competitive ELISA**

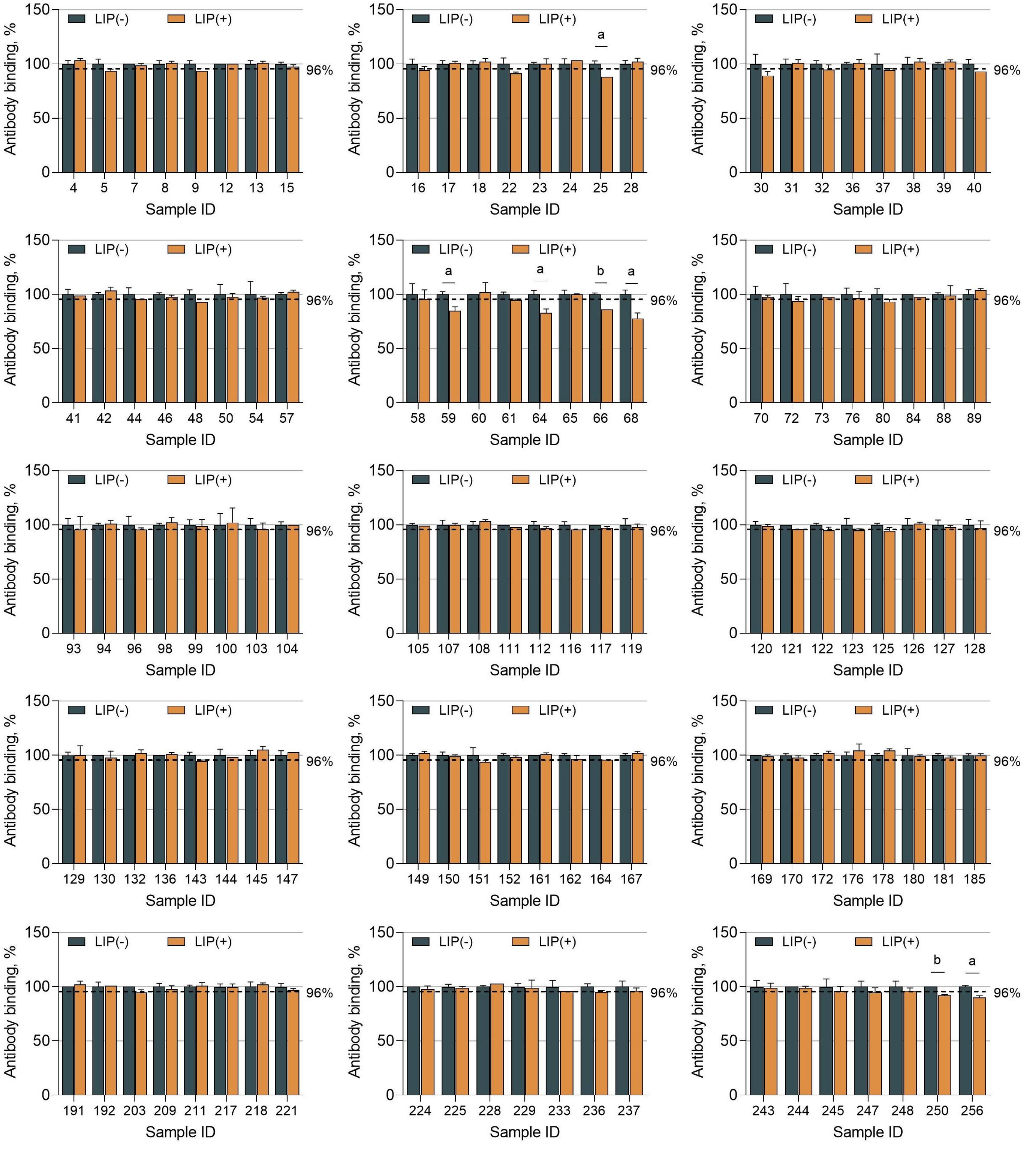


Abbreviations: LIP, PEGylated liposomal doxorubicin (competitors, see eMethods 4); LIP(-), newborn serum samples without the addition of LIP; LIP(+), newborn serum samples with the addition of LIP. Dotted lines indicate specificity cutoffs of neonatal anti-PEG IgG1. All data were presented as "mean ± standard deviation" (n = 2), with differences between two groups analyzed using the two-tailed unpaired t-test. a, $P < 0.05$; b, $P < 0.01$.

**Figure S29. Confirmation of the PEG specificity of anti-PEG IgG2 in maternal serum samples by competitive ELISA**

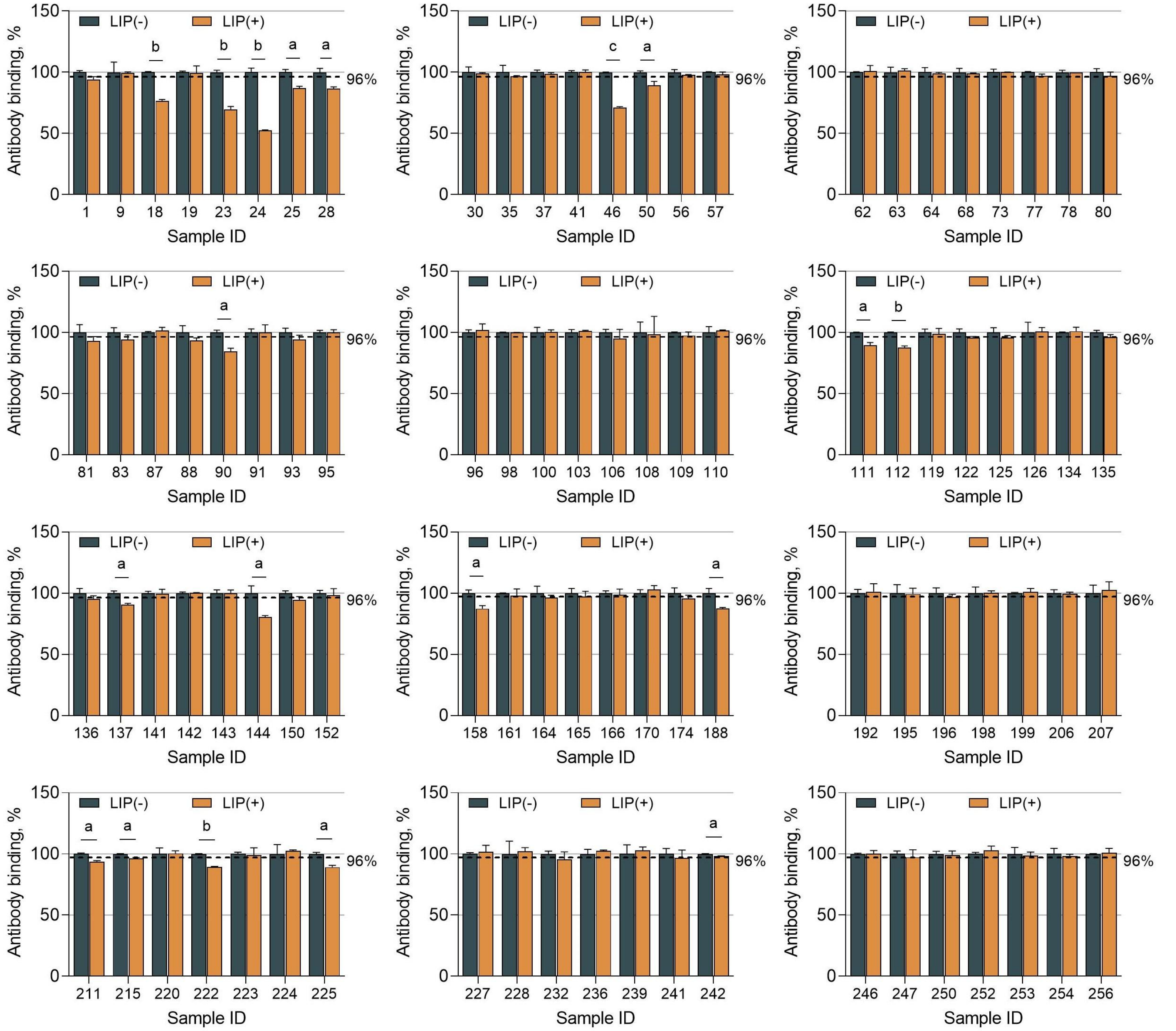


Abbreviations: LIP, PEGylated liposomal doxorubicin (competitors, see eMethods 4); LIP(-), maternal serum samples without the addition of LIP; LIP(+), maternal serum samples with the addition of LIP. Dotted lines indicate specificity cutoffs of maternal anti-PEG IgG2. All data were presented as "mean ± standard deviation" (n = 2), with differences between two groups analyzed using the two-tailed unpaired t-test. a, $P < 0.05$; b, $P < 0.01$; c, $P < 0.001$.

**Figure S30. Confirmation of the PEG specificity of anti-PEG IgG2 in newborn serum samples by competitive ELISA**

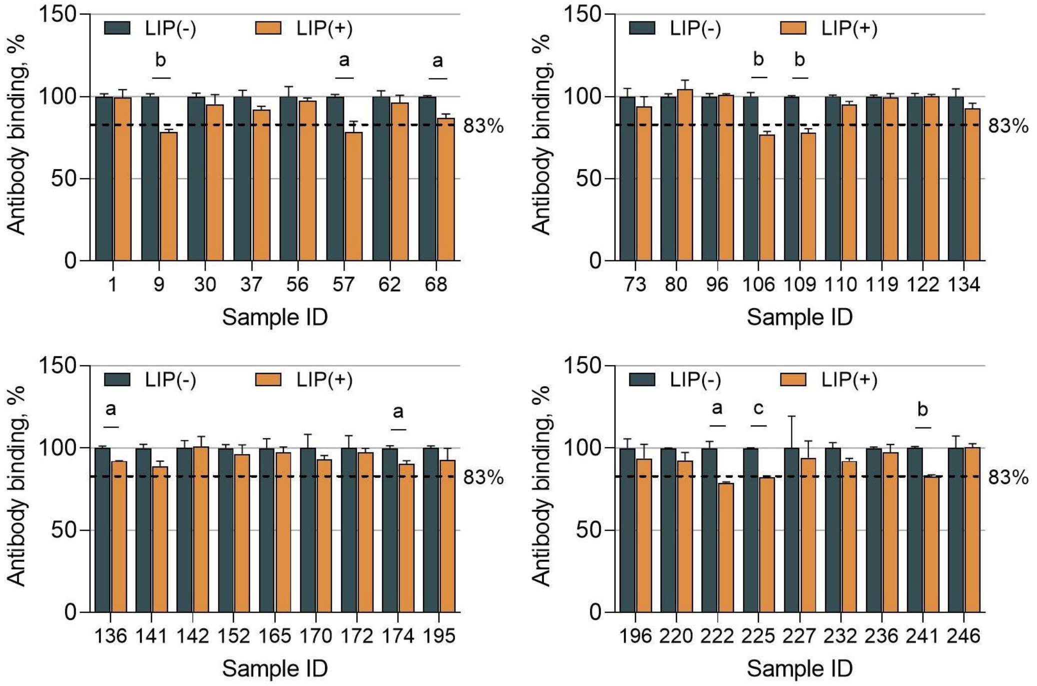


Abbreviations: LIP, PEGylated liposomal doxorubicin (competitors, see eMethods 4); LIP(-), newborn serum samples without the addition of LIP; LIP(+), newborn serum samples with the addition of LIP. Dotted lines indicate specificity cutoffs of neonatal anti-PEG IgG2. All data were presented as "mean ± standard deviation" (n = 2), with differences between two groups analyzed using the two-tailed unpaired t-test. a, $P < 0.05$; b, $P < 0.01$; c, $P < 0.001$.

**Figure S31. Confirmation of the PEG specificity of anti-PEG IgG3 in maternal serum samples by competitive ELISA**

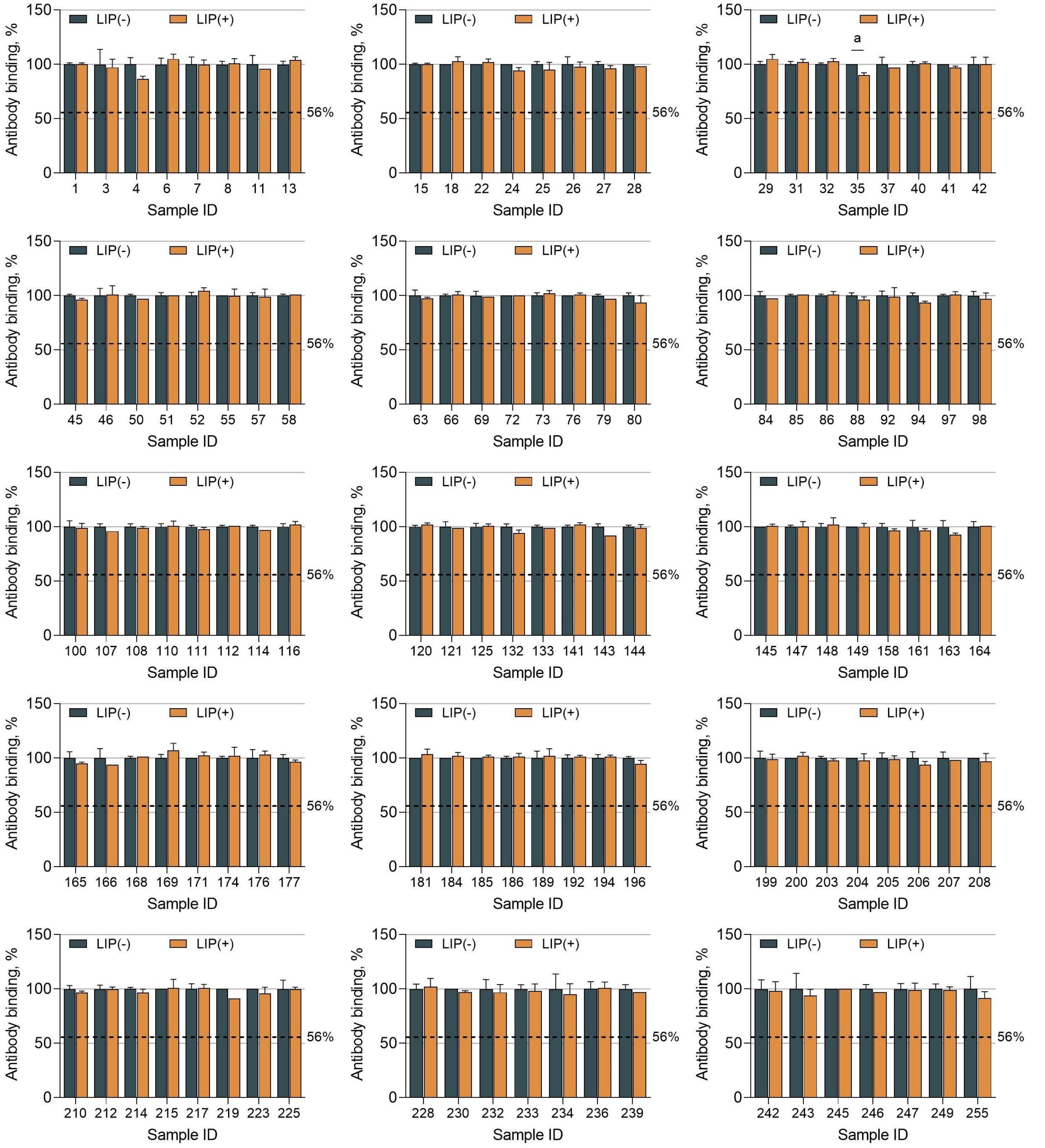


Abbreviations: LIP, PEGylated liposomal doxorubicin (competitors, see eMethods 4); LIP(-), newborn serum samples without the addition of LIP; LIP(+), newborn serum samples with the addition of LIP. Dotted lines indicate specificity cutoffs of maternal anti-PEG IgG3. All data were presented as “mean ± standard deviation” (n = 2), with differences between two groups analyzed using the two-tailed unpaired t-test. a, $P < 0.05$.

#### Figure S32. Confirmation of the PEG specificity of anti-PEG IgG3 in newborn serum samples by competitive ELISA

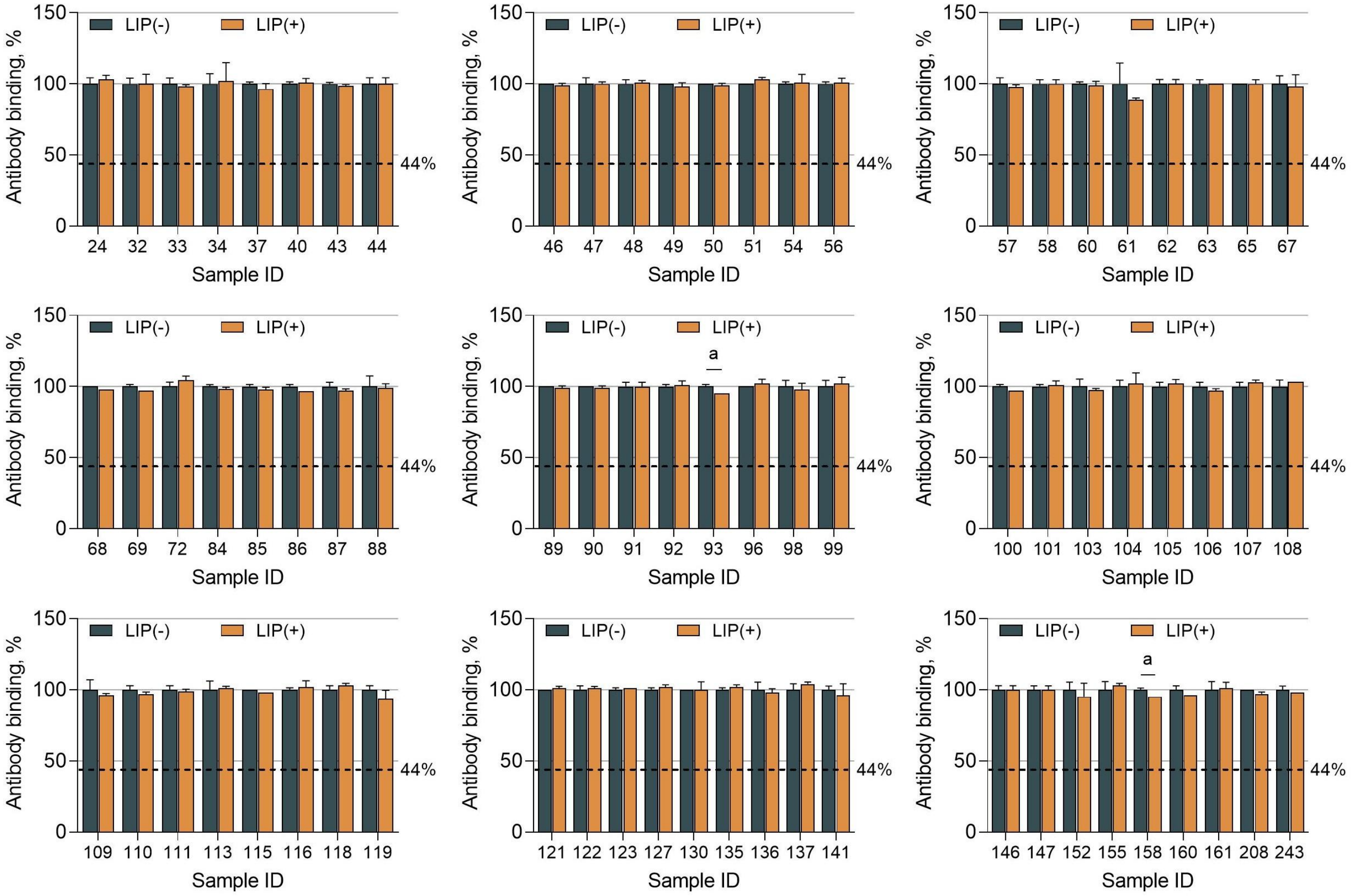


Abbreviations: LIP, PEGylated liposomal doxorubicin (competitors, see eMethods 4); LIP(-), newborn serum samples without the addition of LIP; LIP(+), newborn serum samples with the addition of LIP. Dotted lines indicate specificity cutoffs of neonatal anti-PEG IgG3. All data were presented as "mean ± standard deviation" (n = 2), with differences between two groups analyzed using the two-tailed unpaired t-test. a, $P < 0.05$.

**Figure S33. Confirmation of the PEG specificity of anti-PEG IgG4 in maternal serum samples by competitive ELISA**

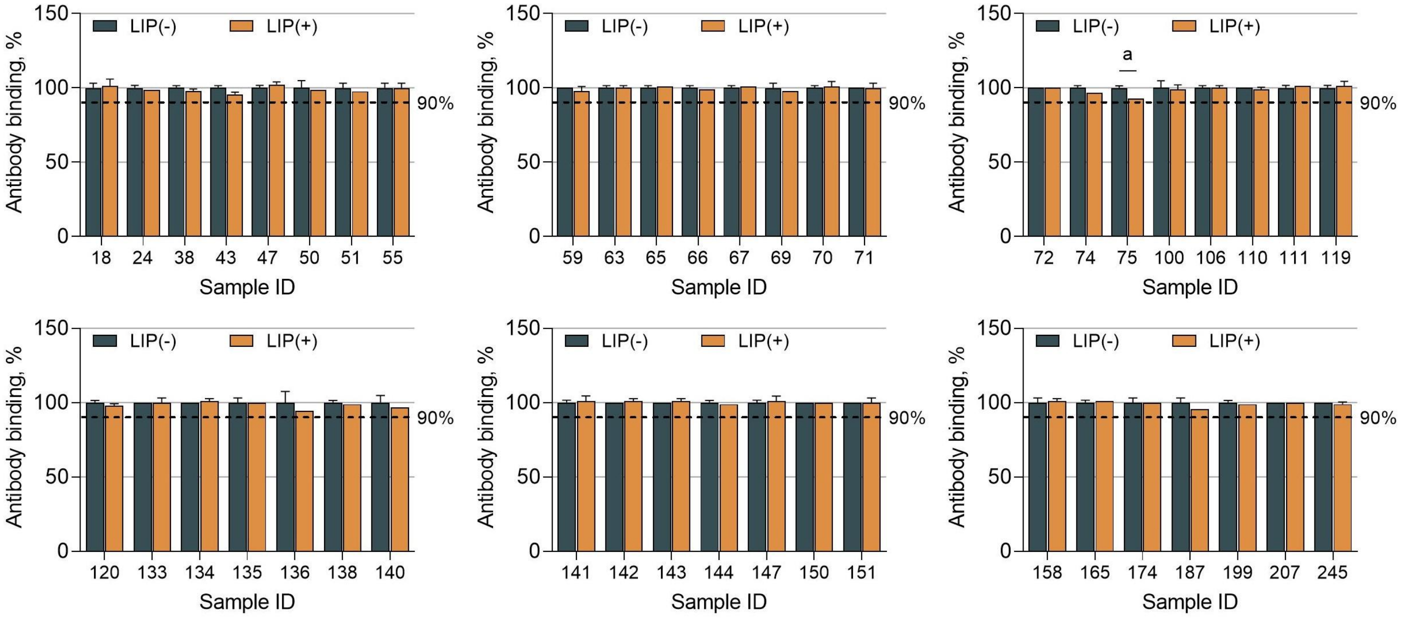


Abbreviations: LIP, PEGylated liposomal doxorubicin (competitors, see eMethods 4); LIP(-), newborn serum samples without the addition of LIP; LIP(+), newborn serum samples with the addition of LIP. Dotted lines indicate specificity cutoffs of maternal anti-PEG IgG4. All data were presented as "mean ± standard deviation" (n = 2), with differences between two groups analyzed using the two-tailed unpaired t-test. a, $P < 0.05$.

**Figure S34. Confirmation of the PEG specificity of anti-PEG IgG4 in newborn serum samples by competitive ELISA**

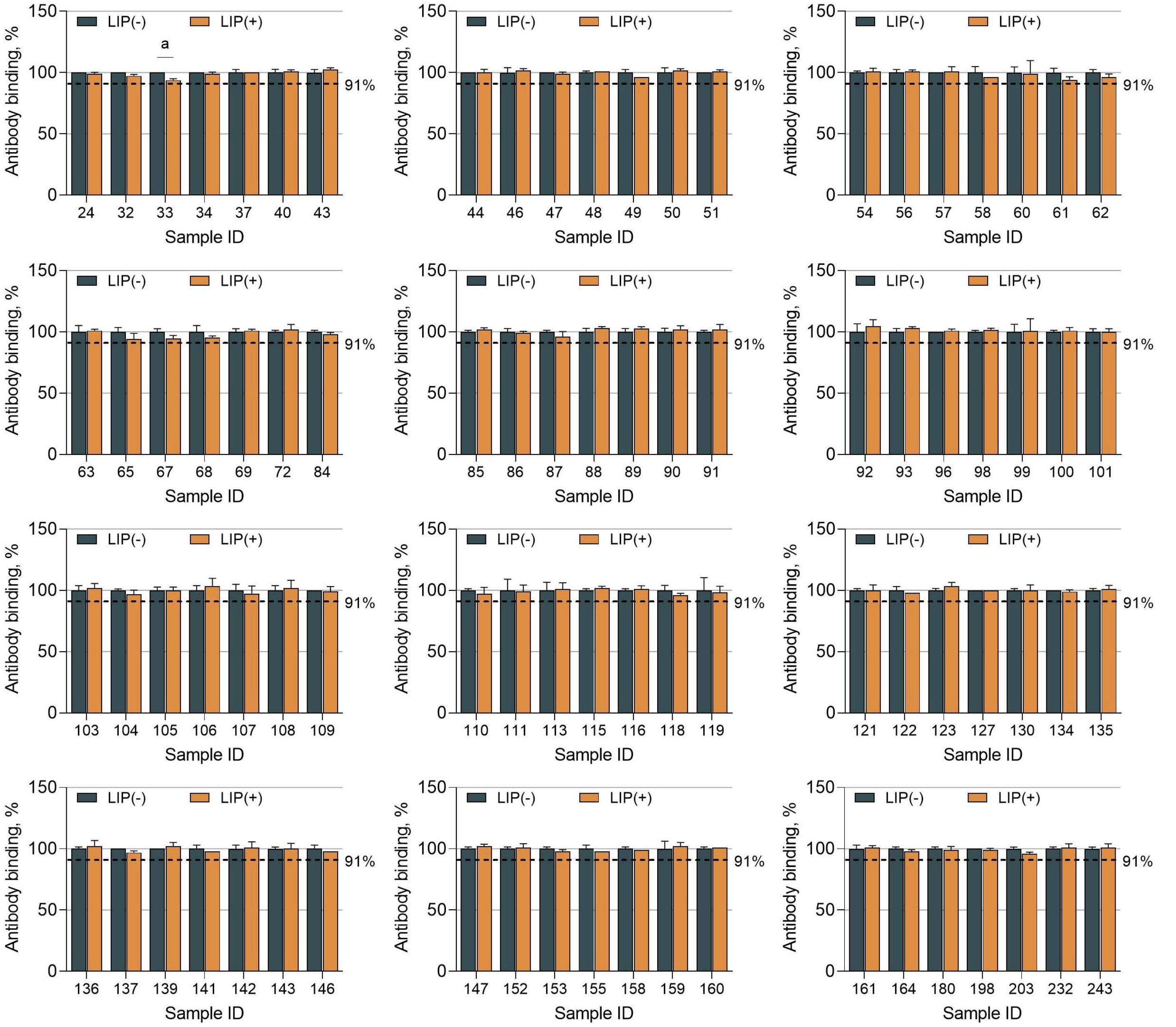


Abbreviations: LIP, PEGylated liposomal doxorubicin (competitors, see eMethods 4); LIP(-), newborn serum samples without the addition of LIP; LIP(+), newborn serum samples with the addition of LIP. Dotted lines indicate specificity cutoffs of neonatal anti-PEG IgG4. All data were presented as "mean ± standard deviation" (n = 2), with differences between two groups analyzed using the two-tailed unpaired t-test. a, $P < 0.05$.

**Figure S35. Confirmation of the PEG specificity of anti-PEG IgM in maternal serum samples by competitive ELISA**

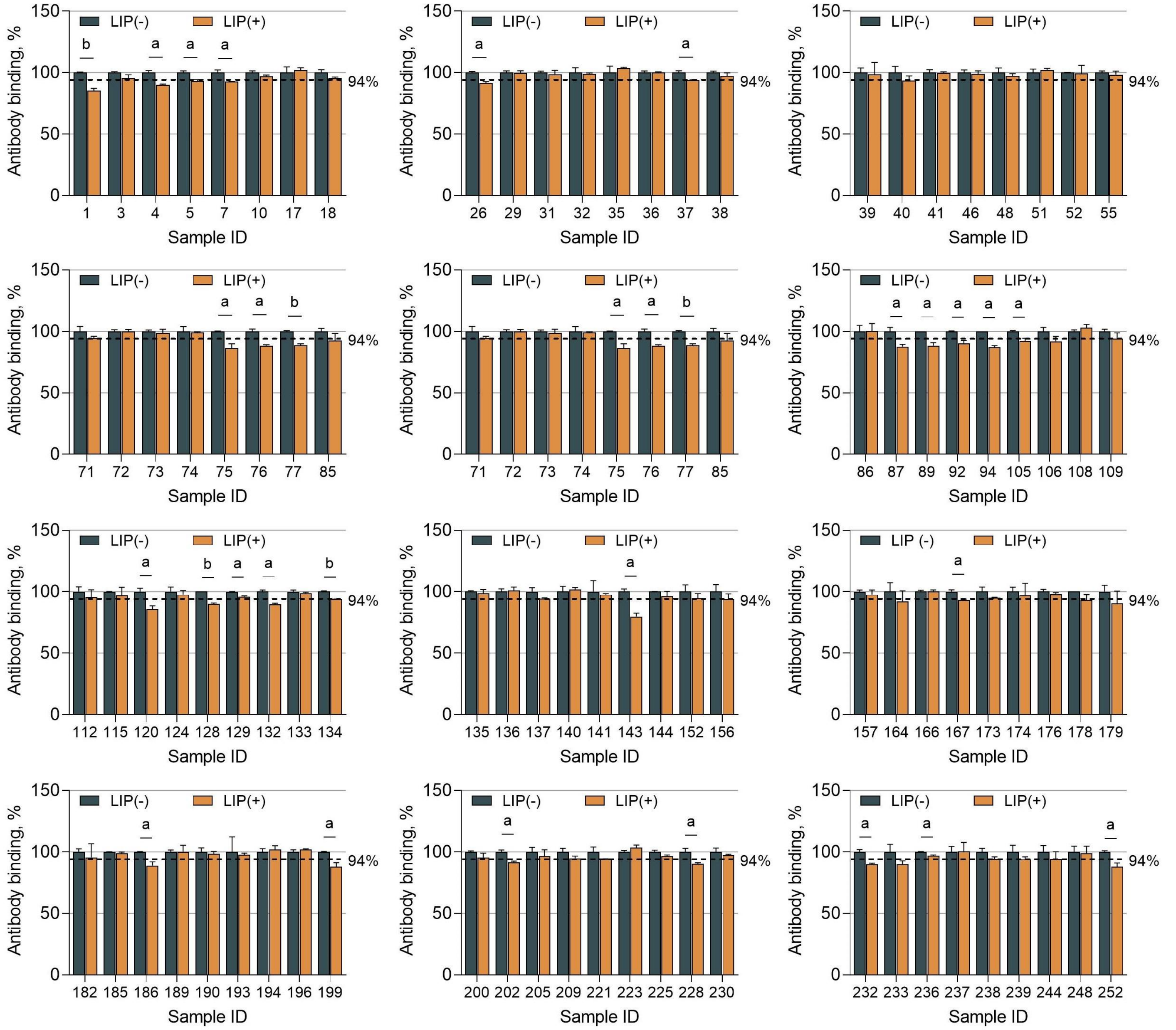


Abbreviations: LIP, PEGylated liposomal doxorubicin (competitors, see eMethods 4); LIP(-), newborn serum samples without the addition of LIP; LIP(+), newborn serum samples with the addition of LIP. Dotted lines indicate specificity cutoffs of maternal anti-PEG IgM. All data were presented as "mean ± standard deviation" (n = 2), with differences between two groups analyzed using the two-tailed unpaired t-test. a, $P < 0.05$; b, $P < 0.01$.

#### Figure S36. Confirmation of the PEG specificity of anti-PEG IgM in newborn serum samples by competitive ELISA

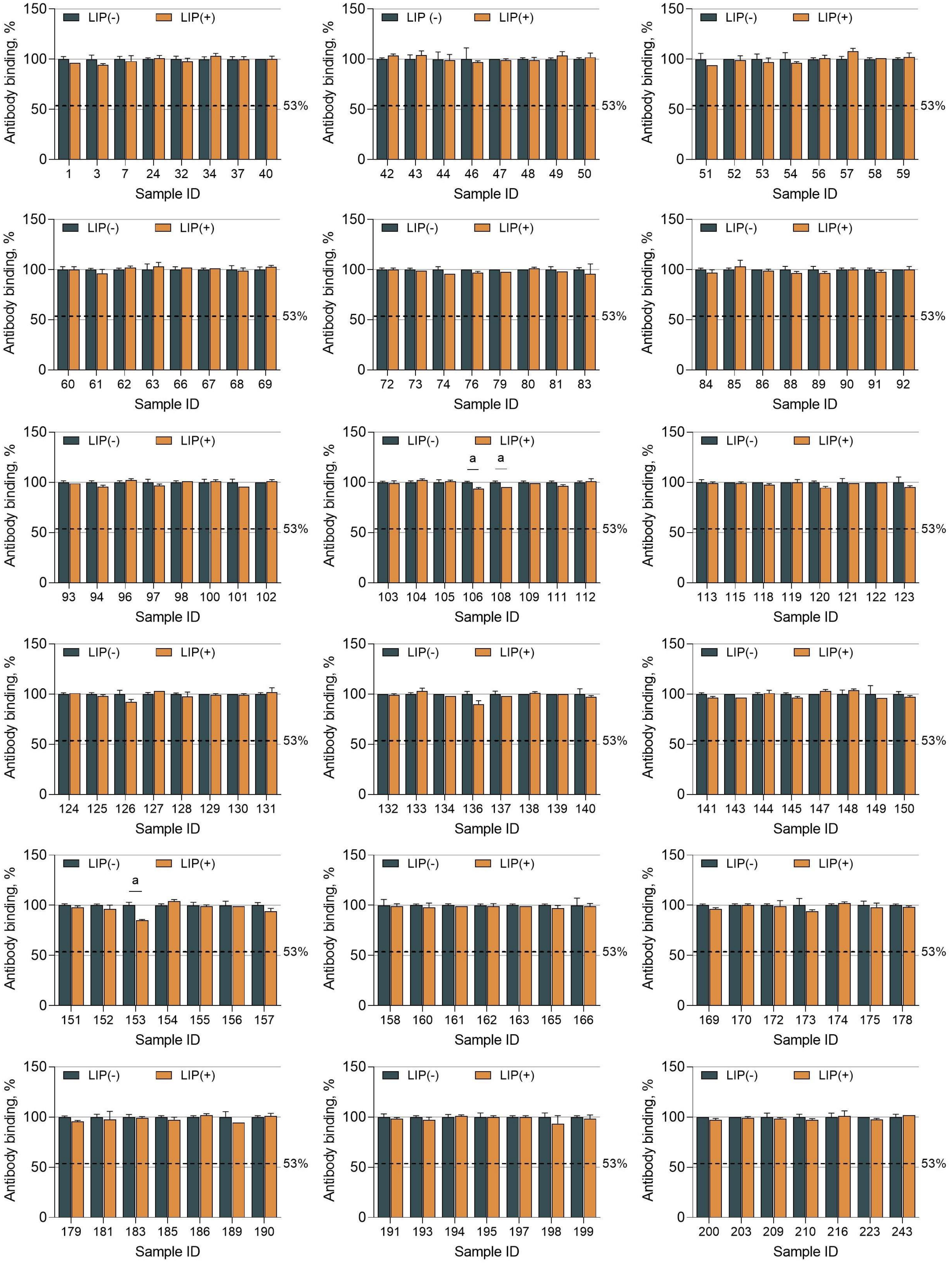


Abbreviations: LIP, PEGylated liposomal doxorubicin (competitors, see eMethods 4); LIP(-), newborn serum samples without the addition of LIP; LIP(+), newborn serum samples with the addition of LIP. Dotted lines indicate specificity cutoffs of neonatal anti-PEG IgM. All data were presented as “mean ± standard deviation” (n = 2), with differences between two groups analyzed using the two-tailed unpaired t-test. a, $P < 0.05$.

**Figure S37. Standard curves of direct ELISA for detecting anti-PEG IgG1 in maternal and newborn serum samples[a]**

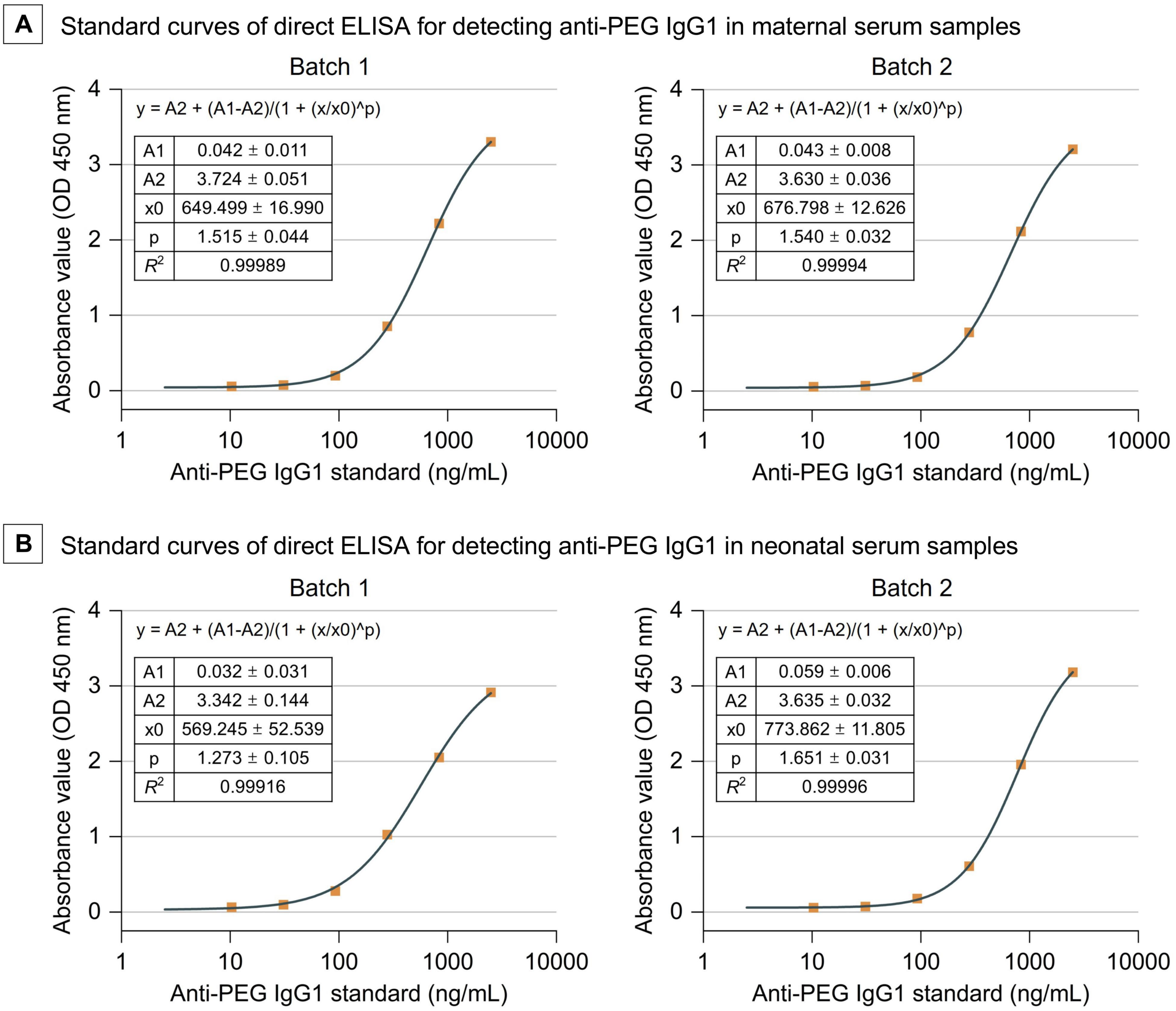


Abbreviations: $R^2$, coefficient of determination.

[a]Standard curves were constructed by plotting the average absorbance values (OD450 nm) and corresponding antibody concentrations with Four Parameter Logistic (4PL) curve fit using Origin 2021 software. Serial dilutions of human anti-PEG IgG1 standards (0, 10.3, 30.9, 92.6, 277.8, 833.3, and 2500.0 ng/mL) were included in each direct ELISA for two independent batches.

**Figure S38. Standard curves of direct ELISA for detecting anti-PEG IgG2 in maternal and newborn serum samples[a]**

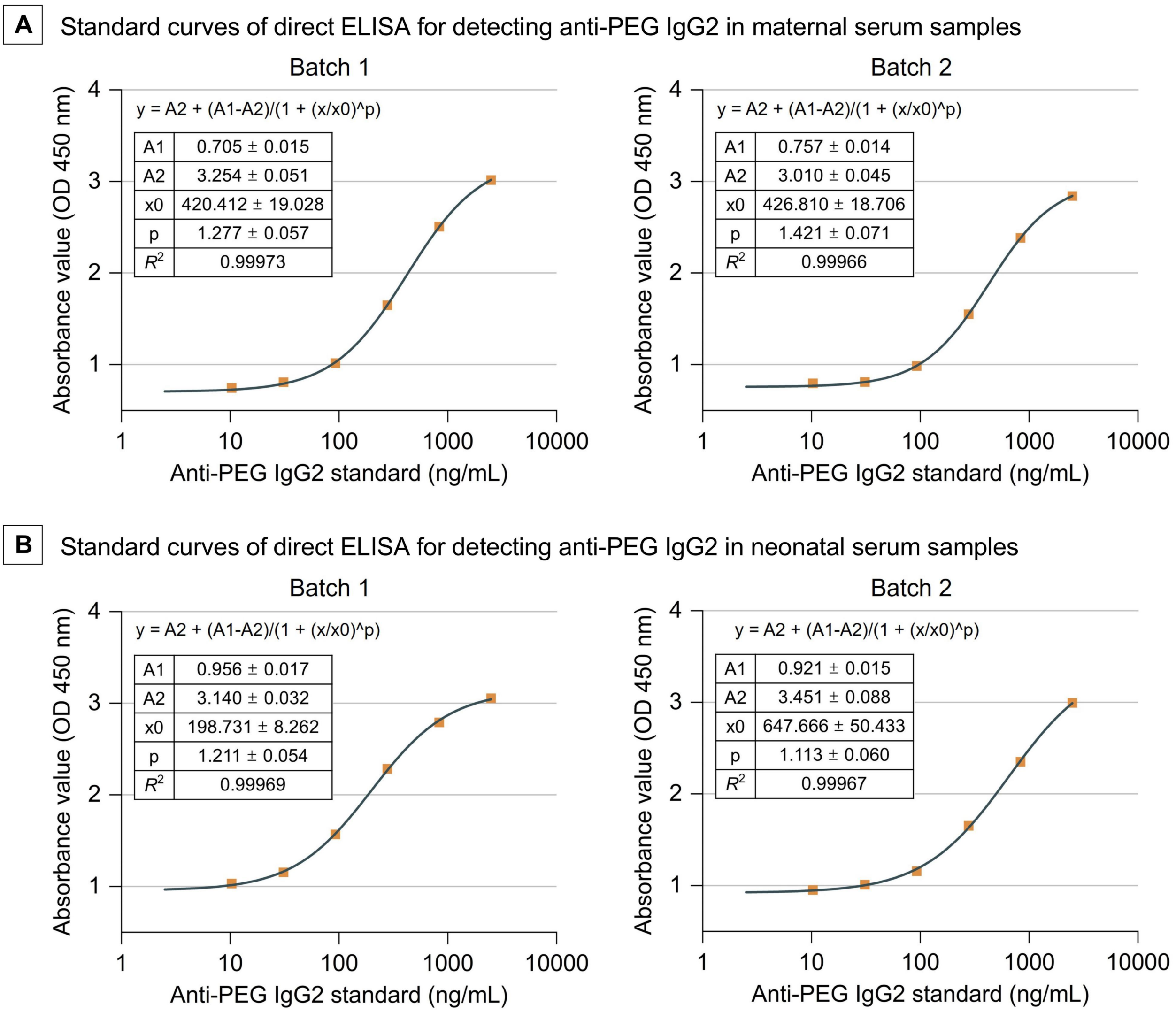


Abbreviations: $R^2$, coefficient of determination.

[a]Standard curves were constructed by plotting the average absorbance values (OD450 nm) and corresponding antibody concentrations with Four Parameter Logistic (4PL) curve fit using Origin 2021 software. Serial dilutions of human anti-PEG IgG2 standards (0, 10.3, 30.9, 92.6, 277.8, 833.3, and 2500.0 ng/mL) were included in each direct ELISA for two independent batches.

**Figure S39. Standard curves of direct ELISA for detecting anti-PEG IgG3 in maternal and newborn serum samples[a]**

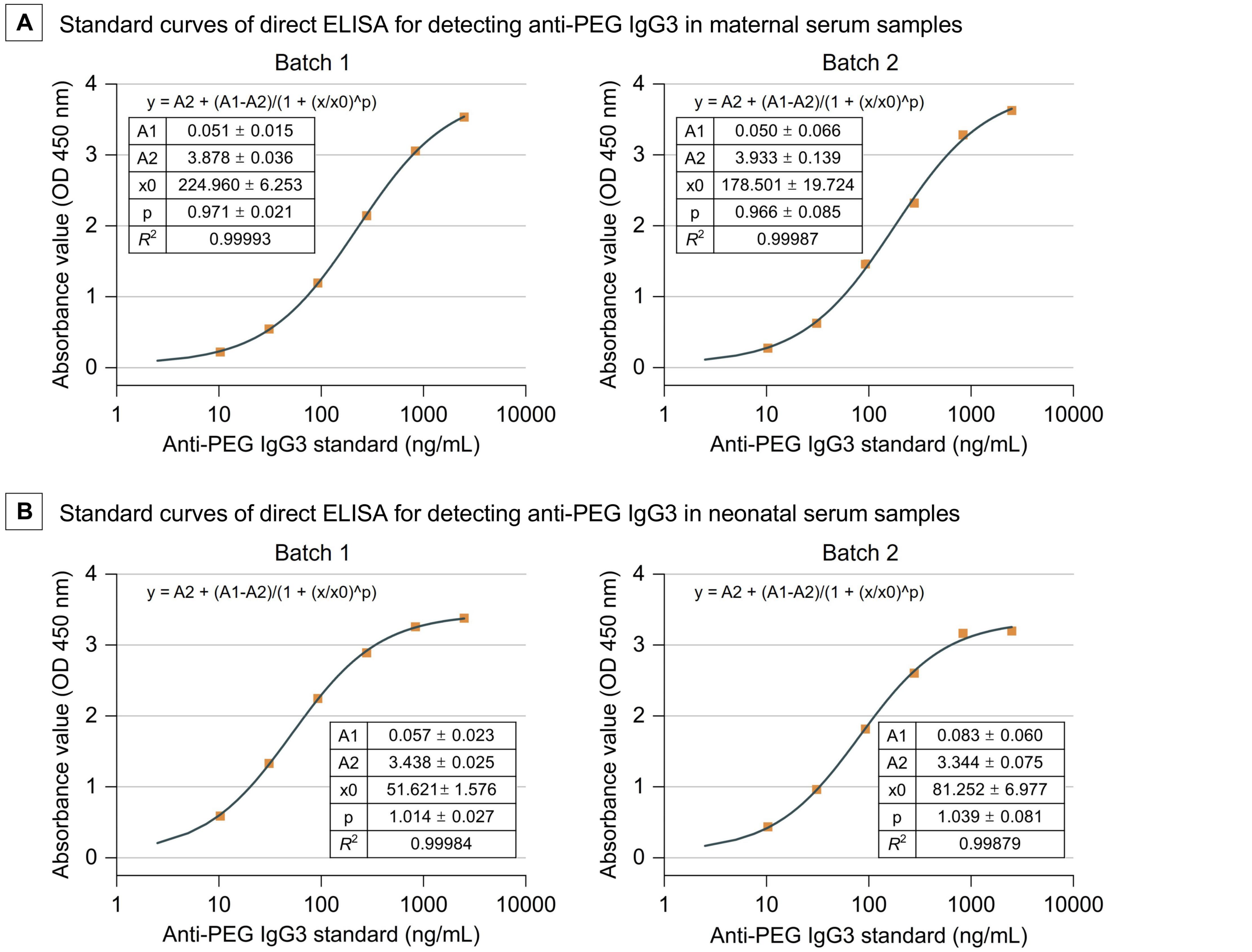


Abbreviations: $R^2$, coefficient of determination.

[a]Standard curves were constructed by plotting the average absorbance values (OD450 nm) and corresponding antibody concentrations with Four Parameter Logistic (4PL) curve fit using Origin 2021 software. Serial dilutions of human anti-PEG IgG3 standards (0, 10.3, 30.9, 92.6, 277.8, 833.3, and 2500.0 ng/mL) were included in each direct ELISA for two independent batches.

#### Figure S40. Standard curves of direct ELISA for detecting anti-PEG IgG4 in maternal and newborn serum samples[a]

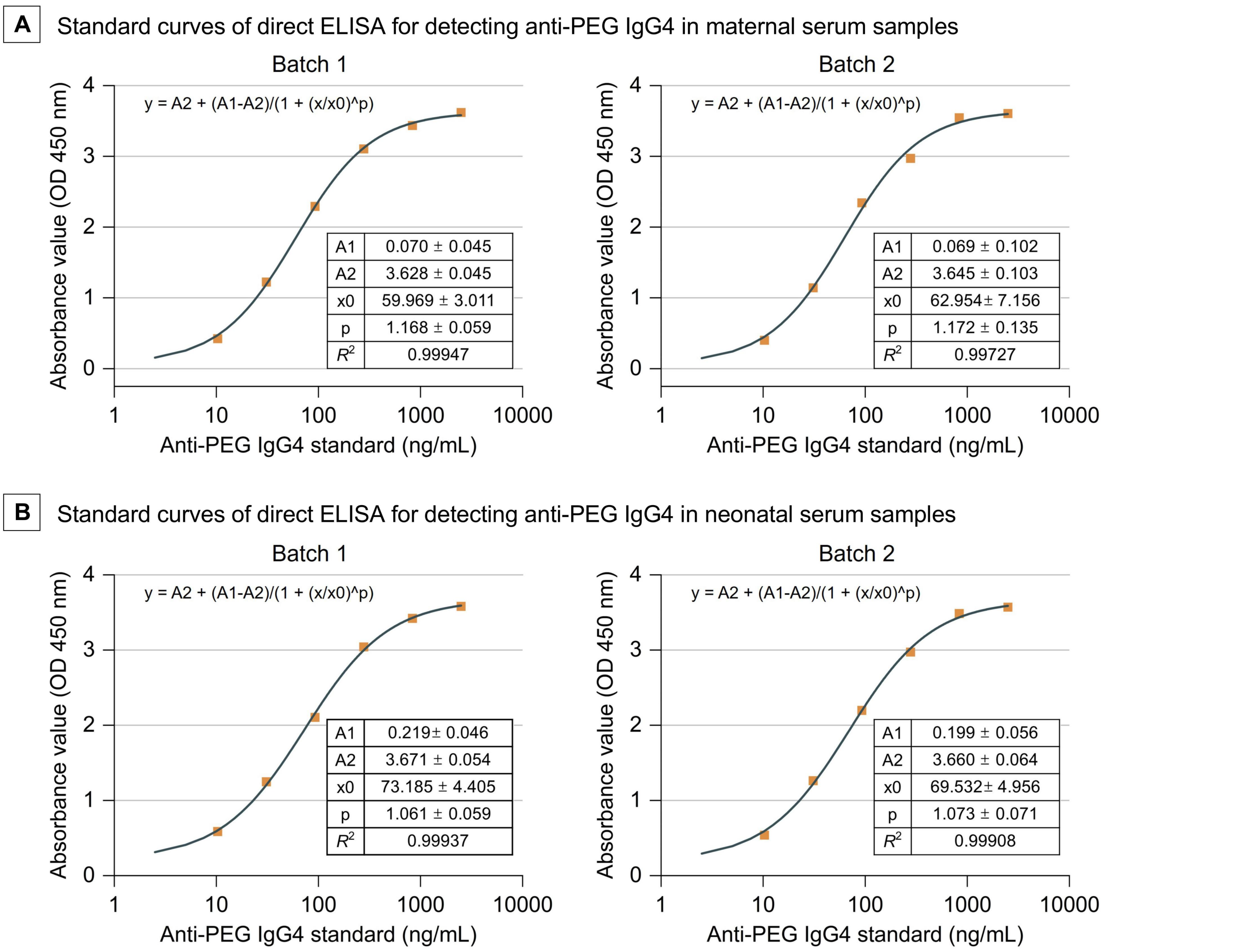


Abbreviations: $R^2$, coefficient of determination.

[a]Standard curves were constructed by plotting the average absorbance values (OD450 nm) and corresponding antibody concentrations with Four Parameter Logistic (4PL) curve fit using Origin 2021 software. Serial dilutions of human anti-PEG IgG4 standards (0, 10.3, 30.9, 92.6, 277.8, 833.3, and 2500.0 ng/mL) were included in each direct ELISA for two independent batches.

**Figure S41. Standard curves of direct ELISA for detecting anti-PEG IgM in maternal and newborn serum samples[a]**

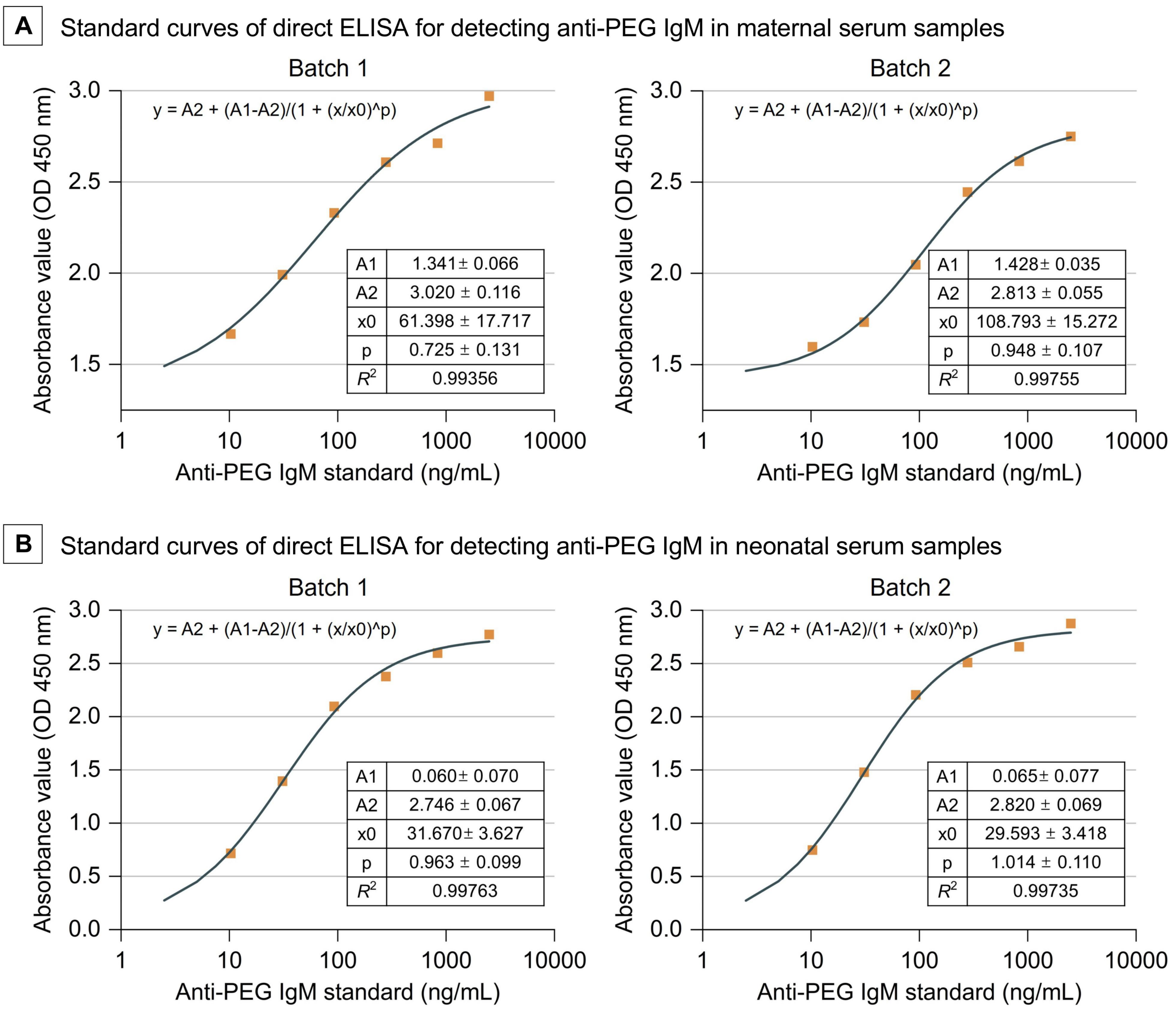


Abbreviations: $R^2$, coefficient of determination.
[a]Standard curves were constructed by plotting the average absorbance values (OD450 nm) and corresponding antibody concentrations with Four Parameter Logistic (4PL) curve fit using Origin 2021 software. Serial dilutions of human anti-PEG IgM standards (0, 10.3, 30.9, 92.6, 277.8, 833.3, and 2500.0 ng/mL) were included in each direct ELISA for two independent batches.

**Figure S42. Standard curves of direct ELISA for detecting anti-PEG IgE in maternal and newborn serum samples[a]**

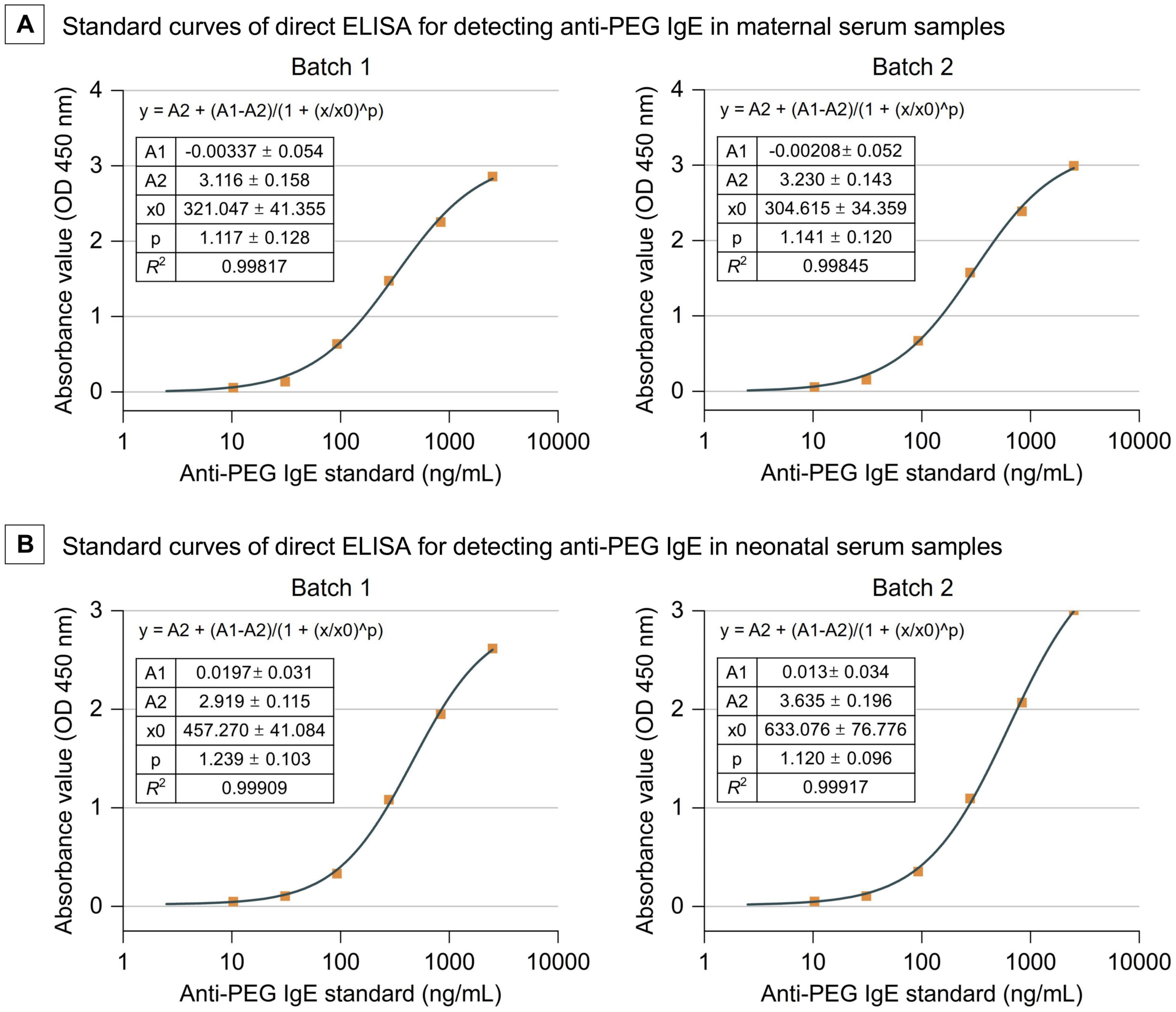


Abbreviations: $R^2$, coefficient of determination.

[a]Standard curves were constructed by plotting the average absorbance values (OD450 nm) and corresponding antibody concentrations with Four Parameter Logistic (4PL) curve fit using Origin 2021 software. Serial dilutions of human anti-PEG IgE standards (0, 10.3, 30.9, 92.6, 277.8, 833.3, and 2500.0 ng/mL) were included in each direct ELISA for two independent batches.

#### Table S5. Anti-PEG IgG1 levels in seropositive pregnant women

| Sample ID | Maternal anti-PEG IgG1 levels (ng/mL) |
|---|---|
| 7 | 259.1147 |
| 105 | 513.8989 |
| 122 | 288.6406 |
| 125 | 194.3535 |
| 128 | 183.7449 |
| 169 | 330.1449 |

#### Table S6. Anti-PEG IgG1 levels in seropositive newborns

| Sample ID | Newborn anti-PEG IgG1 levels (ng/mL) |
|---|---|
| 25 | 207.9177 |
| 59 | 163.7862 |
| 64 | 120.4021 |
| 66 | 524.5799 |
| 68 | 133.9695 |
| 250 | 1513.9751 |
| 256 | 481.4903 |

**Table S7. Anti-PEG IgG2 levels in seropositive pregnant women**

| Sample ID | Maternal anti-PEG IgG2 levels (ng/mL) |
|---|---|
| 18 | 868.0852 |
| 23 | 1412.0005 |
| 24 | 120.2819 |
| 25 | 102.2656 |
| 28 | 75.536 |
| 46 | 85.3274 |
| 50 | 999.4361 |
| 90 | 734.603 |
| 111 | 843.6952 |
| 112 | 476.9167 |
| 137 | 762.0953 |
| 144 | 292.0242 |
| 158 | 671.5611 |
| 188 | 1727.9897 |
| 211 | 1155.2939 |
| 215 | 172.0243 |
| 222 | 1337.3718 |
| 225 | 2604.887 |

**Table S8. Anti-PEG IgG2 levels in seropositive newborns**

| Sample ID | Newborn anti-PEG IgG2 levels (ng/mL) |
| --- | --- |
| 9 | 275.9202 |
| 57 | 527.0678 |
| 106 | 474.5477 |
| 109 | 1069.6205 |
| 222 | 275.8009 |
| 225 | 100.2442 |
| 241 | 336.5249 |

**Table S9. Anti-PEG IgM levels in seropositive pregnant women**

| Sample ID | Maternal anti-PEG IgM levels (ng/mL) |
|---|---|
| 1 | 153.8652 |
| 4 | 188.3983 |
| 5 | 93.8578 |
| 7 | 316.8341 |
| 26 | 95.7684 |
| 37 | 358.0414 |
| 67 | 177.5309 |
| 68 | 57.7931 |
| 75 | 96.4906 |
| 76 | 431.0382 |
| 77 | 225.1164 |
| 87 | 258.4069 |
| 89 | 55.4326 |
| 92 | 23649.1374 |
| 94 | 140.125 |
| 105 | 207.1515 |
| 120 | 172.6121 |
| 128 | 142.561 |
| 132 | 61.5203 |
| 134 | 624.2215 |
| 143 | 100.1753 |
| 167 | 89.1675 |
| 186 | 386.5907 |
| 199 | 308.9327 |
| 202 | 310.2522 |
| 228 | 1024.5648 |
| 232 | 58.2421 |
| 252 | 146.6371 |

**Table S10. Univariable logistic regression analysis for the prevalence of pre-existing total anti-PEG antibodies in pregnant women (n = 256)**

| Variable | OR (95% CI) | *P* value |
|---|---|---|
| **Maternal age** | 0.887 (0.820-0.959) | **0.003** |
| **BMI**[a] | 1.051 (0.954-1.157) | 0.314 |
| **Education** | | |
| Vocational degree or below | 1 [Reference] | NA |
| Associate's degree | 1.056 (0.431-2.589) | 0.905 |
| Bachelor's degree | 0.717 (0.310-1.660) | 0.437 |
| Master's degree or above | 0.665 (0.221-1.995) | 0.467 |
| **Permanent address** | | |
| Town | 1 [Reference] | NA |
| City | 0.904 (0.459-1.781) | 0.770 |
| **Cosmetics use (per week)** | | |
| 0 days | 1 [Reference] | NA |
| 1-3 days | 1.417 (0.566-3.545) | 0.457 |
| 4-7 days | 2.615 (0.974-7.026) | 0.057 |
| **Take-out food consumption (per week)** | | |
| 0 times | 1 [Reference] | NA |
| 1-3 times | 3.129 (1.195-8.191) | **0.020** |
| 4-6 times | 2.987 (1.071-8.328) | **0.036** |
| 7-9 times | 4.148 (1.238-13.901) | **0.021** |

Abbreviations: NA, not applicable; OR, odds ratio; CI, confidence interval. Differences were considered significant at $P < 0.05$.

**Table S11. Univariable logistic regression analysis for the prevalence of pre-existing anti-PEG IgG and IgM in pregnant women (n = 256)**

| | Anti-PEG IgM | | Anti-PEG IgG | |
|---|---|---|---|---|
| **Variable** | **OR (95% CI)** | ***P* value** | **OR (95% CI)** | ***P* value** |
| **Maternal age** | 0.866 (0.783-0.958) | **0.005** | 0.942 (0.852-1.041) | 0.242 |
| **BMI**[a] | 1.046 (0.927-1.180) | 0.463 | 1.003 (0.880-1.144) | 0.961 |
| **Education** | | | | |
| Vocational degree or below | 1 [Reference] | NA | 1 [Reference] | NA |
| Associate's degree | 0.927 (0.311-2.761) | 0.892 | 1.167 (0.347-3.928) | 0.803 |
| Bachelor's degree | 0.704 (0.255-1.939) | 0.497 | 0.630 (0.190-2.092) | 0.451 |
| Master's degree or above | 0.341 (0.067-1.746) | 0.197 | 1.364 (0.365-5.097) | 0.645 |
| **Permanent address** | | | | |
| Town | 1 [Reference] | NA | 1 [Reference] | NA |
| City | 0.842 (0.362-1.957) | 0.689 | 1.244 (0.473-3.269) | 0.658 |
| **Cosmetics use (per week)** | | | | |
| 0 days | 1 [Reference] | NA | 1 [Reference] | NA |
| 1-3 days | 0.848 (0.239-3.011) | 0.798 | 2.021 (0.620-6.591) | 0.243 |
| 4-7 days | 0.879 (0.192-4.036) | 0.869 | 5.841 (1.947-17.558) | **0.002** |
| **Take-out food consumption (per week)** | | | | |
| 0 times | 1 [Reference] | NA | 1 [Reference] | NA |
| 1-3 times | 2.783 (0.875-8.854) | 0.083 | 3.477 (0.727-16.623) | 0.118 |
| 4-6 times | 2.026 (0.564-7.278) | 0.279 | 4.857 (0.991-23.802) | 0.051 |
| 7-9 times | 2.250 (0.467-10.845) | 0.312 | 8.500 (1.531-47.182) | **0.014** |

Abbreviations: NA, not applicable; OR, odds ratio; CI, confidence interval. Differences were considered significant at $P < 0.05$.

**Table S12. Univariable logistic regression analysis for the prevalence of pre-existing total anti-PEG antibodies in newborns (n = 256)**

| Variable | OR (95% CI) | *P* value |
|---|---|---|
| **Maternal age** | 0.963 (0.848-1.094) | 0.563 |
| **BMI**[a] | 1.113 (0.949-1.305) | 0.189 |
| **Education** | | |
| Vocational degree or below | 1 [Reference] | NA |
| Associate's degree | 0.807 (0.191-3.404) | 0.770 |
| Bachelor's degree | 0.564 (0.145-2.197) | 0.446 |
| Master's degree or above | 0.311 (0.033-2.901) | 0.305 |
| **Permanent address** | | |
| Town | 1 [Reference] | NA |
| City | 0.521 (0.174-1.558) | 0.243 |
| **Cosmetics use (per week)** | | |
| 0 days | 1 [Reference] | NA |
| 1-3 days | 0.000 (0.000-0.000) | 0.998 |
| 4-7 days | 7.259 (2.159-24.411) | **0.001** |
| **Take-out food consumption (per week)** | | |
| 0 times | 1 [Reference] | NA |
| 1-3 times | 1.473 (0.355-6.100) | 0.594 |
| 4-6 times | 1.893 (0.434-8.261) | 0.396 |
| 7-9 times | 0.000 (0.000-0.000) | 0.998 |
| **Gestational age at delivery** | 0.874 (0.461-1.656) | 0.679 |
| **Newborn gender** | | |
| Male | 1 [Reference] | NA |
| Female | 0.726 (0.244-2.154) | 0.563 |
| **Newborn weight** | 2.411 (0.616-9.436) | 0.206 |

Abbreviations: NA, not applicable; OR, odds ratio; CI, confidence interval. Differences were considered significant at $P < 0.05$.

**Figure S43. Correlation between the levels of pre-existing total anti-PEG antibodies, anti-PEG IgG and IgM with maternal age and BMI in pregnant women seropositive for anti-PEG antibodies (n = 49), anti-PEG IgG (n = 24) and IgM (n = 28)**

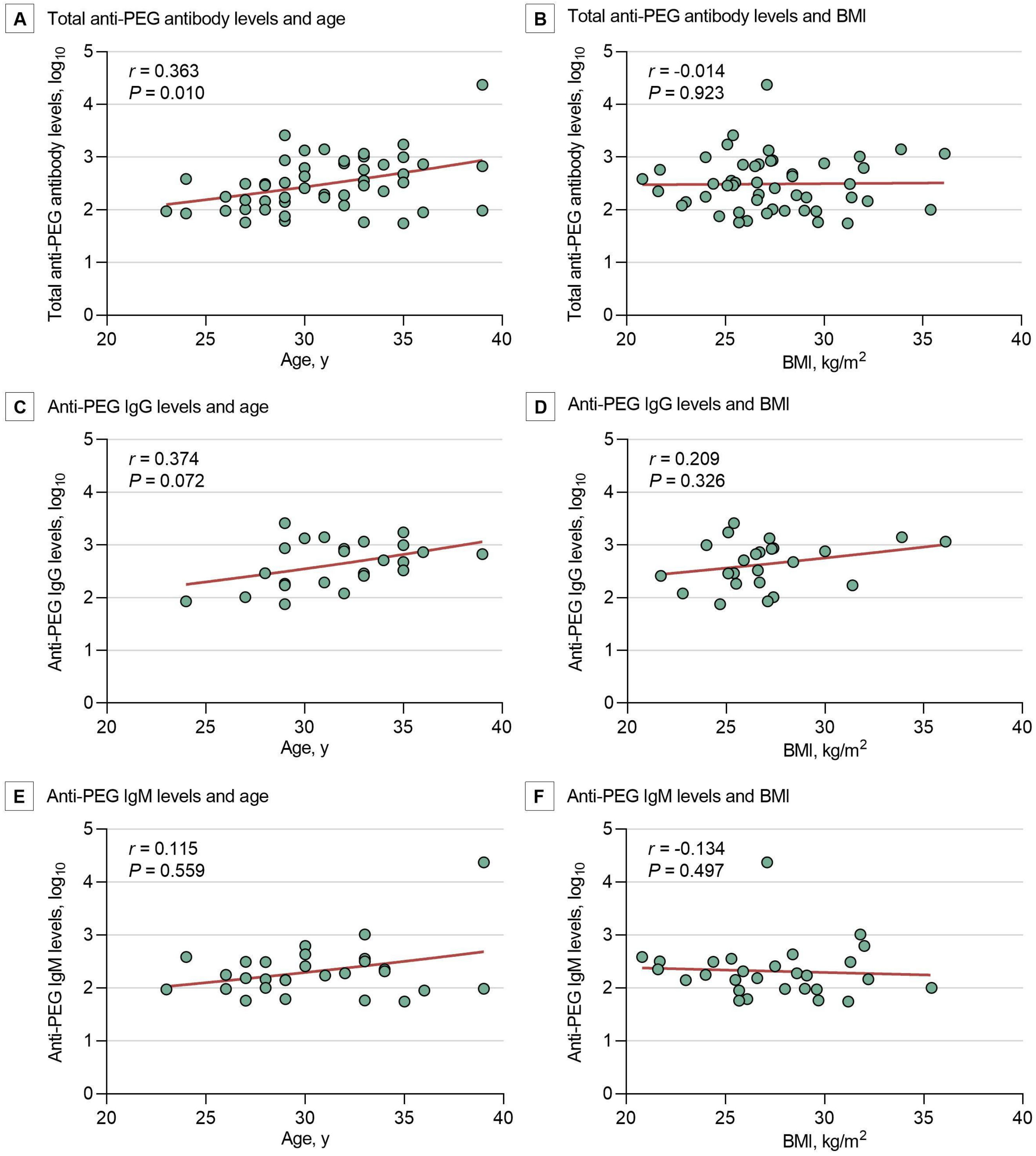


Abbreviations: *r*, correlation coefficients. For pregnant women seropositive for anti-PEG antibodies, correlations between anti-PEG antibody levels after $log_{10}$ transformation and each continuous variable (e.g., maternal age and BM) were analyzed using Spearman correlation coefficients (*r*). Differences were considered significant at $P < 0.05$.

**Table S13. Univariable generalized linear regression analysis for the levels of pre-existing total anti-PEG antibodies in pregnant women seropositive for anti-PEG antibodies (n = 49)**

| Variable | β (95% CI) | *P* value |
|---|---|---|
| **Maternal age** | 0.053 (0.018-0.088) | **0.003** |
| **BMI**[a] | 0.002 (-0.041-0.045) | 0.918 |
| **Education** | | |
| Vocational degree or below | 0 [Reference] | NA |
| Associate's degree | 0.050 (-0.350-0.449) | 0.808 |
| Bachelor's degree | 0.243 (-0.134-0.621) | 0.206 |
| Master's degree or above | 0.040 (-0.463-0.542) | 0.877 |
| **Permanent address** | | |
| Town | 0 [Reference] | NA |
| City | 0.158 (-0.151-0.467) | 0.317 |
| **Cosmetics use (per week)** | | |
| 0 days | 0 [Reference] | NA |
| 1-3 days | 0.163 (-0.221-0.547) | 0.406 |
| 4-7 days | 0.576 (0.191-0.960) | **0.003** |
| **Take-out food consumption (per week)** | | |
| 0 times | 0 [Reference] | NA |
| 1-3 times | 0.160 (-0.302-0.622) | 0.498 |
| 4-6 times | 0.141 (-0.349-0.630) | 0.573 |
| 7-9 times | 0.113 (-0.445-0.671) | 0.691 |

Abbreviations: NA, not applicable; β, standardized regression coefficient; CI, confidence interval. Differences were considered significant at $P < 0.05$.

**Table S14. Univariable generalized linear regression analysis for the levels of pre-existing anti-PEG IgG and IgM in pregnant women seropositive for anti-PEG IgG (n = 24) and IgM (n = 28)**

| | Anti-PEG IgM | | Anti-PEG IgG | |
|---|---|---|---|---|
| **Variable** | **β (95% CI)** | ***P* value** | **β (95% CI)** | ***P* value** |
| **Maternal age** | 0.039 (-0.006-0.084) | 0.087 | 0.057 (0.010-0.104) | **0.018** |
| **BMI** | -0.009 (-0.063-0.044) | 0.737 | 0.039 (-0.013-0.091) | 0.143 |
| **Education** | | | | |
| Vocational degree or below | 0 [Reference] | NA | 0 [Reference] | NA |
| Associate's degree | 0.173 (-0.304-0.650) | 0.477 | -0.114 (-0.577-0.348) | 0.628 |
| Bachelor's degree | 0.453 (0.005-0.900) | **0.047** | 0.064 (-0.398-0.527) | 0.786 |
| Master's degree or above | 0.321 (-0.418-1.060) | 0.395 | -0.346 (-0.845-0.154) | 0.175 |
| **Permanent address** | | | | |
| Town | 0 [Reference] | NA | 0 [Reference] | NA |
| City | 0.224 (-0.169-0.617) | 0.263 | -0.085 (-0.481-0.310) | 0.673 |
| **Cosmetics use (per week)** | | | | |
| 0 days | 0 [Reference] | NA | 0 [Reference] | NA |
| 1-3 days | 0.121 (-0.358-0.600) | 0.621 | 0.095 (-0.381-0.571) | 0.696 |
| 4-7 days | 1.220 (0.645-1.795) | **< 0.001** | 0.057 (-0.353-0.466) | 0.786 |
| **Take-out food consumption (per week)** | | | | |
| 0 times | 0 [Reference] | NA | 0 [Reference] | NA |
| 1-3 times | -0.188 (-0.733-0.358) | 0.500 | 0.702 (0.107-1.297) | **0.021** |
| 4-6 times | -0.224 (-0.827-0.379) | 0.466 | 0.595 (-0.006-1.196) | 0.053 |
| 7-9 times | -0.500 (-1.235-0.234) | 0.182 | 0.642 (0.006-1.279) | **0.048** |

Abbreviations: NA, not applicable; β, standardized regression coefficient; CI, confidence interval. Differences were considered significant at $P < 0.05$.

**Figure S44. Correlation between the levels of pre-existing total anti-PEG antibodies and maternal age, maternal BMI, gestational age at delivery, and newborn weight in newborns seropositive for anti-PEG antibodies (n = 14)**

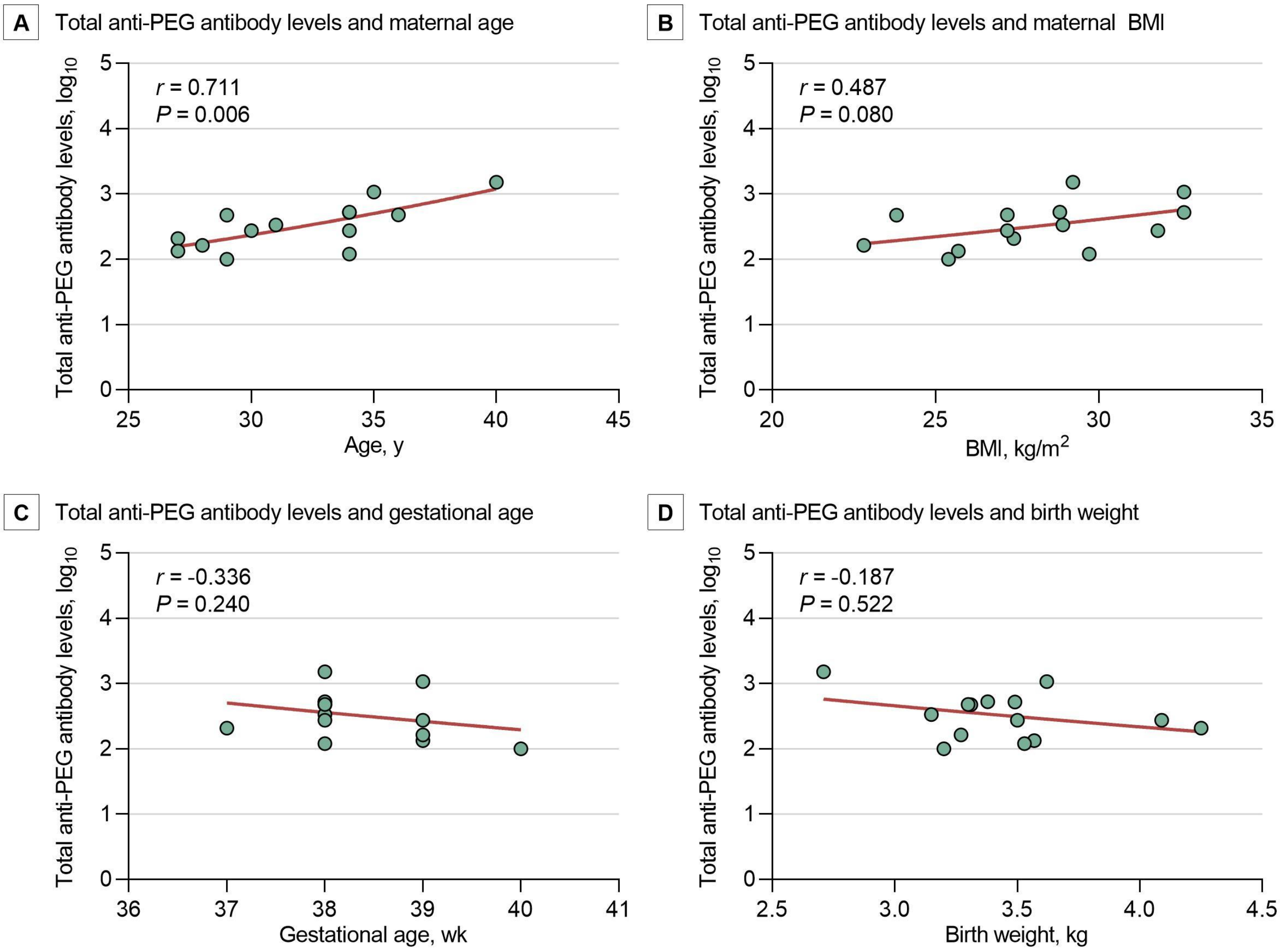


Abbreviations: *r*, correlation coefficients. For newborns seropositive for anti-PEG antibodies, correlations between anti-PEG antibody levels after $\log_{10}$ transformation and each continuous variable (e.g., maternal age, maternal BMI, gestation age at delivery, and newborn weight) were analyzed using Spearman correlation coefficients (*r*). Differences were considered significant at $P < 0.05$.

**Table S15. Univariable generalized linear regression analysis for the levels of pre-existing total anti-PEG antibodies in newborns seropositive for anti-PEG antibodies (n = 14)**

| Variable | β (95% CI) | *P* value |
|---|---|---|
| **Maternal age** | 0.065 (0.032-0.098) | **< 0.001** |
| **BMI** | 0.053 (-5.978E-5-0.106) | 0.050 |
| **Education** | | |
| Vocational degree or below | 0 [Reference] | NA |
| Associate's degree | 0.102 (-0.318-0.523) | 0.633 |
| Bachelor's degree | -0.110 (-0.509-0.289) | 0.590 |
| Master's degree or above | -0.475 (-1.139-0.190) | 0.162 |
| **Permanent address** | | |
| Town | 0 [Reference] | NA |
| City | -0.254 (-0.585-0.077) | 0.133 |
| **Cosmetics use (per week)** | | |
| 0 days | 0 [Reference] | NA |
| 4-7 days | -0.199 (-0.552-0.154) | 0.269 |
| **Take-out food consumption (per week)** | | |
| 0 times | 0 [Reference] | NA |
| 1-3 times | -0.189 (-0.635-0.257) | 0.406 |
| 4-6 times | -0.271 (-0.731-0.190) | 0.250 |
| **Gestational age at delivery** | -0.147 (-0.381-0.086) | 0.217 |
| **Newborn gender** | | |
| Male | 0 [Reference] | NA |
| Female | 0.074 (-0.281-0.428) | 0.684 |
| **Newborn weight** | -0.303 (-0.757-0.152) | 0.192 |

Abbreviations: NA, not applicable; β, standardized regression coefficient; CI, confidence interval. Differences were considered significant at $P < 0.05$.

### Figure S45. Overview diagram describing the design, procedures and major research findings of this study

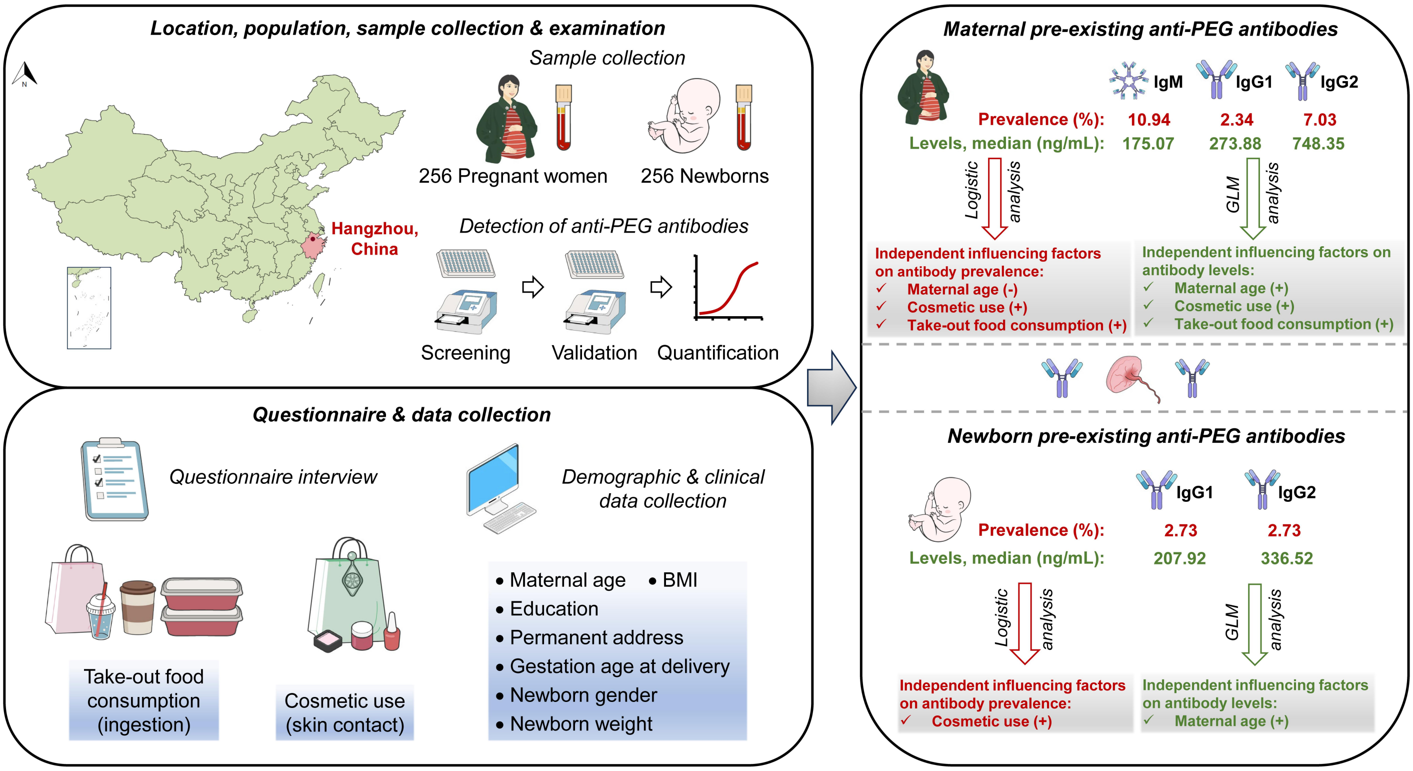


Abbreviations: (-), negative association with antibody prevalence; (+), positive association with antibody prevalence or levels. 256 pregnant women admitted for delivery at the Women's Hospital, Zhejiang University School of Medicine in China and their newborns were enrolled in this cross-sectional study, with corresponding maternal and cord blood samples carefully collected and examined for pre-existing anti-PEG antibodies. Using internationally recognized direct ELISA and competitive ELISA, along with questionnaire interviews, demographic and clinical data collections, and in-depth statistical analysis, the seropositivities and levels of pre-existing anti-PEG antibodies in pregnant women and corresponding newborns were revealed. Moreover, we found that maternal age, take-out food consumption, and cosmetic use were influencing factors of prevalence and levels of maternal anti-PEG antibodies, while the prevalence or levels of newborn anti-PEG antibodies were affected by maternal age and cosmetic use.

#### Table S16. Representative preclinical studies indicating accelerated blood clearance phenomenon of PEGylated Drugs

| Year[Ref] | Animal model | Treatment design | | | Isotype of anti-PEG antibodies induced by the initial dose |
|---|---|---|---|---|---|
| | | PEGylated agent for initial dose | PEGylated agent for subsequent dose | Dosing interval | |
| 2006[4] | Rat | PEGylated liposomes (IV) | Same as initial dose | 4, 5, 6, 7, 10 and 14 days | Anti-PEG IgM |
| 2007[5] | Rat | PEGylated liposomes (IV) | Same as initial dose | 5 days | Anti-PEG IgM |
| 2009[14] | Rat | PEG-modified PLA nanoparticles (IV) | Same as initial dose | 3, 5, 7 and 14 days | Anti-PEG IgM |
| 2012[15] | Mice | PEGylated polymeric micelles (IV) | Same as initial dose | 7 days | Anti-PEG IgM |
| 2012[6] | Mice | PEGylated liposomal topotecan (IV) | Same as initial dose | 7 days | Anti-PEG IgM |
| | Rat | PEGylated liposomal topotecan (IV) | Same as initial dose | 7 days | Anti-PEG IgM |
| | Dog | PEGylated liposomal topotecan (IV) | Same as initial dose | 7 days | Anti-PEG IgM |
| 2015[7] | Mice | PEGylated liposomes (IV) | Same as initial dose | 6 days | Anti-PEG IgM |
| | Rat | PEGylated liposomes (IV) | Same as initial dose | 6 days | Anti-PEG IgM |
| | Rabbit | PEGylated liposomes (IV) | Same as initial dose | 6 days | Anti-PEG IgM |
| | Guinea pig | PEGylated liposomes (IV) | Same as initial dose | 6 days | Anti-PEG IgM |
| 2015[11] | Mice | PEGylated ovalbumin (IV) | Same as initial dose | 5 days | Anti-PEG IgM |
| 2018[16] | Rat | Homemade PEGylated microbubble (IV) | Same as initial dose | 7 and 14 days | Anti-PEG IgM and IgG |
| | Rat | Definity®-PEGylated microbubble (IV) | Same as initial dose | 7 and 14 days | Anti-PEG IgM and IgG |
| 2019[8] | Rat | PEGylated liposomal gambogenic acid (IV) | Same as initial dose | 3, 5 and 7 days | Anti-PEG IgM |
| 2020[12] | Mice | Pegasys®-Peginterferon Alfa-2a (SC) | Same as initial dose | 7 days | Anti-PEG IgM |
| | Mice | PEGylated ovalbumin (IV) | Pegasys®-Peginterferon Alfa-2a (SC) | 7 days | Anti-PEG IgM |
| 2020[13] | Mice | Pegfilgrastim®-PEGylated human G-CSF (IV) | PEGylated liposomes (IV) | 5 days | Anti-PEG IgM |
| | Mice | PEGylated ovalbumin (IV) | Pegfilgrastim®-PEGylated human G-CSF (IV) | 5 days | Anti-PEG IgM |
| 2021[17] | Mice | PEGylated exosomes (IV) | Same as initial dose | 5 days | Anti-PEG IgM |
| | Mice | PEGylated liposomes (IV) | PEGylated exosomes (IV) | 5 days | Anti-PEG IgM |
| | Mice | PEGylated ovalbumin (IV) | PEGylated exosomes (IV) | 5 days | Anti-PEG IgM |
| | Mice | PEGylated exosomes (IV) | PEGylated liposomes (IV) | 5 days | Anti-PEG IgM |
| 2023[10] | Mice | PEG-based pharmaceutical excipients (IV) | PEGylated liposomes (IV) | 7 days | Anti-PEG IgM and IgG |
| | Mice | PEG-based pharmaceutical excipients (IV) | PEGylated liposomal doxorubicin (IV) | 7 days | Anti-PEG IgM and IgG |
| 2023[3] | Rat | PEGylated lipid nanoparticles (IM) | Same as initial dose | 21 days | Anti-PEG IgM |
| 2024[9] | Mice | A cosmetic product containing PEG derivatives (TOP) | Doxil®-PEGylated liposomal doxorubicin (IV) | 7 days | Anti-PEG IgM |
| | Mice | A cosmetic product containing PEG derivatives (TOP) | PEGylated liposomal oxaliplatin (IV) | 7 days | Anti-PEG IgM |

Abbreviations: IV, intravenous injection; SC, subcutaneous injection; IM, intramuscular injection; TOP, daily topical applications for 7 days; G-CSF, granulocyte colony-stimulating factor

#### Table S17. Clinical trials indicating accelerated blood clearance phenomenon of PEGylated Drugs

| Year[Ref] | Population | Clinical medication protocol | | | | | Correlation between treatment-induced anti-PEG antibodies and ABC phenomenon or reduced efficacy |
|---|---|---|---|---|---|---|---|
| | | Drug | Dose | Administration route | Interval | Duration | |
| 2006[19] | 13 patients with refractory gout | Krystexxa®[b] | 4 ~ 24 mg | SC | Single injection | NA | Five out of thirteen patients developed low-titer anti-PEG IgM within 3-7 days of drug treatment. Subsequently, between 7-14 days, these patients exhibited low titers of anti-PEG IgG. These antibodies were associated with ABC phenomenon. |
| 2010[18] | 28 children with acute lymphoblastic leukemia | Oncaspar®[a] | 1000 U/m$^2$ for each treatment | IV | 2 weeks | 8 times | Among serum samples of 15 Oncaspar®-treated patients with undetectable ASNase activity at 0 to 15 days after the last dose, anti-PEG antibodies were detected in 9 samples by serology and 12 samples by flow cytometry. Anti-PEG antibody was detected by flow cytometry in 1 Oncaspar®-treated patient with lower ASNase activity. |
| 2007[20] | 24 patients with refractory gout | Krystexxa®[b] | 0.5 ~ 12 mg | IV | Single injection | NA | Anti-PEG IgG was detected in serum samples of 9 gout patients, with drug clearance rates in these patients significantly accelerated. |
| 2014[21] | 30 patients with refractory gout (including 7 organ transplant recipients) | Krystexxa®[b] | 8 mg for each treatment | IV | 3 weeks | 5 times | Anti-PEG IgG was detected in serum samples of 13 gout patients, with drug clearance rates in these patients significantly accelerated. |
| 2014[22] | 169 patients with refractory gout | Krystexxa®[b] | 8 mg for each treatment | IV | 2 weeks | 12 times | Anti-PEG IgG and/or anti-PEG IgM were detected in serum samples of 67 gout patients, with drug clearance rates in these patients significantly accelerated. |
| 2018[23] | 24 patients with phenylketonuria | Palynziq®[c] | 2.5 mg for each treatment | IV | 1 week | Initiated for 4 times | Anti-PEG IgG and/or anti-PEG IgM were detected in serum samples of approximately 90% of phenylketonuria patients. These antibodies were associated with ABC phenomenon. |

Abbreviations: IV, intravenous injection; SC, subcutaneous injection; NA, not applicable.

[a]Oncaspar® (pegaspargase), a polyethylene glycol (PEG)-conjugated form of Escherichia coli-derived L-asparaginase, was granted FDA approval in 1994 for the treatment of acute lymphoblastic leukemia.

[b]Krystexxa® (pegloticase), a polyethylene glycol (PEG)-conjugated form of recombinant porcine uricase, was granted FDA approval in 2010 for the treatment of refractory chronic gout.

[c]Palynziq® (pegvaliase), a polyethylene glycol (PEG)-conjugated form of recombinant [Anabaena variabilis] phenylalanine ammonia lyase, was granted FDA approval in 2018 for the treatment of adult phenylketonuria.

**Figure S46. Screening of paired maternal and newborn serum samples negative for anti-PEG IgG and IgM**

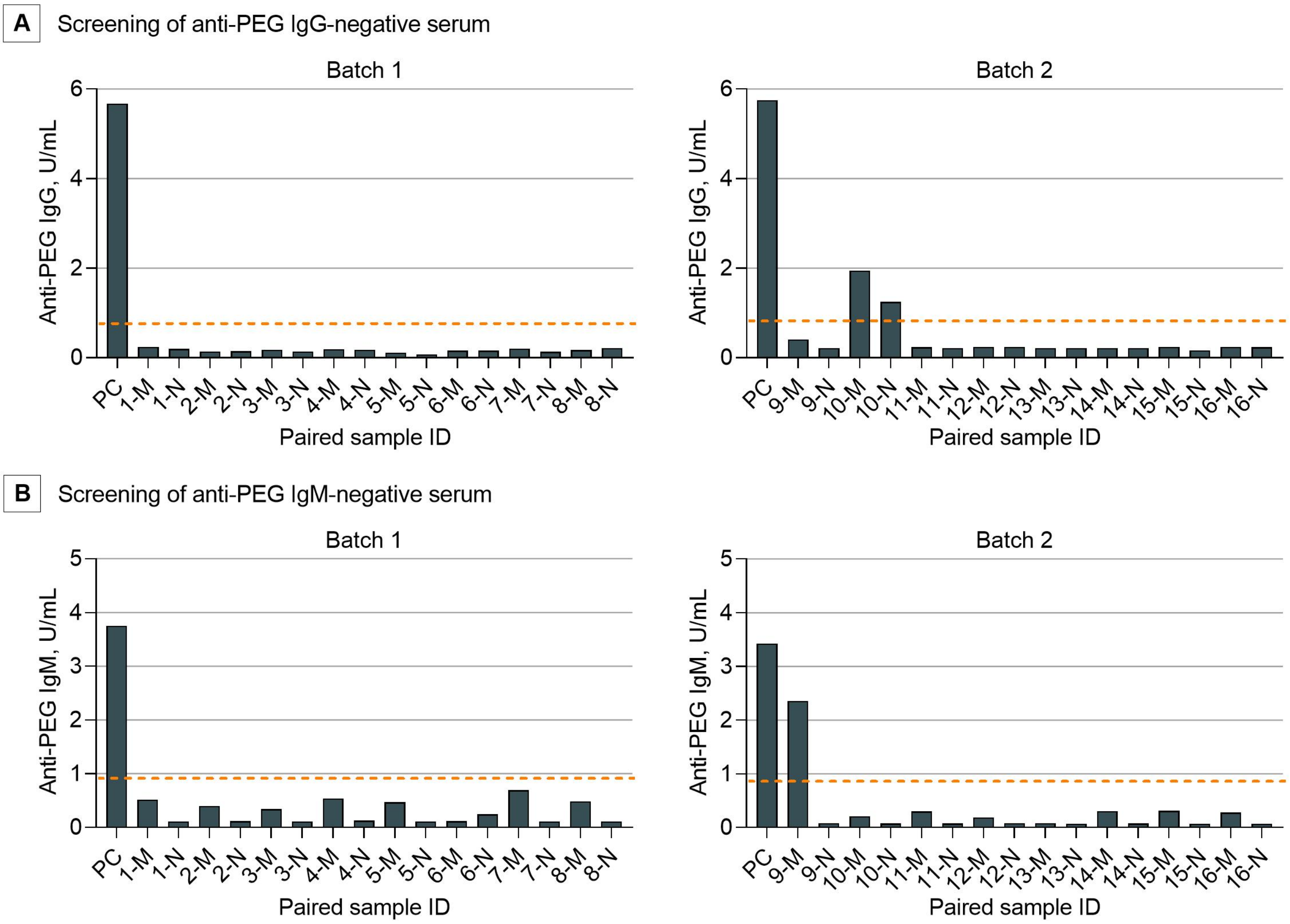


Abbreviations: PC, positive control included in commercial human anti-PEG IgM or IgG ELISA kits; M, maternal serum sample; N, newborn serum sample. Dotted lines indicate threshold values distinguishing between Positive/Negative in each ELISA for two independent batches.

**Figure S47. Confirmation of the absence of anti-PEG IgE in maternal and newborn serum samples negative for anti-PEG IgG and IgM**

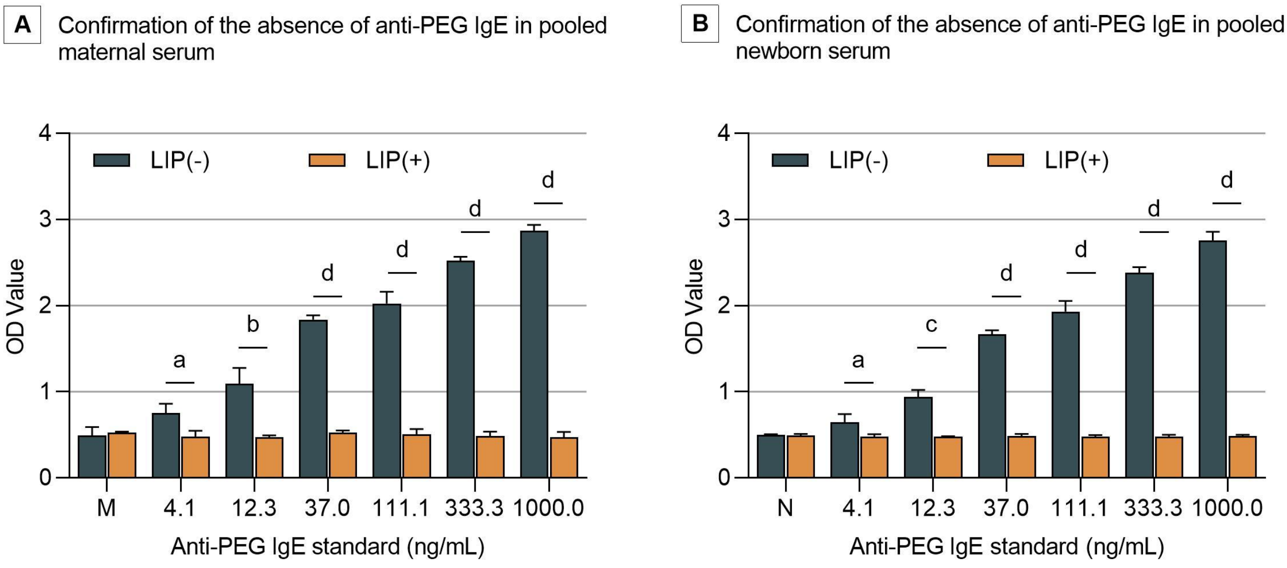


Abbreviations: LIP, PEGylated liposomal doxorubicin (competitors, see eMethods 4). LIP(-), the tested serum samples or anti-PEG IgE standard without the addition of LIP; LIP(+), the tested serum samples or anti-PEG IgE standard with the addition of LIP. M, pooled maternal serum samples negative for anti-PEG IgG and IgM; N, pooled newborn serum samples negative for anti-PEG IgG and IgM. All data were presented as "mean ± standard deviation" ($n = 3$), with differences between two groups analyzed using the two-tailed unpaired t-test. a, $P < 0.05$; b, $P < 0.01$; c, $P < 0.001$; d, $P < 0.0001$.
